# Vaccine Effectiveness Among 5- to 17-year-old Individuals with Prior SARS-CoV-2 Infection: An EHR-Based Target Trial Emulation Study from the RECOVER Project

**DOI:** 10.1101/2025.02.07.25321814

**Authors:** Jiajie Chen, Yuqing Lei, Qiong Wu, Ting Zhou, Bingyu Zhang, Michael J. Becich, Yuriy Bisyuk, Saul Blecker, Elizabeth A. Chrischilles, Dimitri A. Christakis, Lindsay G. Cowell, Mollie R. Cummins, Soledad A. Fernandez, Daniel Fort, Sandy Gonzalez, Sharon J. Herring, Benjamin D. Horne, Carol Horowitz, Mei Liu, Susan Kim, Parsa Mirhaji, Abu Saleh Mohammad Mosa, Jennifer A. Muszynski, Catharine I. Paules, Alice Sato, Hayden T. Schwenk, Soumitra Sengupta, Srinivasan Suresh, Bradley W. Taylor, David A. Williams, Yongqun He, Jeffrey S. Morris, Ravi Jhaveri, Christopher B Forrest, Yong Chen, RECOVER Consortium

## Abstract

**IMPORTANCE:** Prior studies have demonstrated the effectiveness of COVID-19 vaccines in children and adolescents. However, the benefits of vaccination in these age groups with prior infection remain underexplored.

**OBJECTIVE:** To evaluate the effectiveness of COVID-19 vaccination in preventing reinfection with various Omicron subvariants (BA.1/2, BA.4/5, XBB, and later) among 5- to 17-year-olds with prior SARS-CoV-2 infection.

**DESIGN:** A target trial emulation through nested designs with distinct study periods.

**SETTING:** The study utilized data from the Research COVID to Enhance Recovery (RECOVER) initiative, a national electronic health record (EHR) database comprising 37 U.S. children’s hospitals and health institutions.

**PARTICIPANTS:** Individuals aged 5-17 years with a documented history of SARS-CoV-2 infection prior to the study start date during a specific variant-dominant period (Delta, BA.1/2, or BA.4/5) who received a subsequent dose of COVID-19 vaccine during the study periods were compared with those with a documented history of infection who did not receive SARS-CoV-2 vaccine during the study period. Those infected within the Delta-Omicron composite period (December 1, 2021, to December 31, 2021) were excluded. The study period was from January 1, 2022, to August 30, 2023, and focused on adolescents aged 12 to 17 years and children aged 5 to 11 years.

**EXPOSURES:** At least received one COVID-19 vaccination during the study period vs. no receipt of any COVID-19 vaccine during the study period.

**MAIN OUTCOMES AND MEASURES:** The primary outcome is documented SARS-CoV-2 reinfection during the study period (both asymptomatic and symptomatic cases). The effectiveness of the COVID-19 vaccine was estimated as (1-hazard ratio) *100%, with confounders adjusted by a combination of propensity score matching and exact matching.

**RESULTS:** The study analyzed 87,573 participants during the BA.1/2 period, 229,326 during the BA.4/5 period, and 282,981 during the XBB or later period. Among vaccinated individuals, significant protection was observed during the BA.1/2 period, with effectiveness rates of 62% (95% CI: 38%-77%) for children and 65% (95% CI: 32%-81%) for adolescents. During the BA.4/5 period, vaccine effectiveness was 57% (95% CI: 25%-76%) for children, but not statistically significant for adolescents (36%, 95% CI: −16%-65%). For the XBB period, no significant protection was observed in either group, with effectiveness rates of 22% (95% CI: −36%-56%) in children and 34% (95% CI: −10%-61%) in adolescents.

**CONCLUSIONS AND RELEVANCE:** COVID-19 vaccination provides significant protection against reinfection for children and adolescents with prior infections during the early and mid-Omicron periods. This study also highlights the importance of addressing low vaccination rates in pediatric populations to enhance protection against emerging variants.

**Key Points:** *Question:* Does COVID-19 vaccination protect against reinfection with Omicron subvariants (BA.1/2, BA.4/5, XBB, and later) among children and adolescents aged 5-17 years with prior SARS-CoV-2 infection?

*Findings:* COVID-19 vaccination significantly reduced the risk of reinfection during the BA.1/2 period, with effectiveness rates of 62% in children and 65% in adolescents. During the BA.4/5 period, vaccination provided moderate protection in children, with effectiveness rates of 57%, but the effectiveness in adolescents (36%) was not statistically significant. For the XBB period and later subvariants, vaccination did not show significant protection in either group.

*Meaning:* COVID-19 vaccination offered meaningful protection against reinfection in pediatric populations during earlier Omicron periods but became less effective with later emerging subvariants, highlighting the challenges of sustaining vaccine effectiveness as the virus continues to evolve.

## Introduction

The rapid development and distribution of COVID-19 vaccines against SARS-CoV-2 in response to the global pandemic represent a remarkable scientific and public health achievement. During the early stages of the pandemic, extensive research established the effectiveness of these vaccines in preventing severe disease, hospitalization, and death in individuals without prior SARS-CoV-2 exposure.^1–10^ As the pandemic progressed, numerous variants of SARS-CoV-2 emerged, often causing less severe infections, at least partially because of the widespread population immunity from both vaccination and previous infections. The Centers for Disease Control and Prevention (CDC) has recommended COVID-19 vaccination for individuals with prior SARS-CoV-2 infections^11^, underscoring the potential benefits of hybrid immunity—a combination of infection-acquired and vaccine-induced immunity.

Emerging evidence on hybrid immunity indicates that such individuals often exhibit superior and more durable immune responses compared to those with either immunity type alone^12–19^. However, pediatric studies on hybrid immunity are limited, especially those stratified by SARS-CoV-2 variants. Preliminary findings suggest that hybrid immunity may confer enhanced protection against immune-evasive variants such as Delta, Omicron BA.1, and BA.2, but these findings are predominantly derived from adult populations^20,21^. Notably, a study from Qatar reported that hybrid immunity provided the strongest protection against symptomatic Alpha, Beta, and Delta infections, assuming independent contributions of infection- and vaccine-induced immunity across age groups^22^. However, subsequent analysis found no significant differences in protection among prior infection, vaccination, and hybrid immunity against symptomatic Omicron BA.1 and BA.2 infections^23^. Further insights come from a population-based cohort study^24^ that included participants aged 12 and more, which demonstrated that three or more antigenic exposures — achieved through vaccination or infection — significantly reduced the risk of Omicron-associated hospitalization and death across all age groups. Despite the Omicron variant’s rapid global dissemination since November 2021, research quantifying the effects of hybrid immunity against diverse Omicron subvariants, i.e., B.A. 1/2, B.A. 4/5, XBB, and later, is lacking. Addressing this gap is critical to tailoring vaccination strategies for younger populations as the SARS-CoV-2 landscape continues to evolve.

Despite the global dominance of the Omicron variant since November 2021, research quantifying the effects of hybrid immunity against specific Omicron subvariants—such as BA.1/2, BA.4/5, and XBB—is limited. Addressing this gap is crucial for refining vaccination strategies tailored to younger populations as the SARS-CoV-2 landscape continues to evolve.

This study aims to investigate the understudied population of children and adolescents with prior SARS-CoV-2 infection—a significant group given the delayed vaccine rollout for these age groups, during which many had already acquired natural immunity. Using electronic health record (EHR) data from the Research COVID to Enhance Recovery (RECOVER) Project, we evaluate COVID-19 vaccine effectiveness in previously infected children and adolescents. This study represents the largest to date in the United States on vaccine effectiveness in this population, leveraging data from 37 U.S. children’s hospitals and health institutions. Employing a target trial emulation design, we assess vaccine effectiveness against reinfection, encompassing both asymptomatic and symptomatic cases, and stratify our analysis by different Omicron subvariant periods. To strengthen the robustness of our findings, negative control experiments were performed to adjust for potential unmeasured confounding factors.

The results of this study will provide critical insights for informing vaccination strategies and public health policies targeting pediatric populations with prior infection-induced immunity, ultimately contributing to more effective prevention of future infections in these groups.

## Methods

### Data Sources

This study is part of the National Institutes of Health (NIH) funded RECOVER Initiative (https://recovercovid.org/), which aims to learn about the long-term effects of COVID-19. This study included 37 US children’s hospitals and health institutions. The EHR data was standardized to the OMOP Common Data Model (CDM) ^25^ and extracted from the RECOVER Database Version S11. More details are available in the Supplementary Materials **Section S1**.

### Specification and Emulation of Target Trials

This study employed target trial emulation techniques^26–28^ to investigate the effectiveness of the COVID-19 vaccine in preventing SARS-CoV-2 reinfections among previously infected children and adolescents in the United States. To comprehensively assess vaccine effectiveness, we designed six distinct target trials stratified by age group (children aged 5-11 years and adolescents aged 12-17 years) and by the prevalence of specific SARS-CoV-2 Omicron subvariants during the study period. This stratification allows for a more precise evaluation of how vaccine protection varies across different viral landscapes and developmental stages. As detailed in **Table 1**, the six target trials were organized across three major Omicron variant periods.

**Table 1.**
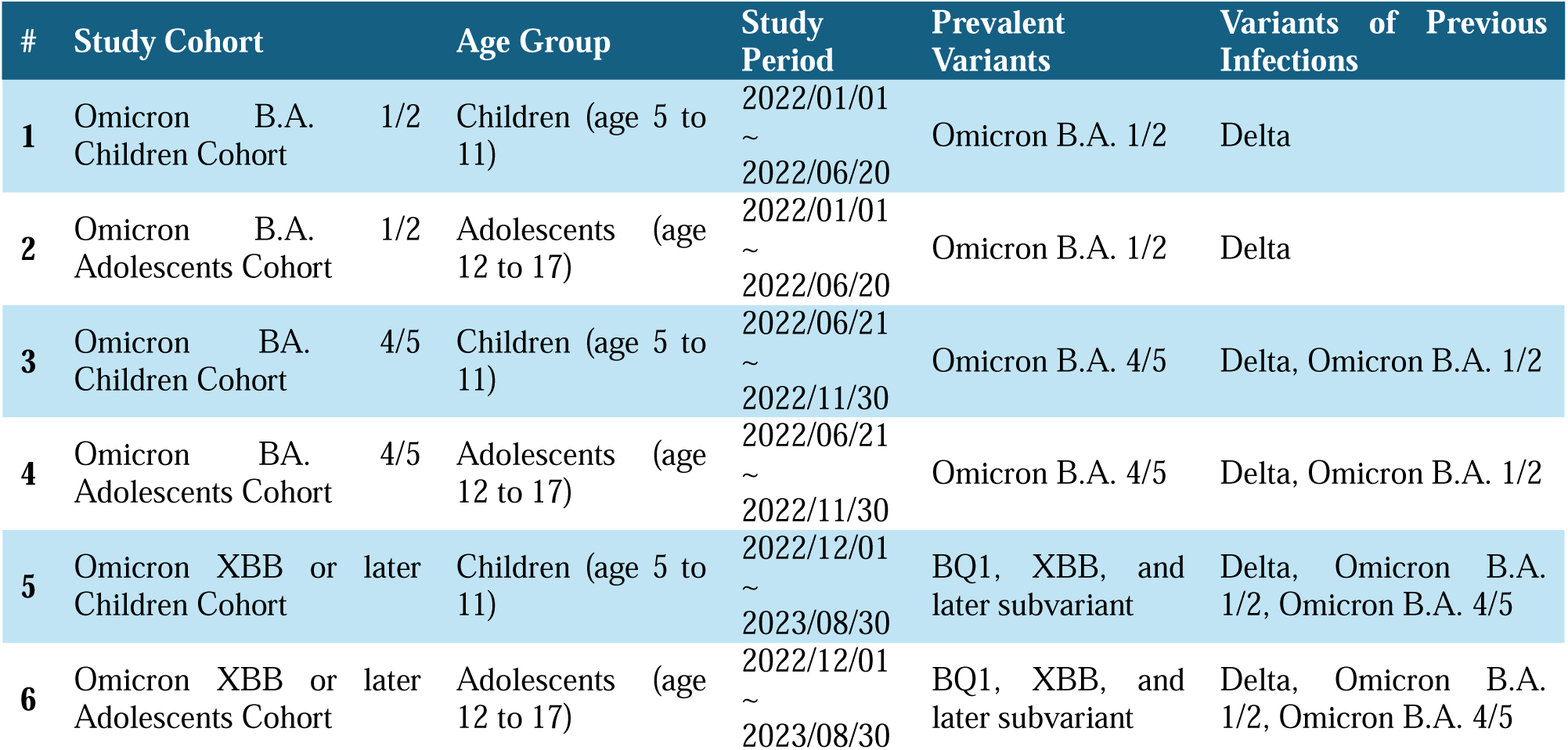
Target Trial Design Stratified by Age Group and Prevalent SARS-CoV-2 Omicron Subvariants During the Study Period.

### Study Design and Population

For each target trial, eligibility criteria included children aged 5 to 11 years and adolescents aged 12 to 17 years at the start of the study period. Eligible participants had a documented SARS-CoV-2 infection during a specific variant-dominant period (Delta, BA.1/2, or BA.4/5) before the study start date. Individuals younger than 5 or older than 18 years, or those infected during the Delta-Omicron overlap (December 1-31, 2021), were excluded. First infections were confirmed by a positive polymerase chain reaction (PCR) test, serology, antigen test, clinical diagnosis, a prescription for nirmatrelvir/ritonavir, or documentation of post-acute sequelae of SARS-CoV-2 (PASC).

The intervention of interest was receiving at least one dose of a COVID-19 vaccine during the study period, compared to no vaccination during the study period. Target trial emulation was conducted every 7 days within each study period, creating sequential 7-day trials. Participants could initially belong to the comparator group before receiving vaccination. For each 7-day trial, participants were required to remain uninfected and unvaccinated during the study period up until the trial start date. Vaccinated participants without infection during the trial were assigned to the intervention group, with the index date as their vaccination date. Participants who remained unvaccinated or were infected before vaccination during the trial were placed in the comparator group, with the trial start date as their index date. Participants with both infection-acquired and vaccine-induced immunity during the study period were categorized as having hybrid immunity.

### Outcomes

The primary outcome was documented SARS-CoV-2 reinfection during each study period. Reinfection was identified by a subsequent positive PCR or antigen test, clinical diagnosis, or prescription for nirmatrelvir/ritonavir, occurring at least 60 days after the previous infection. Reinfections within 60 days were considered ongoing infections. Participants were followed from their index date until the earliest of the following: documented SARS-CoV-2 reinfection, matched participant’s reinfection, matched participant’s vaccination, or the study period’s end.

### Statistical Analysis

To balance baseline characteristics, we applied exact matching and propensity score matching to the intervention and comparator groups. We first performed exact matching of each eligible participant who is vaccinated within the 7-days trial period to all eligible participants who were unvaccinated during the trial period using factors including gender, race/ethnicity (Hispanic, Non-Hispanic White, Non-Hispanic Black, Asian American/Pacific Islander, Other), indicators from the data-contributing sites, number of COVID-19 vaccine doses before cohort entry, and time since the last vaccination. Prior infection characteristics, including the variant and time since infection, were also matched. Subsequently, within the exact matching strata, propensity scores were estimated using logistic regression models, adjusting for age, obesity status, and chronic conditions defined by the Pediatric Medical Complexity Algorithm (PMCA)^29^. Conditions included cardiovascular, respiratory, neurological, and other system-specific diseases. Healthcare utilization, medication records, days since the last infection, and prior infection severity were also incorporated. Up to 1:5 (at least 1:1) matching with replacement was used within each nested sequential trial. An unvaccinated comparator could serve as a match for vaccinated persons in more than one sequential trial as long as they remained unvaccinated. Detailed variable definitions are provided in **Table S1** of the Supplementary Appendix. **Figure 1** provides a visual demonstration of the target trial emulation design used in this study. It illustrates the sequential 7-day trial framework, where vaccinated individuals were compared with matched unvaccinated controls.

**Figure 1.**
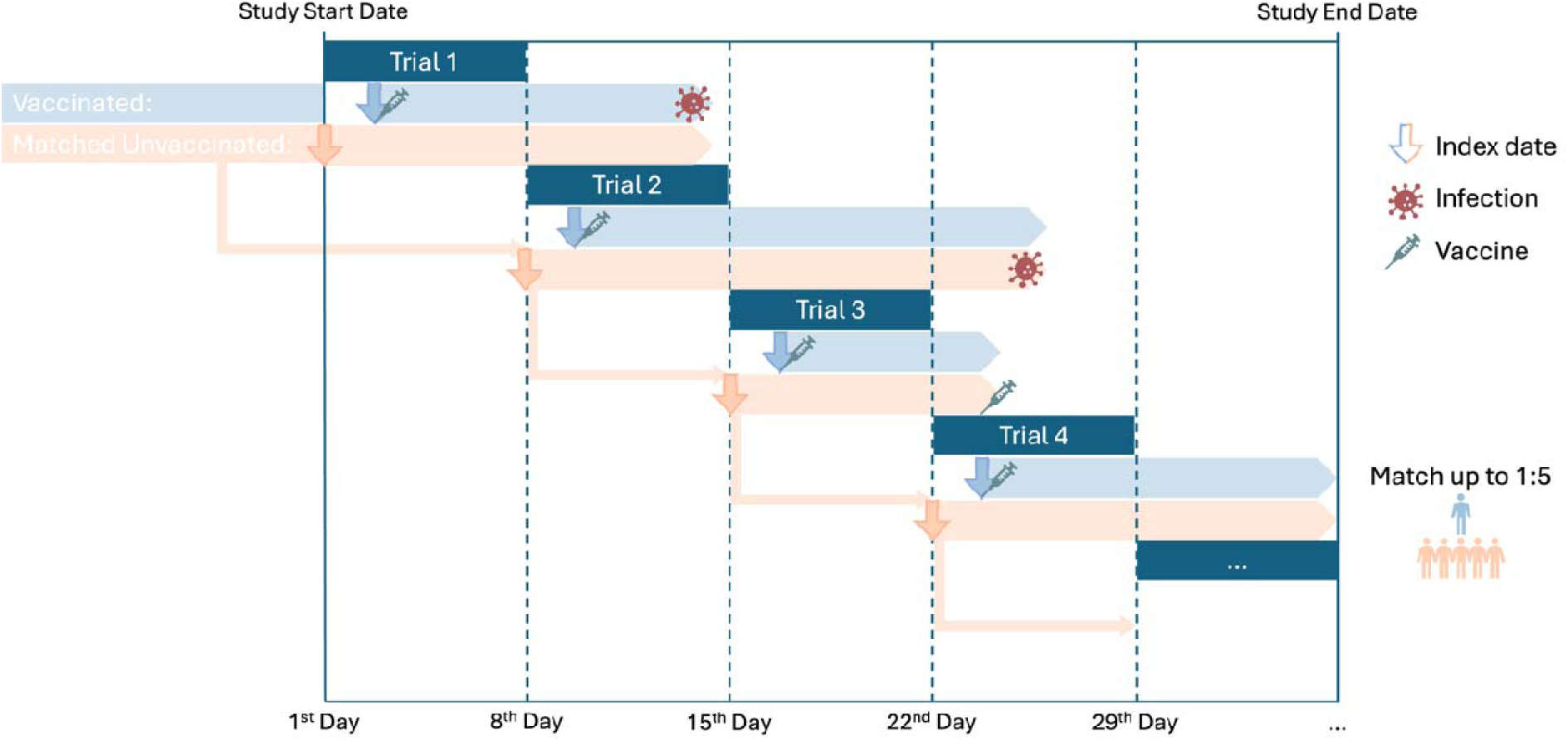
Study design for the emulation of a target trial evaluating the effectiveness of COVID-19 vaccination versus no vaccination in preventing SARS-CoV-2 reinfection among previously infected children and adolescents. Participants were enrolled in sequential 7-day trials, with vaccinated individuals compared to matched unvaccinated controls. The index date was defined as the vaccination date for the intervention group and the trial start date for the comparator group. Up to 1:5 matching with replacement was used.

We used Cox proportional hazards models to estimate vaccine effectiveness, defined as 100*(1 - hazard ratio), with 95% confidence intervals (CIs). This analysis examined the relationship between post-infection vaccination and two outcomes: SARS-CoV-2 reinfection and symptomatic reinfection. We emulated 25 target trials for the B.A.1/2 period, 24 for B.A.4/5, and 39 for XBB/later variants, pooling participants across each study period for analysis. Detailed statistical methods are described in **Section S2** of the Supplementary Appendix.

### Sensitivity Analysis

To assess the robustness of our findings, we conducted a series of sensitivity analyses. First, we stratified the analysis by the last infected variant, as detailed in Supplementary Materials **Section S3 part A**. Second, we evaluated vaccine effectiveness against varying severities of COVID-19 illness (mild, moderate, severe), which is reported in **Section S3 Part B**. Additionally, as shown in Supplementary **Figure S19**, the majority of vaccinated participants in the study cohorts received the Pfizer COVID-19 vaccine. Across all cohorts, Pfizer accounted for more than 90% of vaccinations, with only a small proportion receiving Moderna or unspecified vaccines. This dominant use of the Pfizer vaccine justified conducting a sensitivity analysis restricted to Pfizer recipients, which presented in **Section S3 Part C**. Finally, we implemented negative control outcome experiments using a list of 40 negative control outcomes, determined by board-certified pediatricians, to calibrate residual study bias from unmeasured confounders, assuming the null hypothesis of no effect. The empirical null distribution and calibrated risks are presented in **Section S3 Part D**.

All analyses were performed using R software, version 4.1.0 (R Foundation).

## Results

### Study Population

A total of 48,249 children and 39,324 adolescents from the Omicron BA.1/2 cohort, 123,417 children and 105,909 adolescents from the Omicron BA.4/5 cohort, and 151,961 children and 131,020 adolescents from the Omicron XBB or later cohort within the RECOVER Database were identified to study the effectiveness of vaccination against reinfection during the subsequent variant periods. Baseline characteristics are detailed in **Table 2**.

**Table 2.**
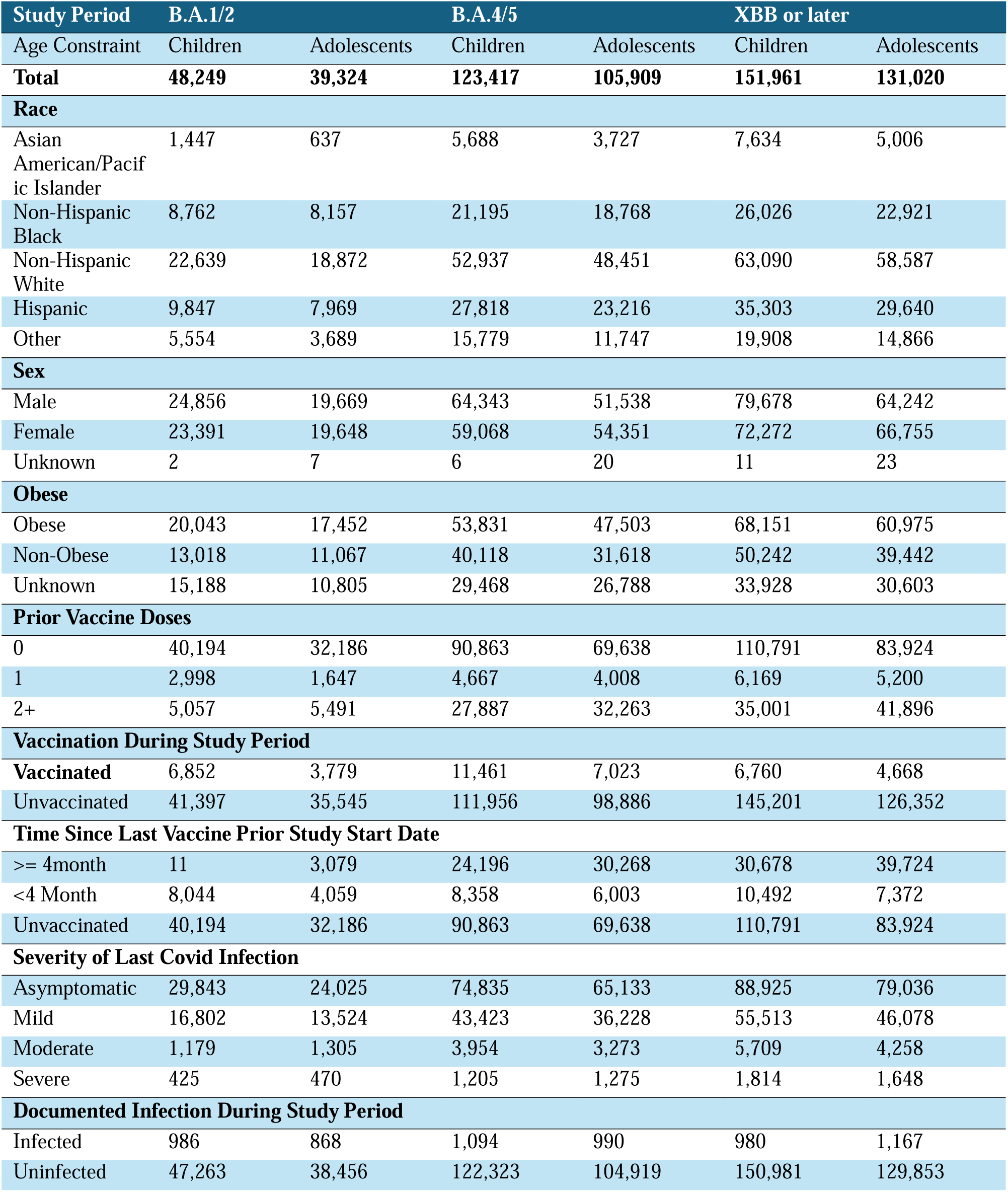
Baseline characteristics of patients before matching.

Across all cohorts, the majority of both children and adolescents were Non-Hispanic White, with 53.0% of children and 51.4% of adolescents in the B.A.1/2 cohort, 52.1% of children and 50.7% of adolescents in the B.A.4/5 cohort, and 51.2% of children and 50.3% of adolescents in the XBB or later cohort. Hispanic participants were the second-largest group, comprising 20.4% of children and 23.7% of adolescents in the B.A.1/2 cohort, 22.5% of children and 23.8% of adolescents in the B.A.4/5 cohort, and 21.8% of children and 23.5% of adolescents in the XBB or later cohort. The prevalence of obesity was higher among adolescents compared to children, ranging from 43.0% to 46.5% in adolescents and 40.0% to 43.8% in children across cohorts. Most participants (over 83%) had received no prior COVID-19 vaccine doses before the study period. During the study period, 16.6% of children and 9.6% of adolescents in the B.A.1/2 cohort, 9.3% of children and 6.6% of adolescents in the B.A.4/5 cohort, and 7.2% of children and 5.1% of adolescents in the XBB or later cohort received at least one dose of the COVID-19 vaccine. The remaining majority of participants remained unvaccinated throughout the respective study periods. Documented SARS-CoV-2 reinfections during the study period were rare. In the BA.1/2 cohort, reinfections occurred in 2.0% of children and 2.2% of adolescents; in the BA.4/5 cohort, 0.9% of children and 0.9% of adolescents experienced reinfection; and in the XBB or later cohort, 0.6% of children and 0.9% of adolescents had documented reinfections.

### Vaccine Effectiveness

The COVID-19 vaccination demonstrated effectiveness in preventing SARS-CoV-2 reinfection among children and adolescents during the Omicron B.A.1/2 and B.A.4/5 periods, but reduced effectiveness was observed during the XBB and later period.

**Figure 2** summarizes the estimated vaccine effectiveness in sequential trials emulations framework for 6 target trials. For B.A.1/2 study cohort, 8,712 (5,667 children and 3,045 adolescents) were eligible for the vaccinated group for the vaccine effectiveness analysis against B.A. 1/2, and 34,394 (22,154 children and 12,240 adolescents) were matched to unvaccinated controls. For the B.A.4/5 study cohort, 16,589 (10,295 children and 6,294 adolescents) were eligible for the vaccinated group for the vaccine effectiveness analysis against B.A.4/5, and 72,920 (45,491 children and 27,429 adolescents) were matched to unvaccinated controls. For the XBB or later study cohort, 9,962 (5,964 children and 3,998 adolescents) were eligible for the vaccinated group for the vaccine effectiveness analysis against XBB or later, and 42,734 (25,542 children and 17,192 adolescents) were matched to unvaccinated controls. The analysis results showed those who received at least one dose of the COVID-19 vaccine during the study period demonstrated statistically significant protection against BA.1/2 variant infection, with an effectiveness of 62.3% (95% CI: 38.3% to 77.0%) in children aged 5 to 11 years and 64.5% (95% CI: 32.4% to 81.3%) in adolescents aged 12 to 18 years. For the BA.4/5 variant, the vaccine conferred protection of 57.1% (95% CI: 24.6% to 75.6%) in children, while in adolescents, the protection was not statistically significant (36.5%, 95% CI: -15.7% to 65.1%). Additionally, the COVID-19 vaccine did not confer statistically significant protection against the XBB variant in either children (22.5%, 95% CI: -36.2% to 55.9%) or adolescents (34.5%, 95% CI: -10.3% to 61.2%).

**Figure 2.**
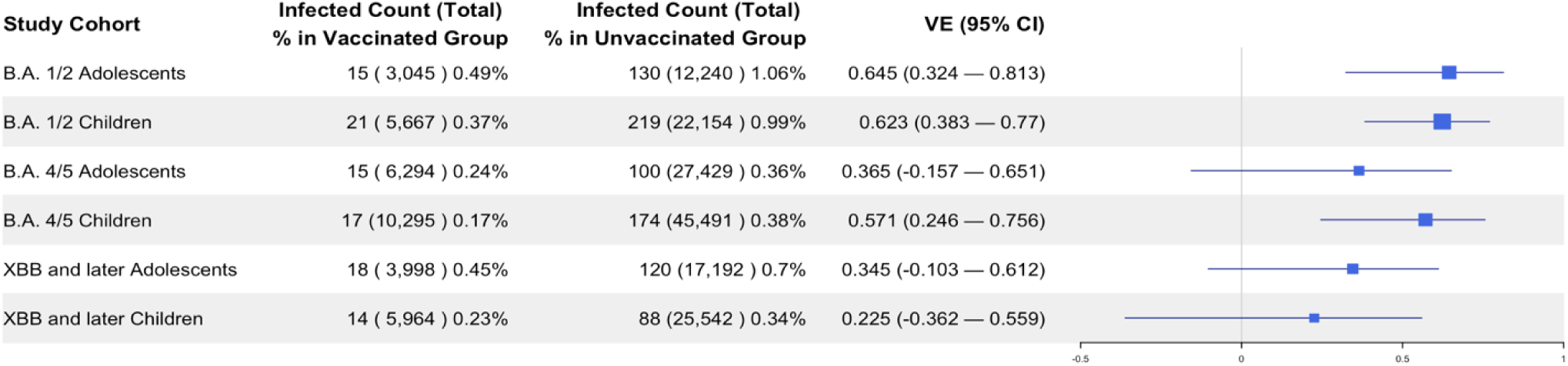
Vaccine Effectiveness Against SARS-CoV-2 Reinfection Across Omicron Subvariants (BA.1/2, BA.4/5, XBB and Later) in Vaccinated and Unvaccinated Children and Adolescents.

### Sensitivity Analysis

The sensitivity analysis was firstly stratified based on the last infected variants of the participants, which yielded that the COVID-19 vaccine provided significant protection against the Omicron BA.1/2 variant in both children and adolescents previously infected with the Delta variant. However, vaccine effectiveness against subsequent Omicron sub-variants, including BA.4/5 and XBB, showed non-significant results, particularly in adolescents. These inconsistencies in vaccine effectiveness are largely because of the low numbers of observed outcomes for these specific variants, which likely impacted the analysis. The detailed results are presented in **Section S3.A** of the Supplementary Appendix.

In the sensitivity analysis assessing vaccine effectiveness in preventing symptomatic COVID-19 illness among children and adolescents with prior SARS-CoV-2 infections, severity outcomes were categorized as asymptomatic, mild, moderate, or severe, following Forrest et al (2022)^30^. The incidence rates are summarized in Supplementary **Table S3**. The analysis showed that the COVID-19 vaccine had positive effectiveness in preventing symptomatic illness during the Omicron BA.1/2 and BA.4/5 periods, as presented in Supplementary **Figure S38**.

Moreover, the sensitivity analysis restricted to Pfizer recipients yielded results consistent with the primary analysis, confirming similar patterns of vaccine effectiveness across all study cohorts (Supplementary **Figure S39**).

Finally, negative control experiments (**Section S3. D**) indicated the presence of a slight positive systematic error, as evidenced by a minor shift in point estimates to the right (Supplementary **Table S7**). This rightward shift suggests a potential deflation of the primary results, meaning the vaccine effectiveness might be slightly underestimated.

## Discussion

This target trial emulation study represents the largest investigation into hybrid immunity in a pediatric cohort, examining the effectiveness of the COVID-19 vaccine in children and adolescents with a history of SARS-CoV-2 infection. Significant protection against the Omicron B.A.1/2 and B.A.4/5 variants was demonstrated, highlighting the importance of vaccination even in those previously infected. Our findings indicate that vaccine effectiveness varied across different Omicron sub-variants, with reduced efficacy, particularly against the newer XBB and later variant. The results underscore the challenges of maintaining vaccine efficacy amidst evolving viral landscapes.

Despite the benefits of vaccination, the persistently low vaccination rates in the pediatric population with prior infection has remained a significant public health concern. This is particularly troubling given the observed differences in vaccine effectiveness between age groups. Children consistently showed higher protection rates than adolescents, which may be attributed to age-related variations in immune responses. Younger children typically exhibit more robust immunological responses compared to older adolescents.^31^ Additionally, the timing of vaccine availability plays a critical role; Pfizer’s vaccine was initially approved for individuals aged 16 and older in December 2020, and eligibility expanded to include children aged 12-15 approximately six months later, in May 2021.^32,33^ As a result, older adolescents may have experienced a longer time interval since their initial vaccinations, potentially contributing to reduced vaccine effectiveness. These results reinforce the need for tailored vaccination strategies that account for the baseline immunity conferred by prior infections.

Hybrid immunity can provide enhanced protection against SARS-CoV-2, including various Omicron subvariants such as B.A.1/2, B.A.4/5, and XBB. The synergistic effect of hybrid immunity is attributed to the broader and more robust immune responses it elicits, including increased antibody diversity and durability, along with heightened T-cell responses. Studies suggest that hybrid immunity leads to a more comprehensive recognition of viral epitopes, enabling effective neutralization even against subvariants with significant mutations in the spike protein.^34,35^ Additionally, evidence indicates that hybrid immunity reduces the risk of severe outcomes and may also protect against long COVID.^36^ However, hybrid immunity can also present challenges. Individuals with prior infections might experience immune system overactivation upon vaccination, potentially leading to heightened inflammatory responses or rare adverse events.^37^ Furthermore, the immunological landscape created by hybrid immunity is complex and variable, influenced by factors such as the timing of infection relative to vaccination and the variant involved in the prior infection.^38^ Understanding these dynamics is crucial for tailoring vaccination strategies, particularly in pediatric populations where immune responses may differ significantly from adults.

Our findings regarding the estimated vaccine effectiveness among prior infected participants were consistent with those reported in other studies.^23,24^ On the other hand, our study’s design did not capture unreported home infections of mild severity. Such cases, if undetected, can lead to an overestimation of vaccine effectiveness. The focus on medically detectable infections only further contributes to this limitation, emphasizing the need for a more comprehensive surveillance and reporting system to capture the full spectrum of COVID-19 infection outcomes. The implied upside of this observation is that infections managed at home and not in any medical setting are likely to be minor, so the estimates of vaccine effectiveness may be inflated due to the lack of mild infection data.

### Limitations

The study has several limitations. The reliance on electronically recorded health data may omit less severe infection events not requiring medical attention, potentially skewing the perceived effectiveness of the vaccine. Additionally, the focus on medically detectable infections might not fully reflect the actual burden of disease, particularly for mild and asymptomatic infections. Future research should aim to evaluate the long-term effectiveness of COVID-19 vaccines in pediatric populations, particularly in the context of emerging variants. Studies that assess the impact of vaccination on mild and asymptomatic infections are also needed to provide a more complete picture of vaccine-induced protection. Such research will help refine vaccination strategies and inform public health policies designed to control the pandemic among younger populations.

## Conclusions

The COVID-19 vaccine appears to be effective for children and adolescents with prior COVID-19 infection during the Omicron period. This study also underscores the need to address the persistently low vaccination rates in the pediatric population, highlighting an area for public health improvement to enhance protection against emerging variants.

## Data Availability

The data used in this study are derived from electronic health records and contain sensitive patient information. Due to privacy regulations and institutional policies, these data cannot be shared publicly. Access to the dataset is restricted and subject to ethical and regulatory approvals.

## Disclosures

### Disclaimer

This content is solely the responsibility of the authors and does not necessarily represent the official views of the RECOVER Initiative, the NIH, or other funders.

## Acknowledgements

This study is part of the NIH Researching COVID to Enhance Recovery (RECOVER) Initiative, which seeks to understand, treat, and prevent the post-acute sequelae of SARS-CoV-2 infection (PASC). For more information on RECOVER, visit https://recovercovid.org/.

We would like to thank the Patient Representative: Leyna Aragon and Doug Lindsay, the National Community Engagement Group (NCEG), all patient, caregiver and community Representatives, and all the participants enrolled in the RECOVER Initiative.

