## Supplement for "Vaccine Effectiveness Among 5- to 17-year-old Individuals with Prior SARS-CoV-2 Infection: An EHR-Based Target Trial Emulation Study from the RECOVER Project"

Supplementary Appendix to:

**Vaccine Protection among pediatric population with naturally acquired immunity: a Target Trial Emulation in the RECOVER Project (TBD)**

### Section S1 Data description

#### A. Description of electronic health records data

The real-world data utilized in our analysis is derived from electronic health records (EHRs), covering a wide range of healthcare interaction information routinely collected and stored by hospitals. This includes clinical data such as diagnoses and treatments, laboratory and test results, and administrative data including patient demographics and billing information. The hospital-based EHR data from the Researching COVID to Enhance Recovery (RECOVER) Initiative COVID-19 Database served as the basis for defining and determining exposure, outcomes, and covariates. Unlike General Practitioner (GP) data, self-reported data, or external data sources, our study used the structured, standardized EHR entries made by healthcare providers within hospital settings. EHR data provides a more detailed and integrated view of a patient's health status, medical history, and healthcare interactions across various providers and settings.

#### B. RECOVER population and generalizability

The National Institutions of Health (NIH) launched the new RECOVER initiative in 2021 to leverage electronic health record (EHR) data to better estimate the SARS-CoV-2 related topics. RECOVER obtains EHRs from three large national healthcare networks within the United States, covering regional catchment areas across 41 states. These networks collectively hold the EHRs of over 60 million patients, including records from more than 7 million individuals who have been affected by COVID-19. RECOVER collaborates with the National Institutes of Health's (NIH) All of Us Research Program, which contributes additional health records to this vast database. Together, these sources comprise one of the world's largest collections of EHRs.

In our study, participating institutions in this study included: Ann & Robert H. Lurie Children’s Hospital of Chicago, Children's Hospital of Colorado, Children's Hospital of Philadelphia, Children's National Medical Center, Cincinnati Children’s Hospital Medical Center, Columbia University Irving Medical Center, Duke University, InterMountain Healthcare, Medical College of Wisconsin, Montefiore, Nationwide Children’s Hospital, Nemours Children’s Health System (in Delaware and Florida), Nicklaus Children's Hospital, Northwestern University, OCHIN, Inc., Ochsner Health System, Seattle Children’s Hospital, Stanford Children’s Health, Temple University, University of California, San Francisco, University of Iowa Healthcare, University of Michigan, University of Missouri, University of Nebraska Medical Center, University of Pittsburgh/UPMC, University of Utah, UT Southwestern Medical Center, Vanderbilt University Medical Center, Wake Forest Baptist Health, Weill Cornell Medicine Collage, Ochsner LSU Health Shreveport, Emory University, New York University Langone Health, Ohio State University, Penn State University, University of Florida Health. For this study, we used the s11 version of the data, collected till the end of August 2023.

### Section S2 Supplemental Methods

#### A. Study Cohort Definition

In this retrospective study, we first assessed eligibility criteria to extract general cohorts, which served as the foundation for subsequent trial emulation. The general cohorts included patients with at least one documented infection by a specific variant prior to the study’s start date. Eligible participants were restricted to those aged 5 to 18 years, further stratified into two sub-cohorts: children aged 5–11 years and adolescents aged 12–18 years. Patients with documented infections occurring between December 1, 2021, and December 31, 2022, corresponding to the Omicron-Delta mixed period, were excluded to avoid potential confounding. Ultimately, six cohorts were identified for analysis, each with study periods specified as outlining:

1. **Omicron B.A. 1/2 Children Cohort**
   - Age Constrain: aged 5 to 11 years
   - Previous Infected Period: Delta variant was prevalent (2021/07/01 ~ 2021/11/30)
   - Study Period: when Omicron B.A. 1/2 was prevalent (2022/01/01 ~ 2022/06/20)
2. **Omicron B.A. 1/2 Adolescents Cohort**
   - Age Constrain: aged 12 to 18 years
   - Previous Infected Period: Delta variant was prevalent (2021/07/01 ~ 2021/11/30)
   - Study Period: when Omicron B.A. 1/2 was prevalent (2022/01/01 ~ 2022/06/20)
3. **Omicron BA.4/5 Children Cohort**
   - Age Constrain: aged 5 to 11 years
   - Previous Infected Period:
     - Delta variant was prevalent (2021/07/01 ~ 2021/11/30)
     - Omicron B.A. 1/2 variant was prevalent (2022/01/01 ~ 2022/06/20)
   - Study Period: when Omicron B.A. 4/5 was prevalent (2022/06/21 ~ 2022/11/30)
4. **Omicron BA.4/5 Adolescents Cohort**
   - Age Constrain: aged 12 to 18 years
   - Previous Infected Period:
     - Delta variant was prevalent (2021/07/01 ~ 2021/11/30)
     - Omicron B.A. 1/2 variant was prevalent (2022/01/01 ~ 2022/06/20)
   - Study Period: when Omicron B.A. 4/5 was prevalent (2022/06/21 ~ 2022/11/30)
5. **Omicron XBB or later Children Cohort**
   - Age Constrain: aged 5 to 11 years
   - Previous Infected Period:
     - Delta variant was prevalent (2021/07/01 ~ 2021/11/30)
     - Omicron B.A. 1/2 variant was prevalent (2022/01/01 ~ 2022/06/20)
     - Omicron B.A. 4/5 was prevalent (2022/06/21 ~ 2022/11/30)
   - Study Period: BQ1, XBB, and later subvariant (2022/12/01 ~ 2023/08/30)
6. **Omicron XBB or later Adolescents Cohort**
   - Age Constrain: aged 12 to 18 years
   - Previous Infected Period:
     - Delta variant was prevalent (2021/07/01 ~ 2021/11/30)
     - Omicron B.A. 1/2 variant was prevalent (2022/01/01 ~ 2022/06/20)
     - Omicron B.A. 4/5 was prevalent (2022/06/21 ~ 2022/11/30)
   - Study Period: BQ1, XBB, and later subvariant (2022/12/01 ~ 2023/08/30)

#### B. Sequential Trials Emulation

We implemented the sequential trial emulation pipeline^1^ to analyze six cohorts, where vaccination status was sequentially defined and assigned. Trial emulations were performed every seven days throughout the study period. The eligibility criteria for each trial included the following:

- In the age group at the start of each trial date, i.e. age constraint
- No infection and no vaccination from study start date until the trial start date

Eligibility for the vaccinated and unvaccinated groups and the corresponding index dates in each trial emulation were defined as:

- Eligible for Treated (Vaccinated) group:
  - Vaccinated during the trial period
  - No reinfection from trial start date until the date of vaccination
  - Index date: vaccination date
- Eligible for Untreated (Unvaccinated) group:
  - No vaccination during the trial period, or reinfection prior to date of vaccination
  - Index date: the trial start date

To balance the intervention and comparator groups, we employed propensity score matching and exact matching at a ratio of up to 1:5, accounting for a wide range of measured confounders. The detailed definitions of the study variables are provided in **Table S1**.

Follow-up began on the index date and continued until one of the following events: documented SARS-CoV-2 reinfection, the next documented reinfection of a matched participant, the vaccination record of a matched participant, or the end of the study period

#### C. Study Variables

**Table S1**: Variables used in the emulation of study cohorts

| Variable | Functional form | Values | Detail | Codes/references |
| --- | --- | --- | --- | --- |
| Treatment  (i.e., Exposure) |  |  |  |  |
| COVID-19  vaccination | Indicator | Yes/No | Based on the immunization domain, using either the presence of a CVX code designating an administered or patient-reported dose or of a source value containing the terms “COVID” or “SARS” in the immunization table. | CVX code: 207, 208, 210, 211, 212, 213, 217, 218 |
| Outcomes |  |  |  |  |
| Documented Reinfection | Indicator | Yes/No | Based on the observation and visit occurrence domains. Defined as a polymerase-chain-reaction (PCR), serology, or antigen tests positive for COVID-19, or diagnoses of COVID-19, or a prescription for nirmatrelvir/ritonavir, provided these occurred at least 60 days after the previous infection prior to the study period. Cases with reinfection records within 60 days were considered ongoing infections and not new infection events. | See <https://github.com/PEDSnet/PASC/tree/main/observation_derivation_recover_ml_phenotype/specs> for detailed codes. |
| Mild/Moderate/Severe COVID-19 | Indicator | Yes/No | Based on the condition occurrence, procedure occurrence, drug, device and death domains. | Forrest, C. B., Burrows, E. K., Mejias, A., Razzaghi, H., Christakis, D., Jhaveri, R., Lee, G. M., Pajor, N. M., Rao, S., Thacker, D., & Bailey, L. C. (2022). Severity of Acute COVID-19 in Children &amp;lt;18 Years Old March 2020 to December 2021. *Pediatrics*, *149*(4). <https://doi.org/10.1542/peds.2021-055765>  Code sets are available through https://github.com/PEDSnet/COVID19_Severity/tree/main. |
| Confounding variables |  |  |  |  |
| Age (years) | Linear | NA | Based on records in the person domain. | Age is defined as the integer of (date – birth date)/365.25 |
| Sex | Indicator | Male/Female | Based on records in the person domain. | NA |
| Race/Ethnicity | 4 categories | Non-Hispanic White  Non-Hispanic Black  Hispanic  Other/unknown | Based on records in the person domain. | NA |
| Site | 37 Categories | Listed in Section S1 Part B. | Based on records in the site domain. | https://pcornet.org/network/ |
| Obesity | Indicator | Yes/No | Based on records in the measurement domain.  If measured at age < 24*30.5 days, NHANES weight z score > 1.64  If measured at 24*30.5 < age <240*30.5, NHANES BMI z score > 1.64  If measured at age >= 240*30.5, BMI kg/m2 > 30 | Zhou, T., Zhang, B., Zhang, D., Wu, Q., Chen, J., Li, L., ... & Chen, Y. (2024). Body Mass Index and Post acute Sequelae of SARS-CoV-2 Infection in Children and Young Adults. *JAMA Network Open*, *7*(10), e2441970-e2441970. |
| PMCA-defined body systems | 18 categories | cardiac, craniofacial, dermatological, endocrinological, gastrointestinal, genetic, genitourinary, hematological, immunological, malignancy, mental health, metabolic, musculoskeletal, neurological, ophthalmological, otologic, pulmonary respiratory (excluding asthma)  renal. | Based on the condition occurrence and visit occurrence domains. | Simon, T. D., Haaland, W., Hawley, K., Lambka, K., & Mangione-Smith, R. (2018). Development and validation of the Pediatric Medical Complexity Algorithm (PMCA) version 3.0. *Academic pediatrics*, *18*(5), 577-580. |
| Number of visits to emergency department in 18 months to 7 days prior to the entry | 4 categories | 0  1  2  >=3 | Based on the condition occurrence and visit occurrence domains. | NA |
| Number of inpatient visits in 18 months to 7 days prior to the entry | 4 categories | 0  1  2  >=3 | Based on the condition occurrence and visit occurrence domains, including Inpatient Hospital Stay, Emergency Department Admit to Inpatient Hospital Stay, and Observation Stay | NA |
| Number of outpatient visits in 18 months to 7 days prior to the entry | 4 categories | 0  1  2  >=3 | Based on the condition occurrence and visit occurrence domains including Ambulatory/Outpatient Visit (With a Physician) and Interactive Telemedicine Service | NA |
| Number of unique medications in 18 months to 7 days prior to the entry | 4 categories | 0  1  2  >=3 | Based on the drug exposure domain | RxNorm code |
| Number of COVID-19 vaccine doses prior to the entry | 4 categories | 0/1/2/≥3 | Based on the immunization domain, using either the presence of a CVX code designating an administered or patient-reported dose or of a source value containing the terms “COVID” or “sars” in the immunization table. | CVX code: 207, 208, 210, 211, 212, 213, 217, 218 |
| Interval since the last COVID-19 immunization date | 3 categories | No vaccination  <4 months  ≥4 months | Based on the immunization domain | NA |
| Days since the last infection date | Linear | NA | Based on the immunization domain. | NA |
| Index of the severity for last infection | Indicator | Yes/No | Based on the condition occurrence, procedure occurrence, drug, device and death domains. | Forrest, C. B., Burrows, E. K., Mejias, A., Razzaghi, H., Christakis, D., Jhaveri, R., Lee, G. M., Pajor, N. M., Rao, S., Thacker, D., & Bailey, L. C. (2022). Severity of Acute COVID-19 in Children & < 18 Years Old March 2020 to December 2021. *Pediatrics*, *149*(4). <https://doi.org/10.1542/peds.2021-055765>  Code sets are available through https://github.com/PEDSnet/COVID19_Severity/tree/main. |
| Last Variant infected prior to the entry | 3 categories | Delta,  Omicron B.A. 1/2  Omicron B.A. 4/5 | Based on the observation and visit occurrence domains. | NA |

Note: All of the domains in the table above are based on the OMOP common data model (CDM). More details are available through this link: <https://data-models-service.research.chop.edu>.

#### D. Detailed participant characteristics at baseline

To complement **Table 1**, which outlines the baseline characteristics of each study cohort, we provide additional details on the distribution of key participant attributes. These include race (**Figure S1–S6**), the number of prior vaccine doses (**Figure S7–S12**), the time since the last vaccine dose before the study start date (**Figure S13–S18**), and the vaccine brand administered to vaccinated participants during the study period (**Figure S19**).

**Figure S1: Distribution of Race for Omicron B.A. 1/2 Children Cohort.**
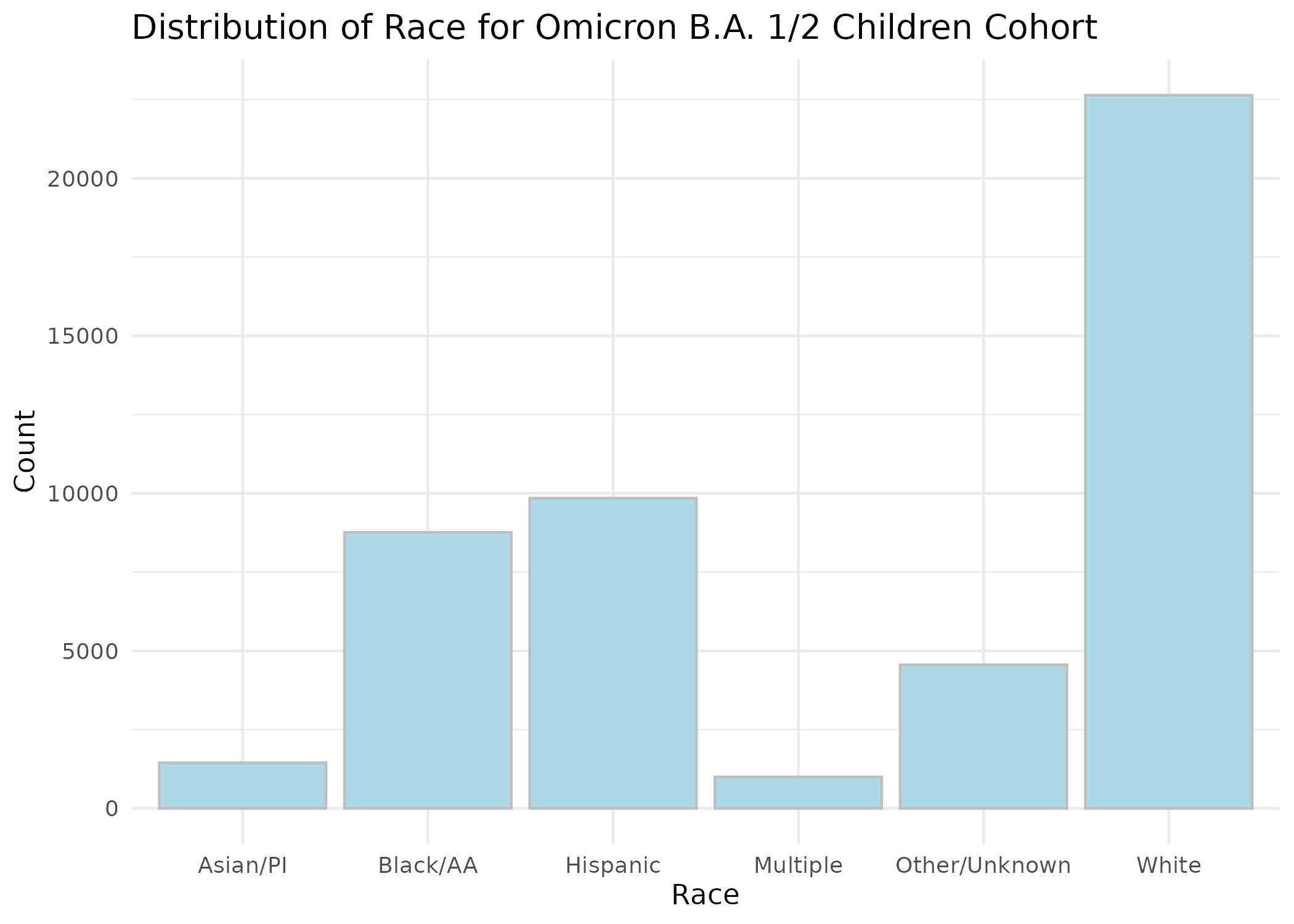

**Figure S2: Distribution of Race for Omicron B.A. 1/2 Adolescent Cohort.**
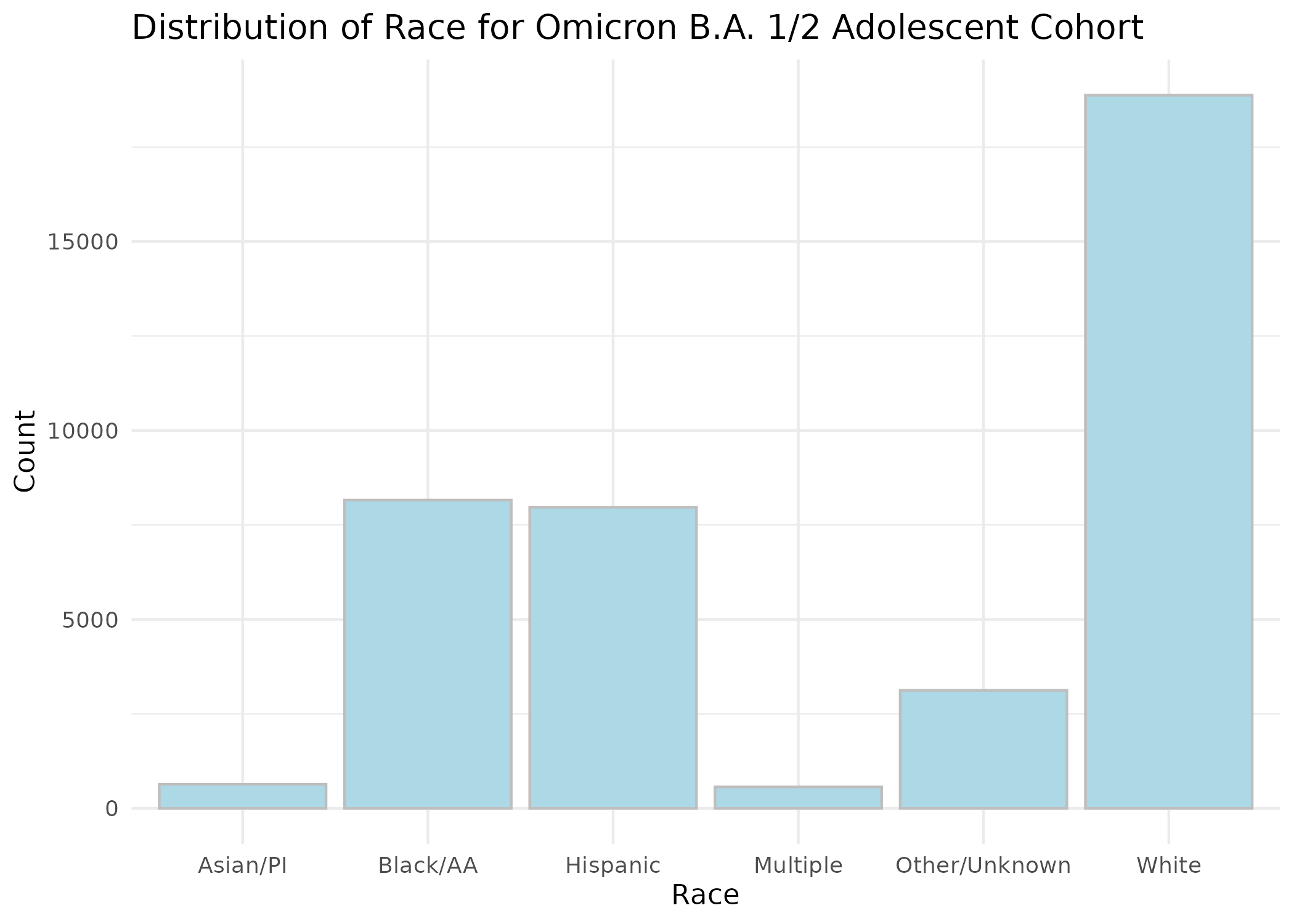

**Figure S3: Distribution of Race for Omicron B.A. 4/5 Children Cohort.**
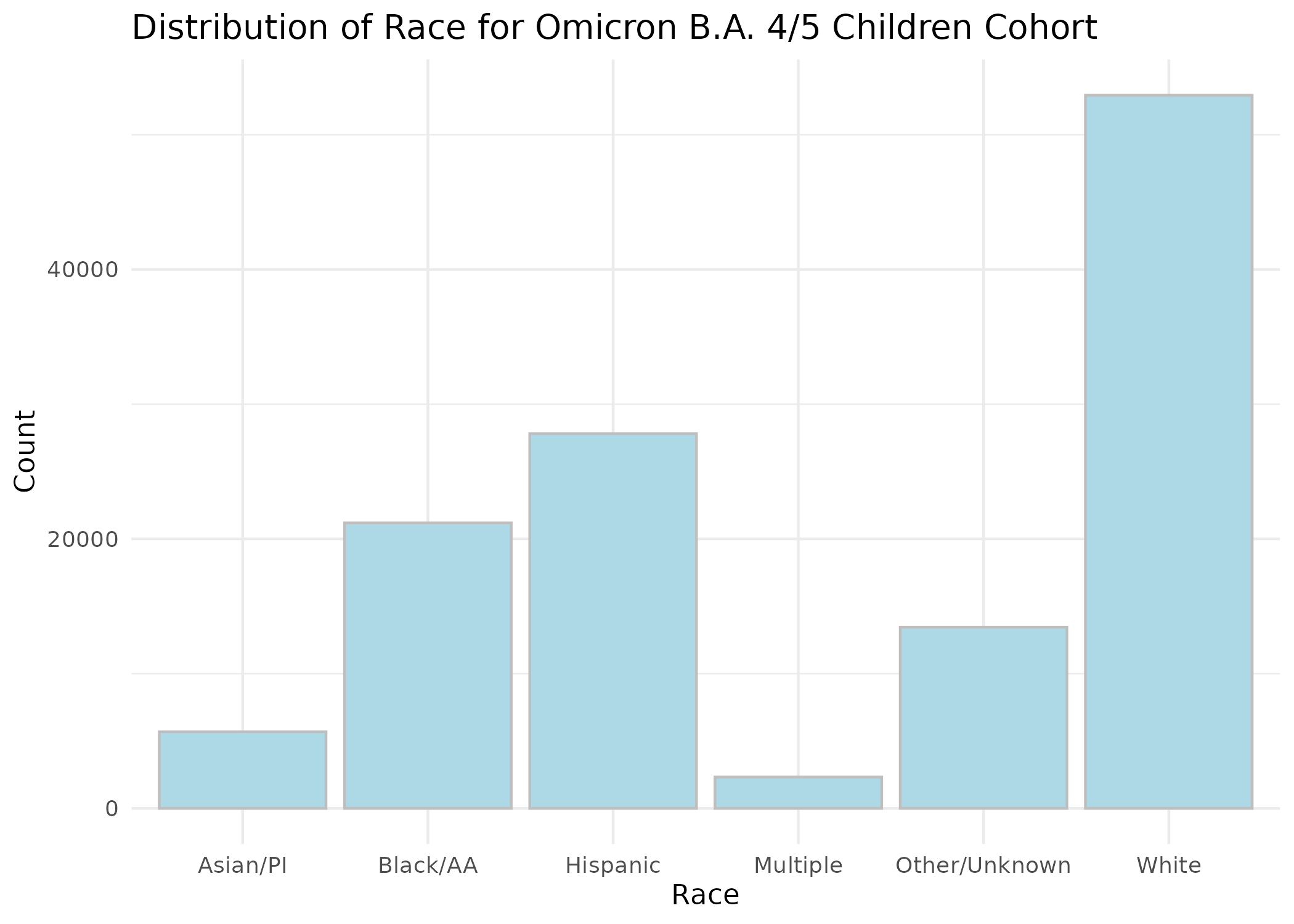

**Figure S4: Distribution of Race for Omicron B.A. 4/5 Adolescent Cohort.**
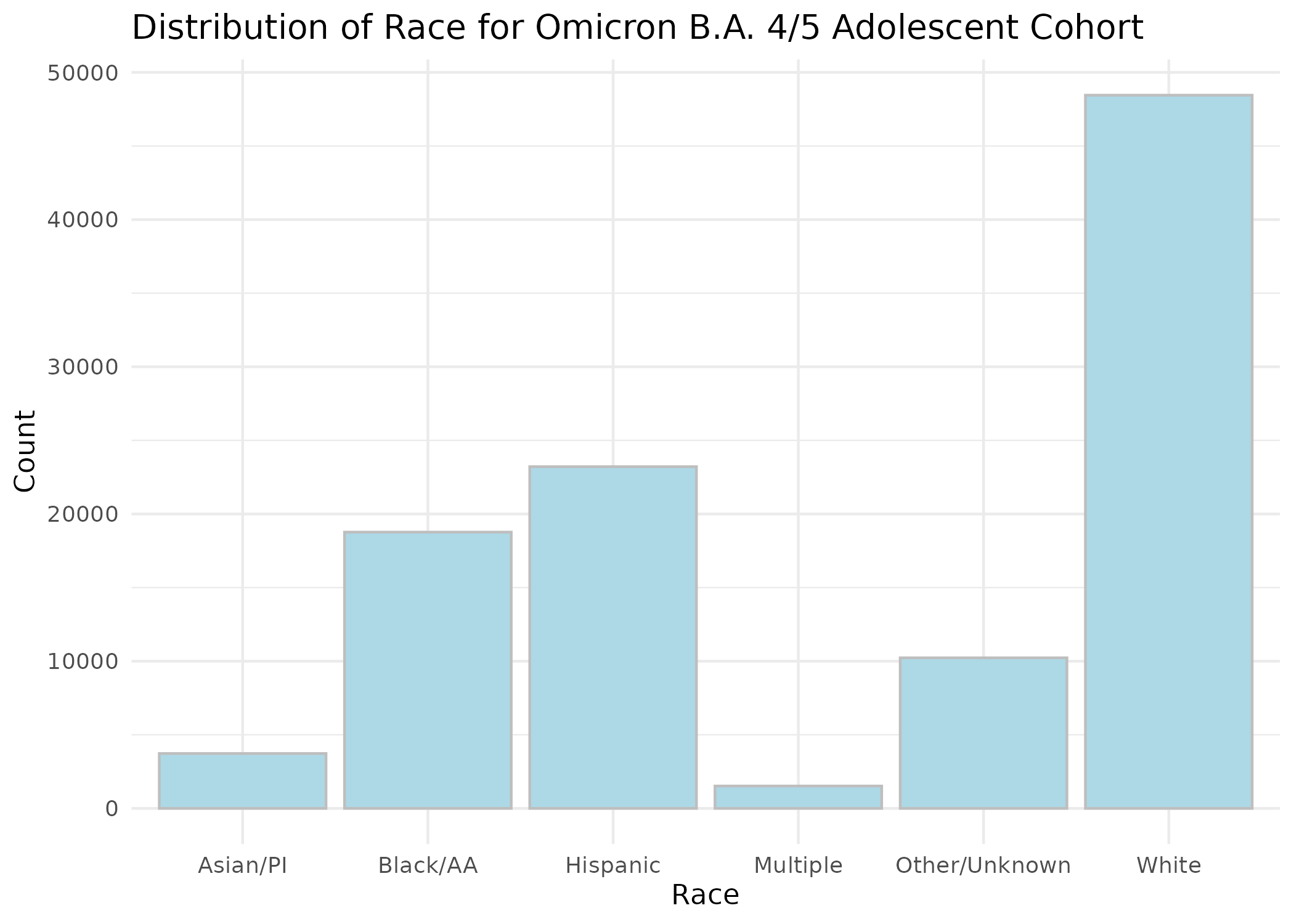

**Figure S5: Distribution of Race for Omicron XBB or later Children Cohort.**
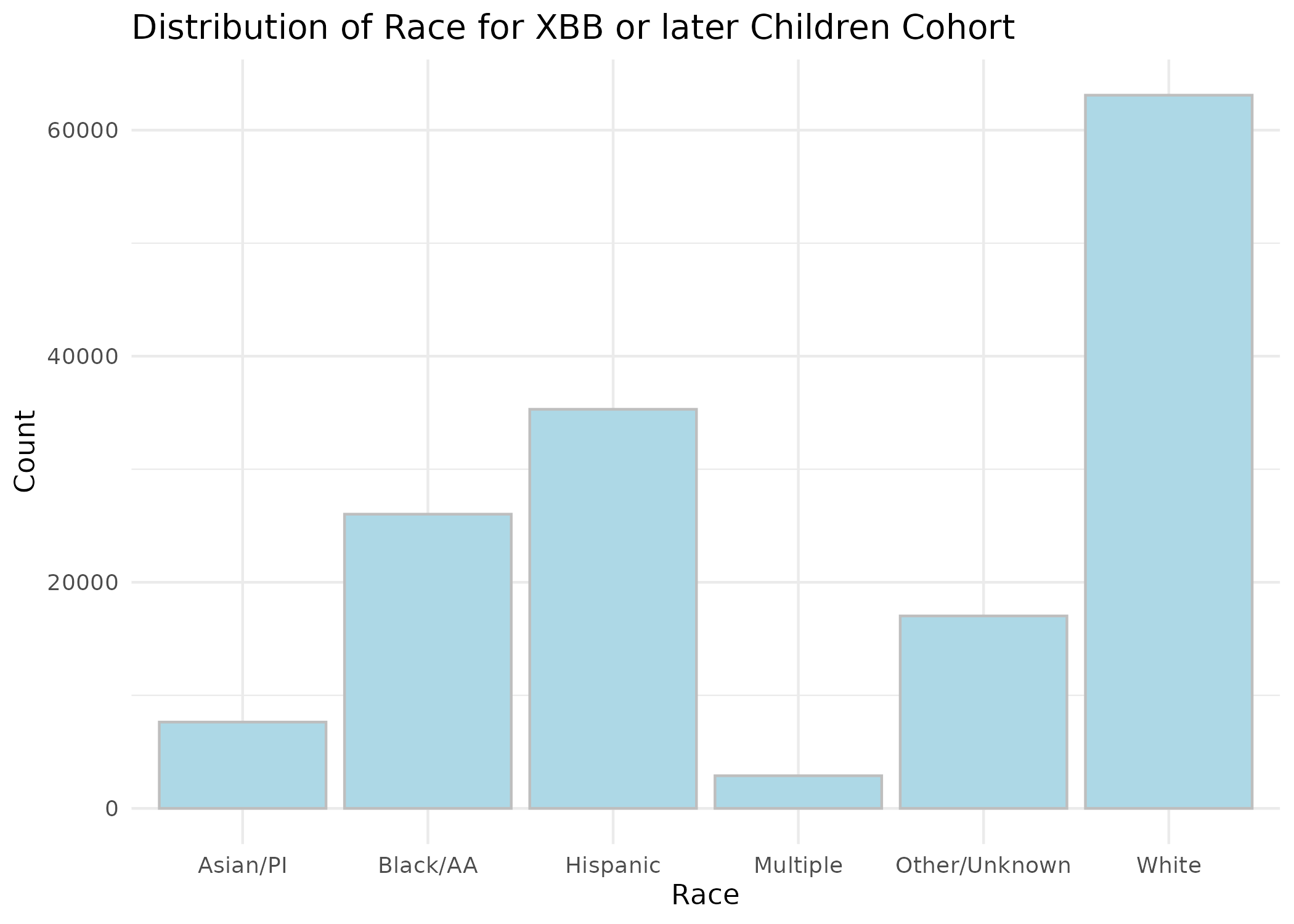

**Figure S6: Distribution of Race for Omicron XBB or later Adolescent Cohort.**
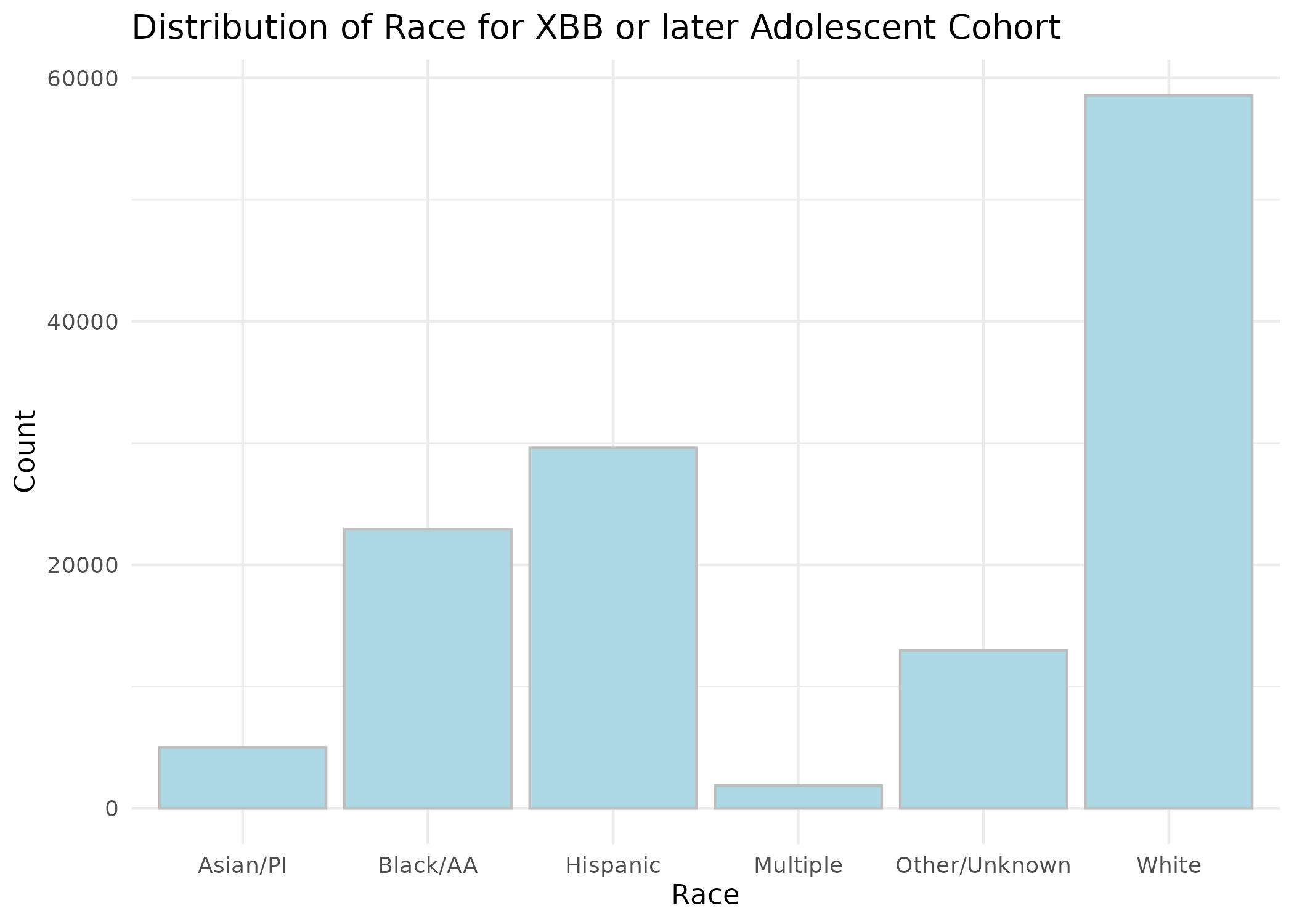

**Figure S7: Distribution of Prior Vaccine Doses for Omicron B.A. 1/2 Children Cohort.**
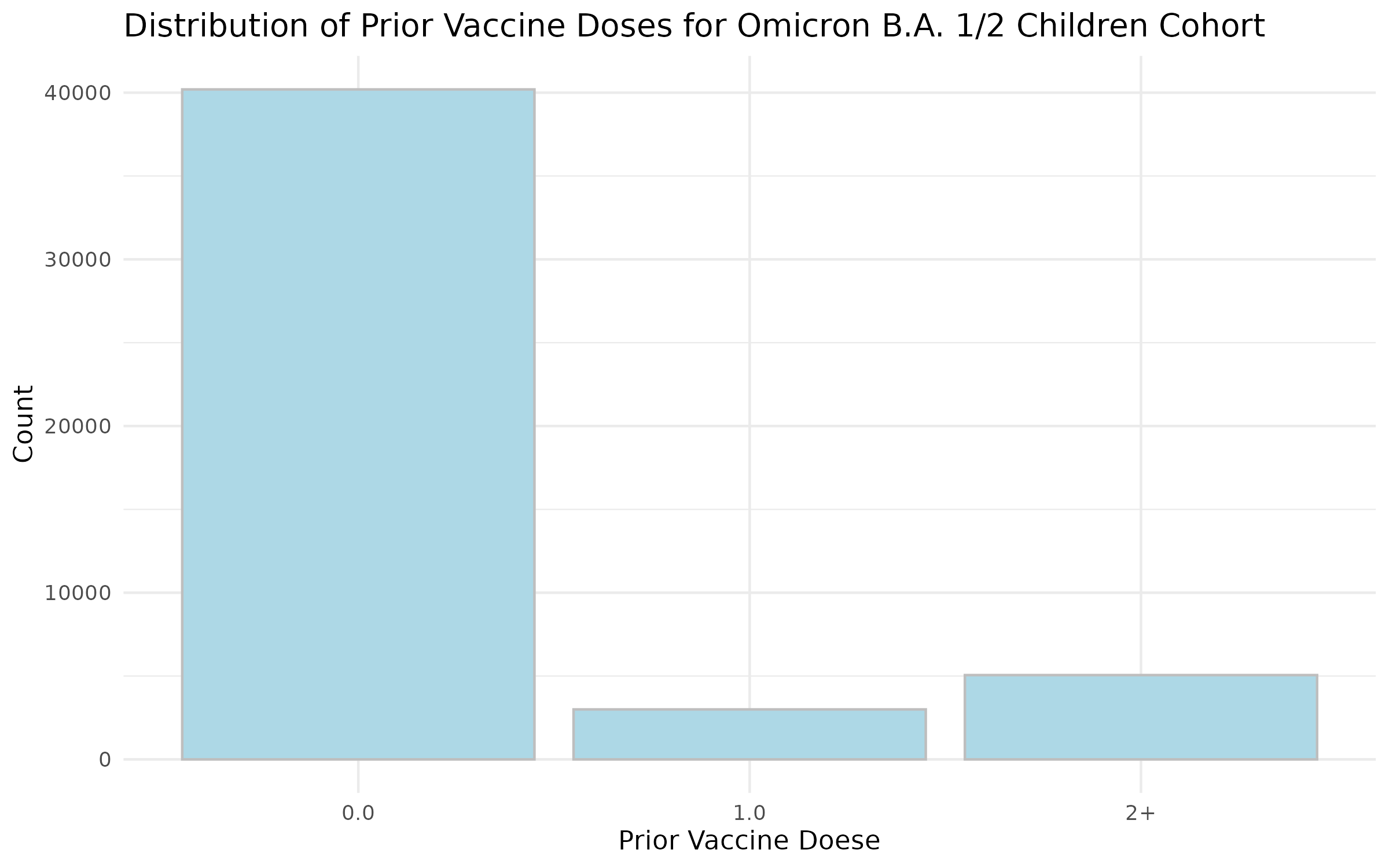

**Figure S8: Distribution of Prior Vaccine Doses for Omicron B.A. 1/2 Adolescent Cohort.**
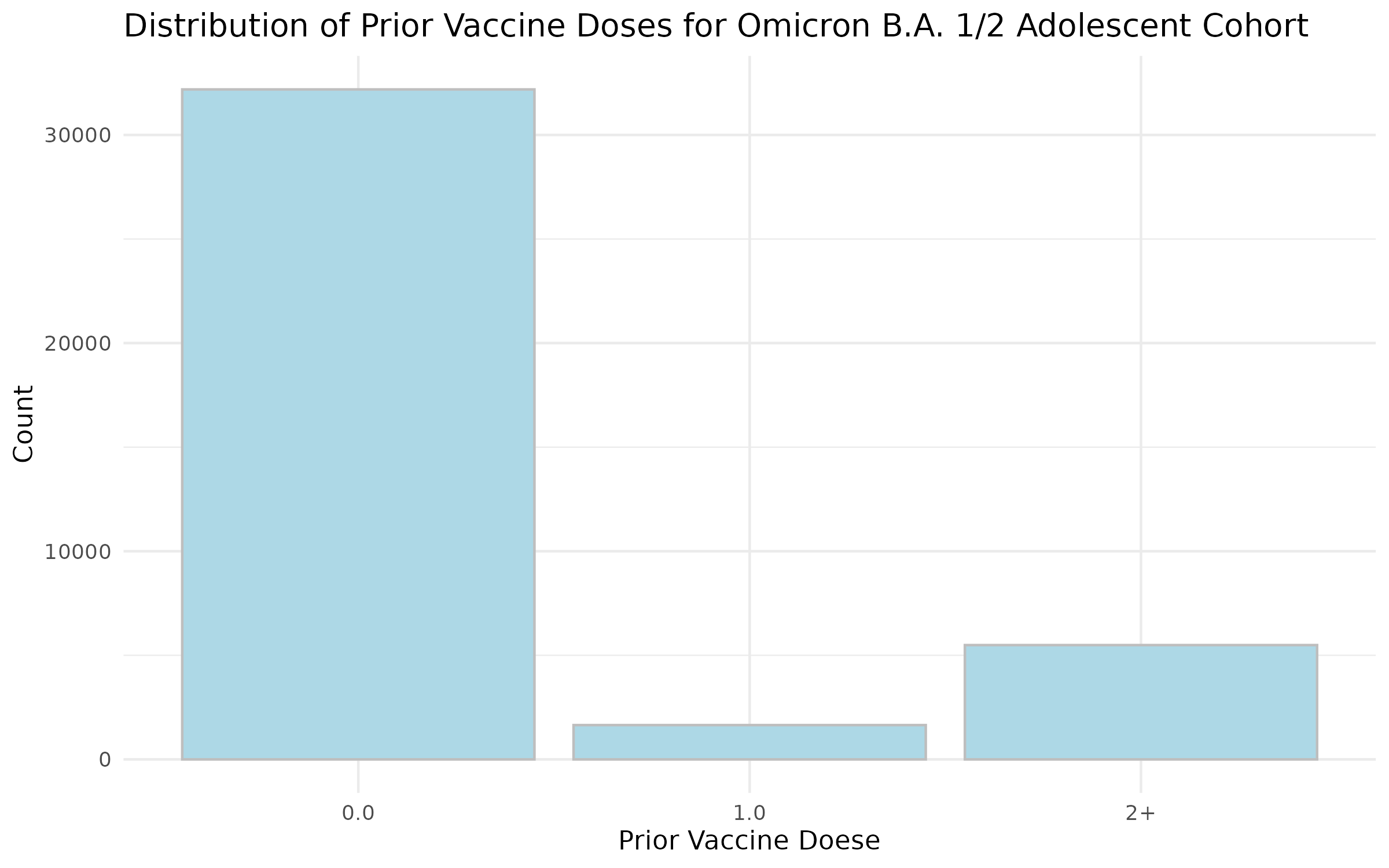

**Figure S9: Distribution of Prior Vaccine Doses for Omicron B.A. 4/5 Children Cohort.**
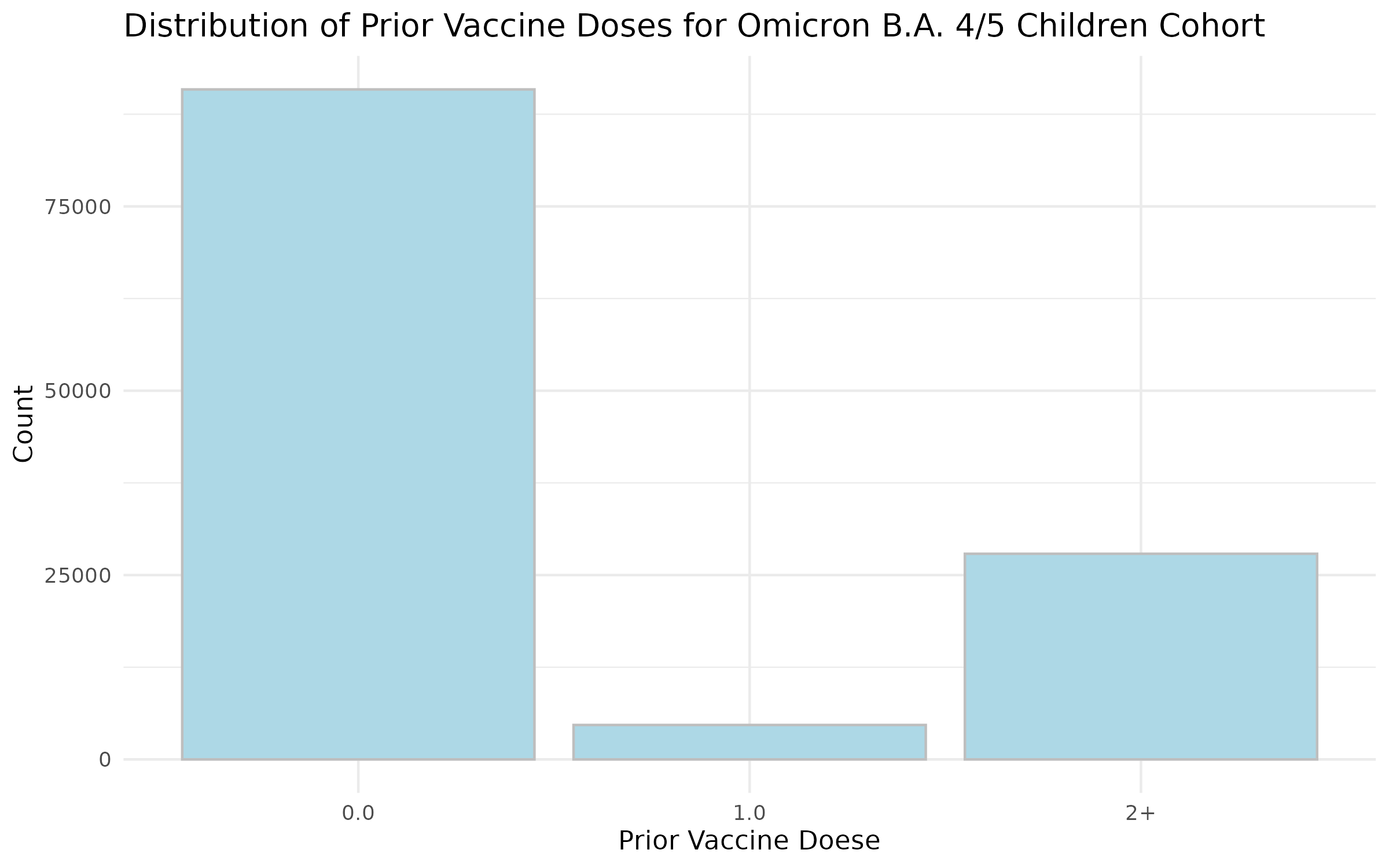

**Figure S10: Distribution of Prior Vaccine Doses for Omicron B.A. 4/5 Adolescent Cohort.**
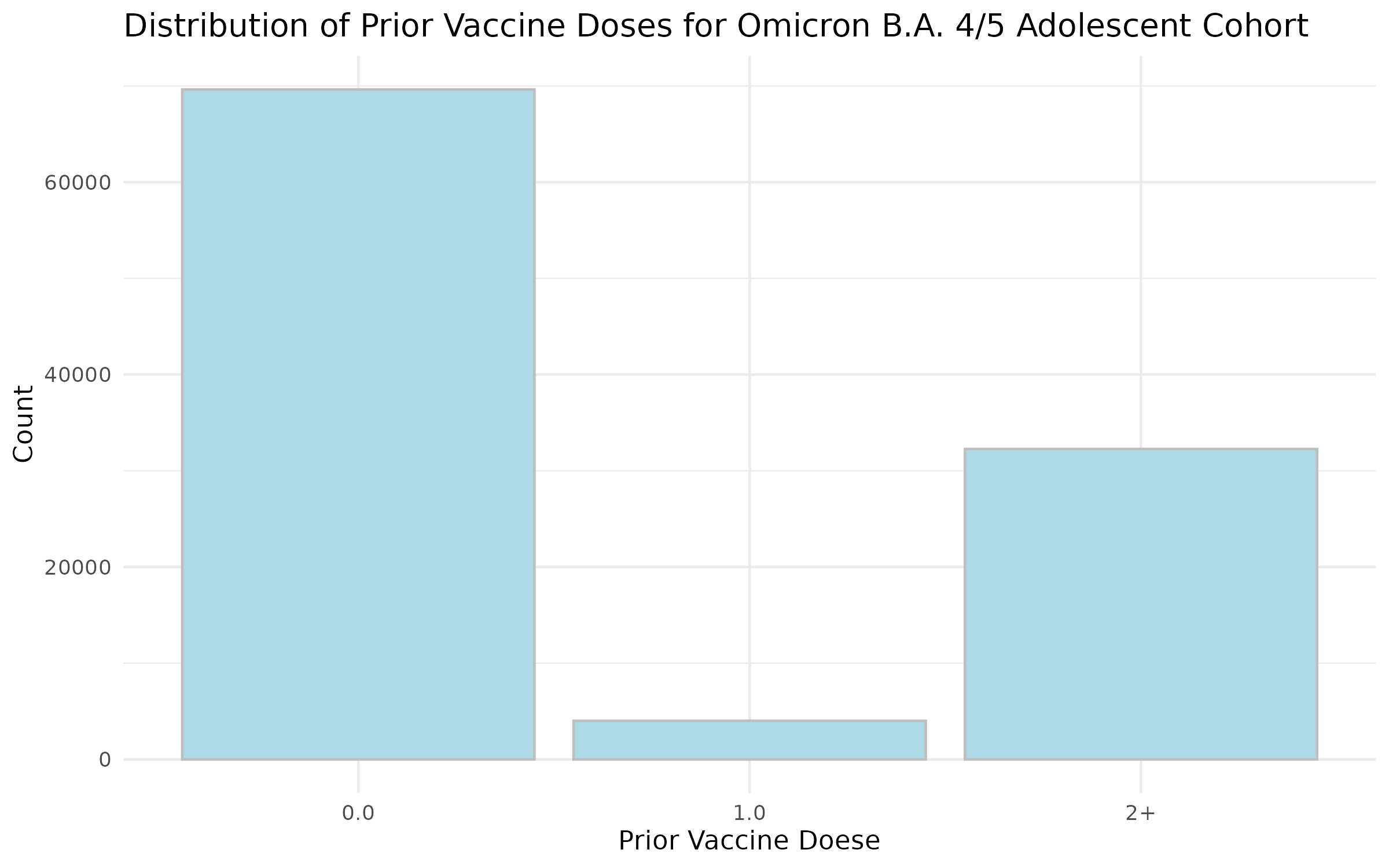

**Figure S11: Distribution of Prior Vaccine Doses for Omicron XBB or later Children Cohort.**
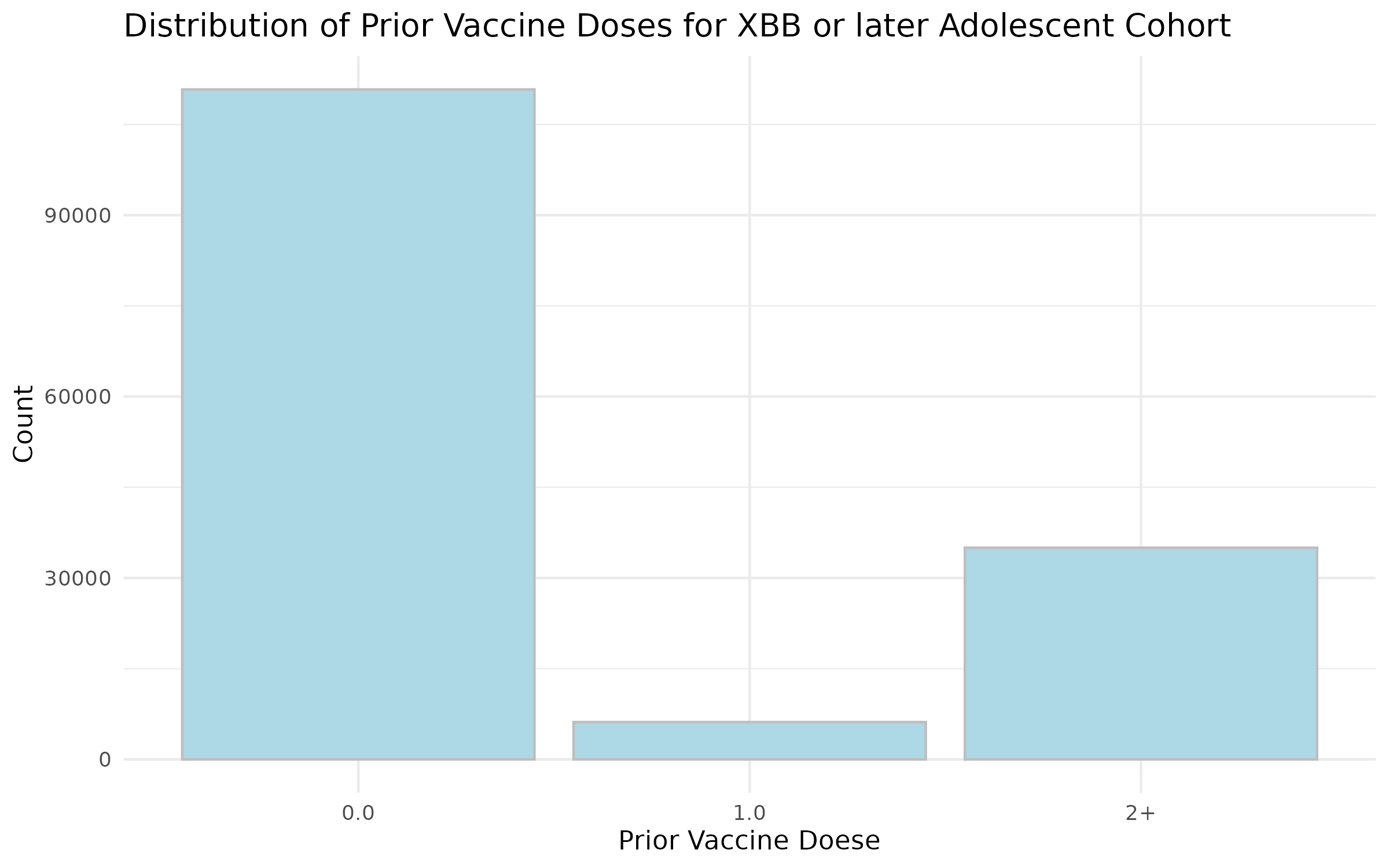

**Figure S12: Distribution of Prior Vaccine Doses for Omicron XBB or later Adolescent Cohort.**
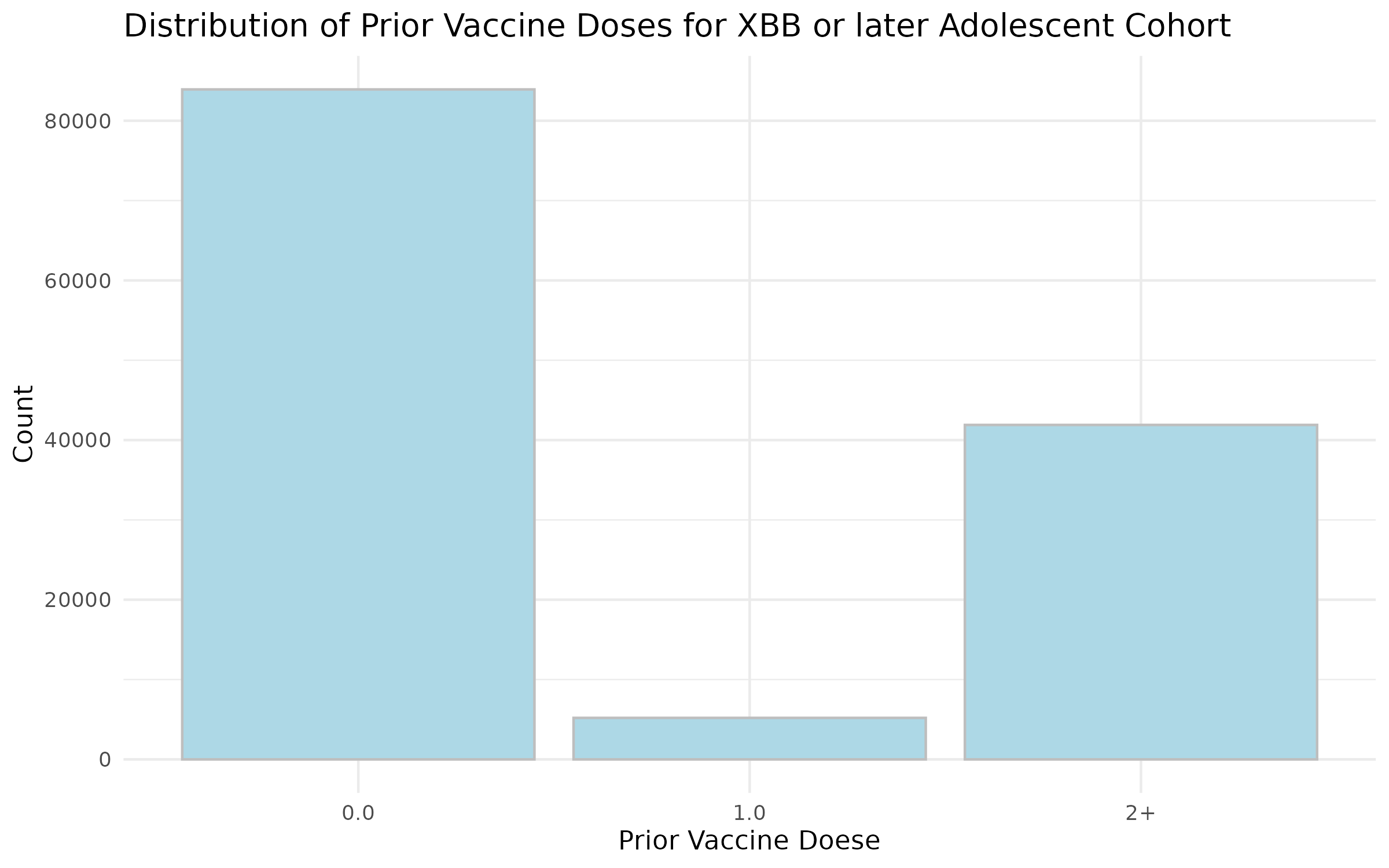

**Figure S13: Distribution of Time since the Last Vaccine dose before the Study Start Date for Omicron B.A. 1/2 Children Cohort.**
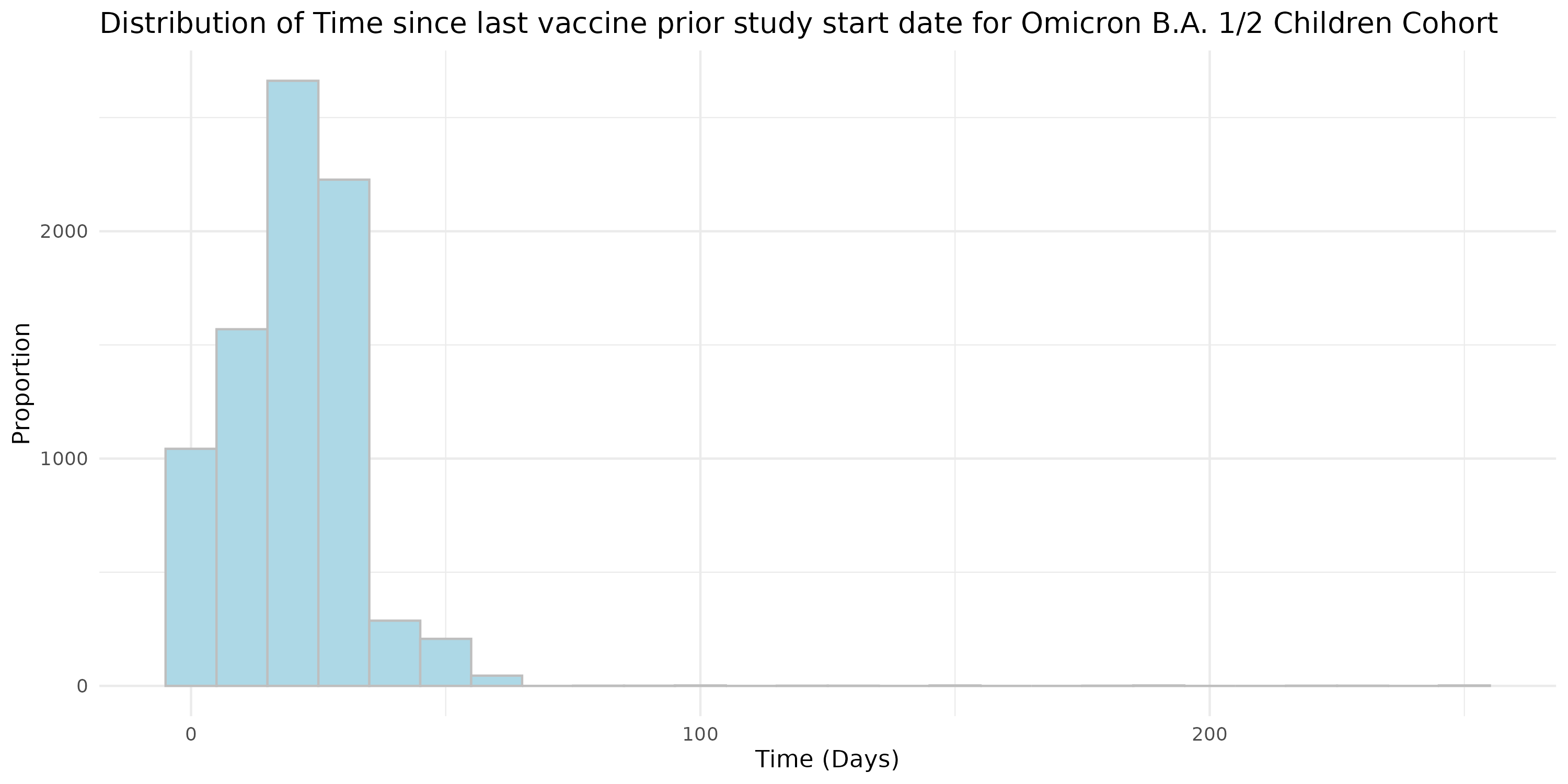

**Figure S14: Distribution of Time since the Last Vaccine dose before the Study Start Date for Omicron B.A. 1/2 Adolescent Cohort.**
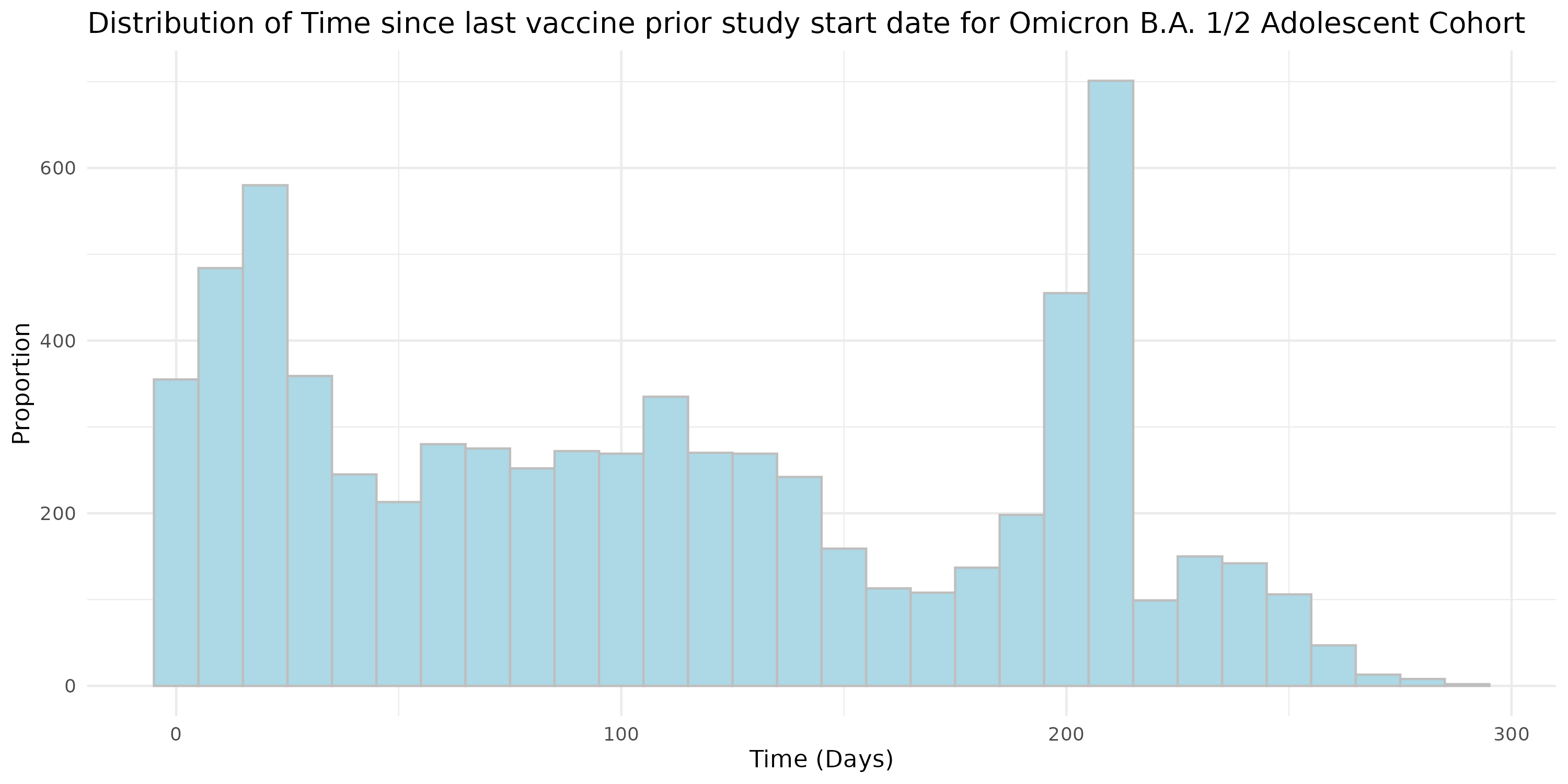

**Figure S15: Distribution of Time since the Last Vaccine dose before the Study Start Date for Omicron B.A. 4/5 Children Cohort.**
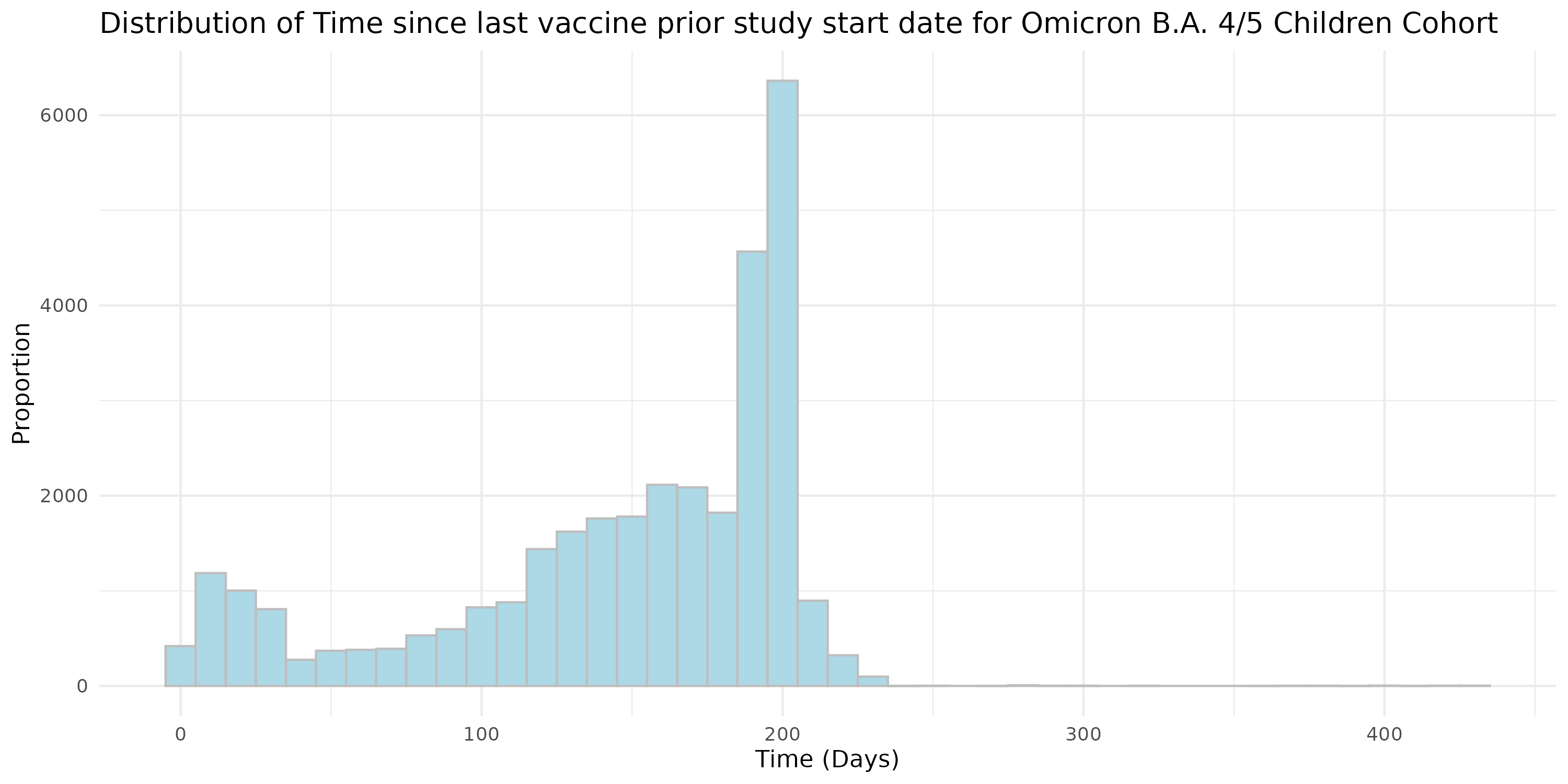

**Figure S16: Distribution of Time since the Last Vaccine dose before the Study Start Date for Omicron B.A. 4/5 Adolescent Cohort.**
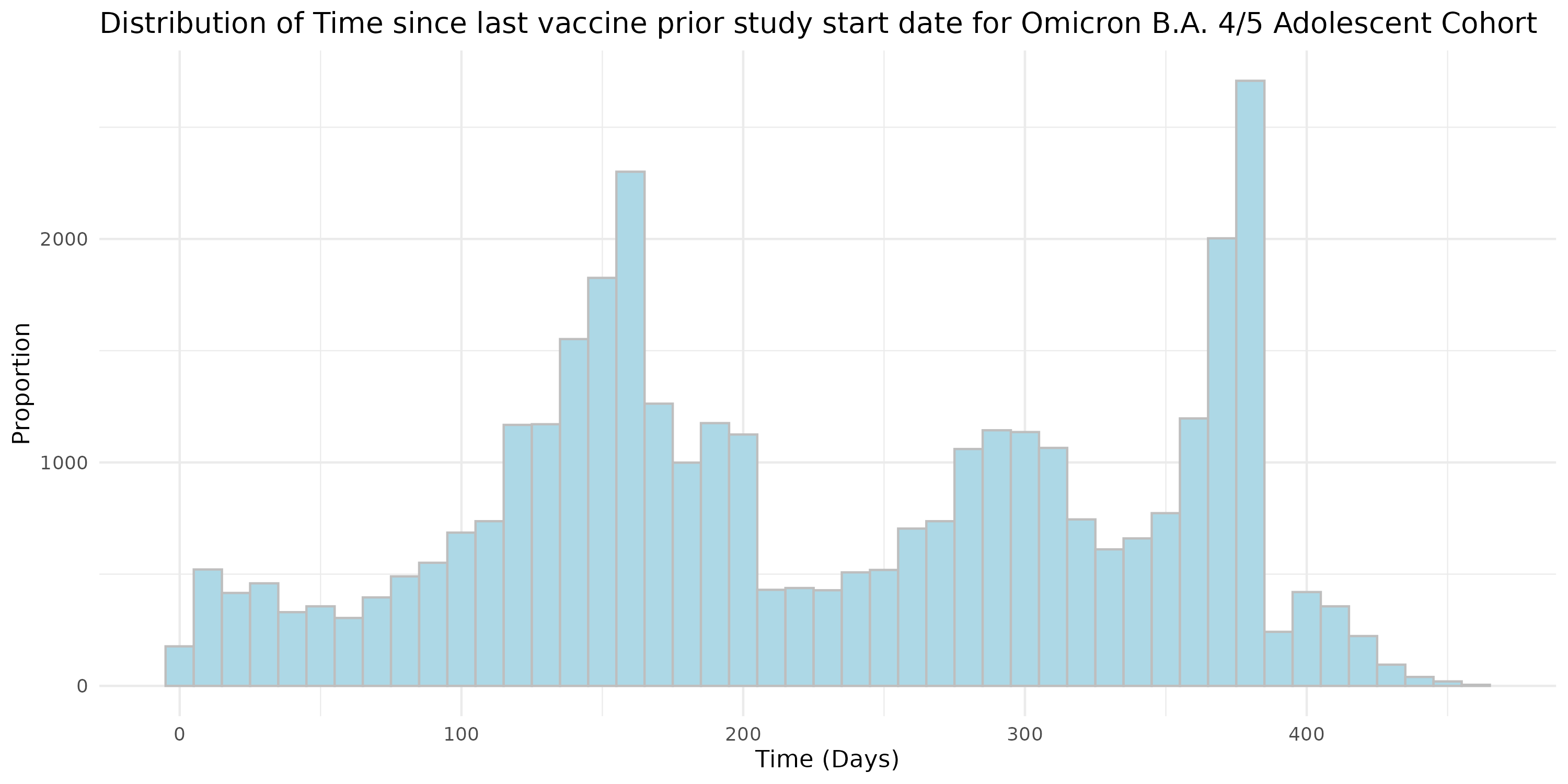

**Figure S17: Distribution of Time since the Last Vaccine dose before the Study Start Date for Omicron XBB or later Children Cohort.**
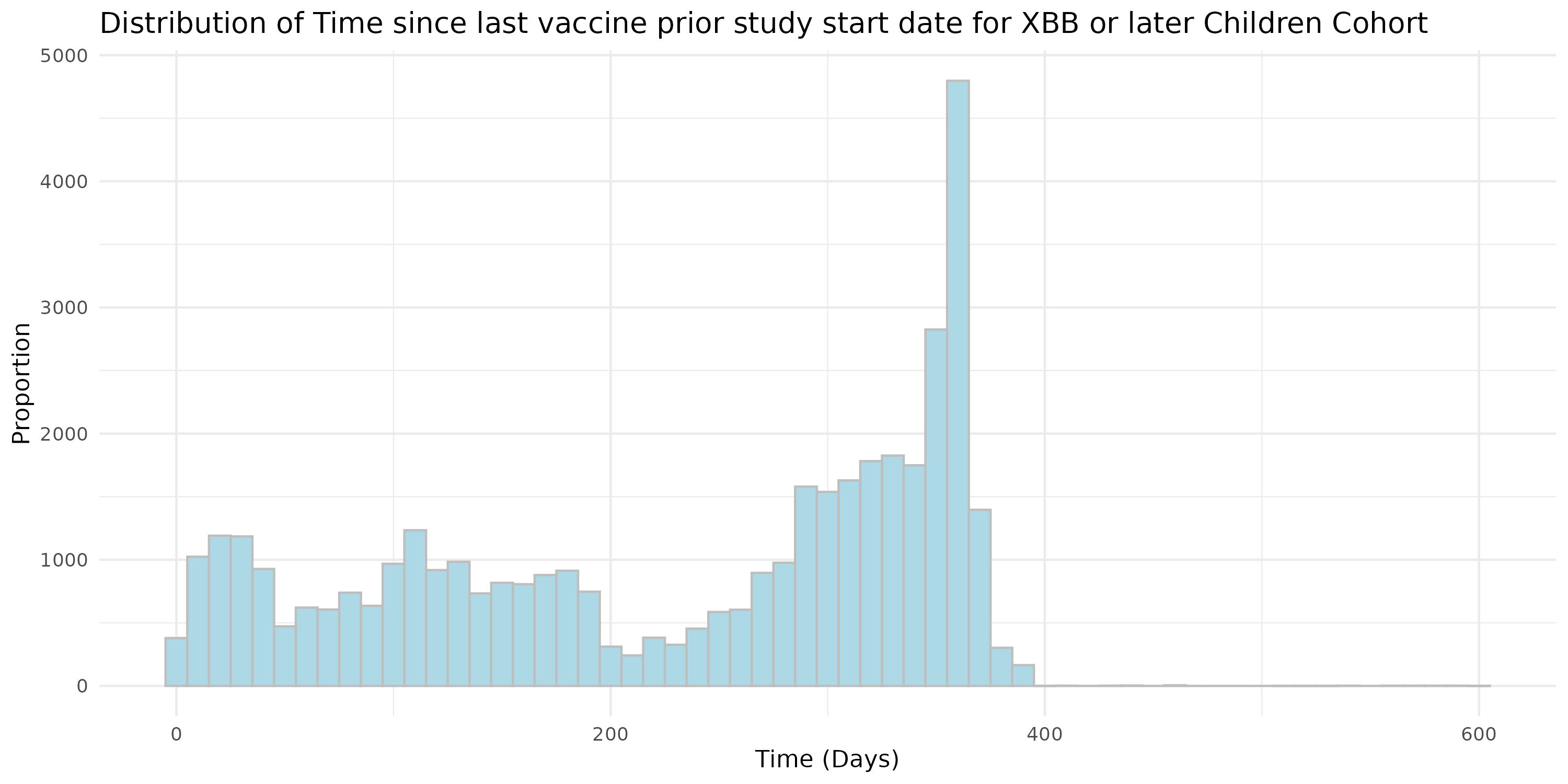

**Figure S18: Distribution of Time since the Last Vaccine dose before the Study Start Date for Omicron XBB or later Adolescent Cohort.**
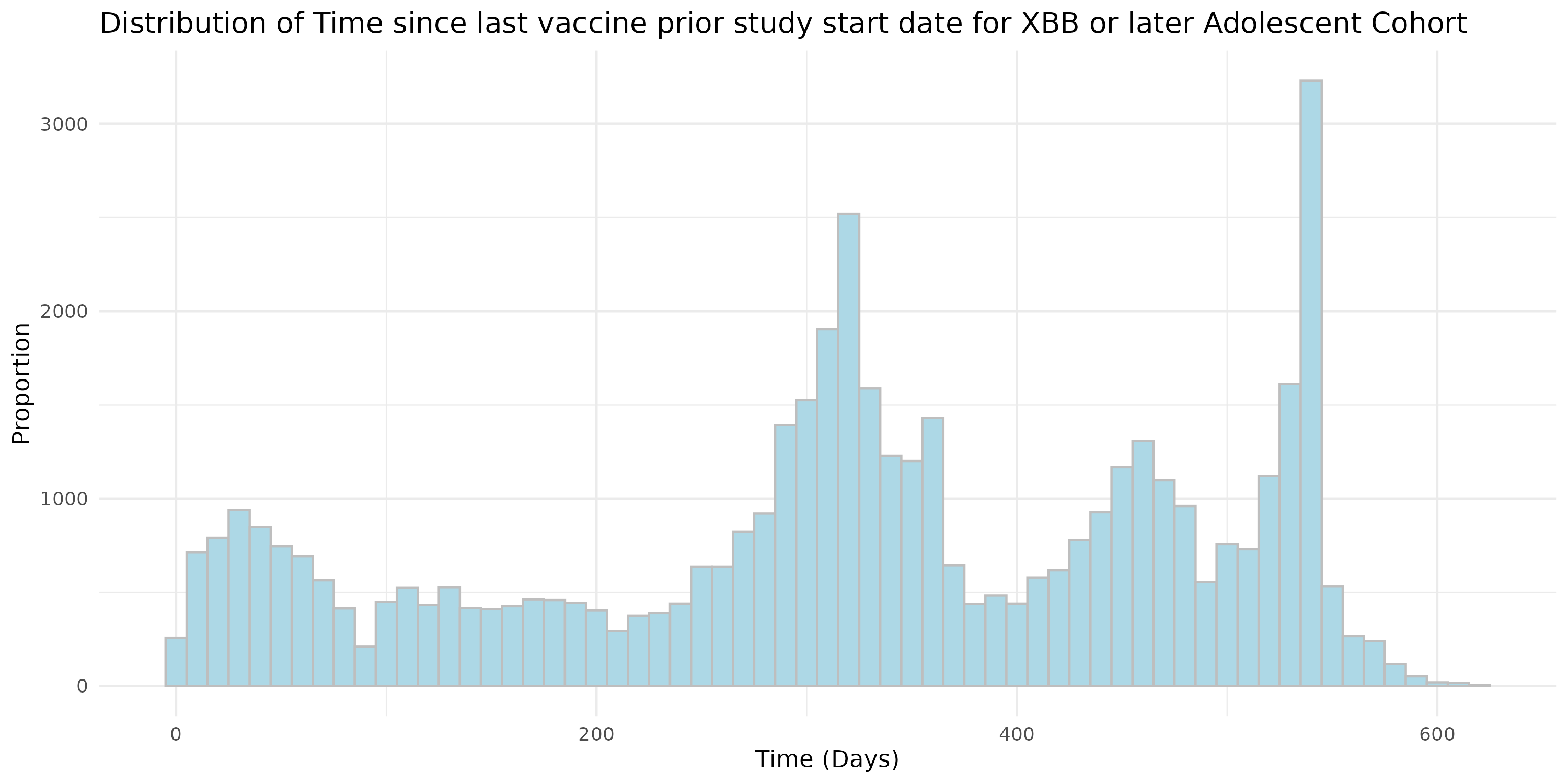

**Figure S19: Distribution of Vaccine Brand for Vaccinated Persons in each cohort.**
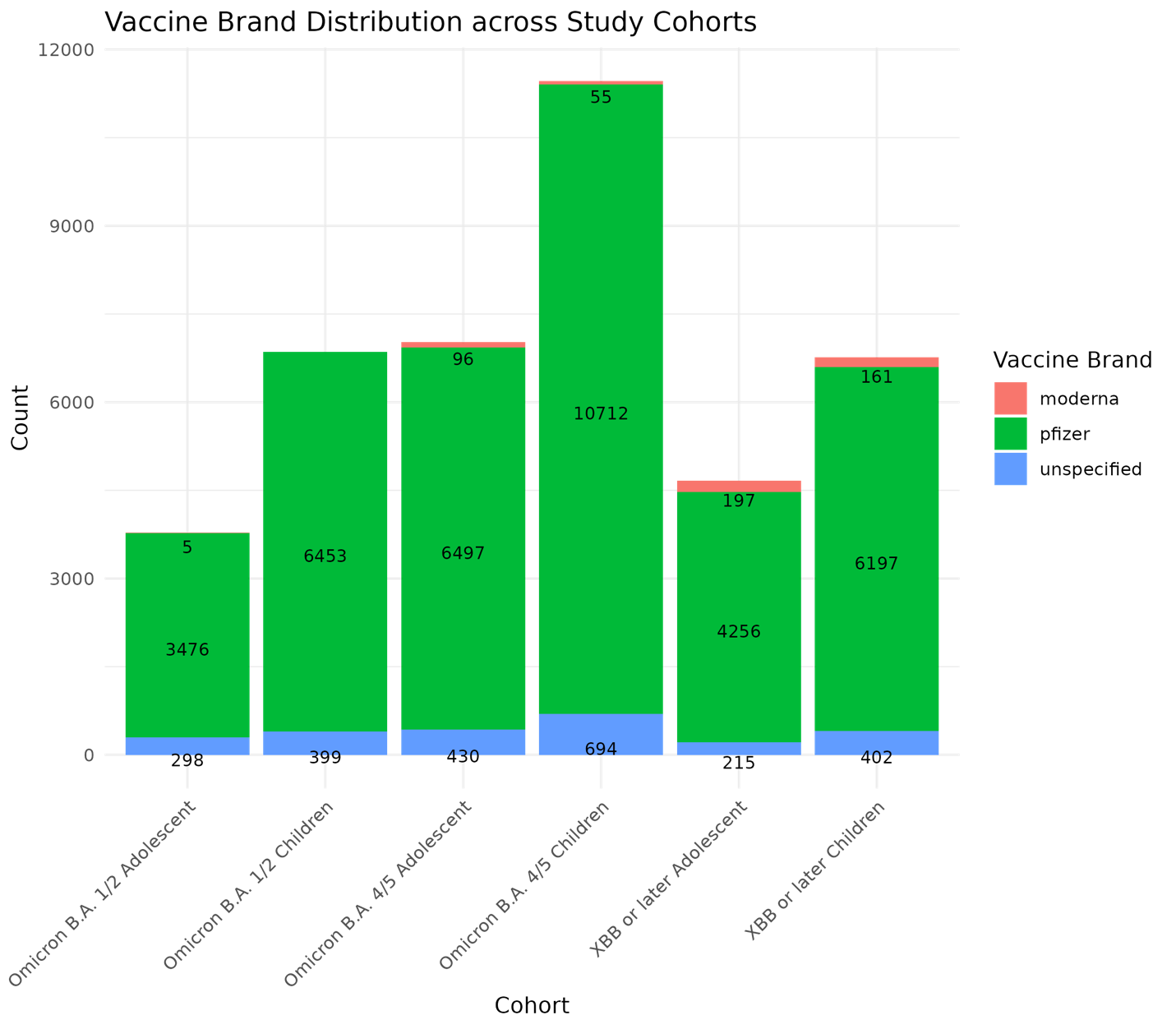

#### E. Patient characteristic balance

We evaluate the balance of patient characteristics using the standardized difference of means (SMD). Figures S1-S6 presents the SMD of six study cohorts after employing propensity score matching and exact matching for assigning vaccinated and unvaccinated group.

**Figure S20: Patient characteristic balance for Omicron B.A. 1/2 Children Cohort after matching.**
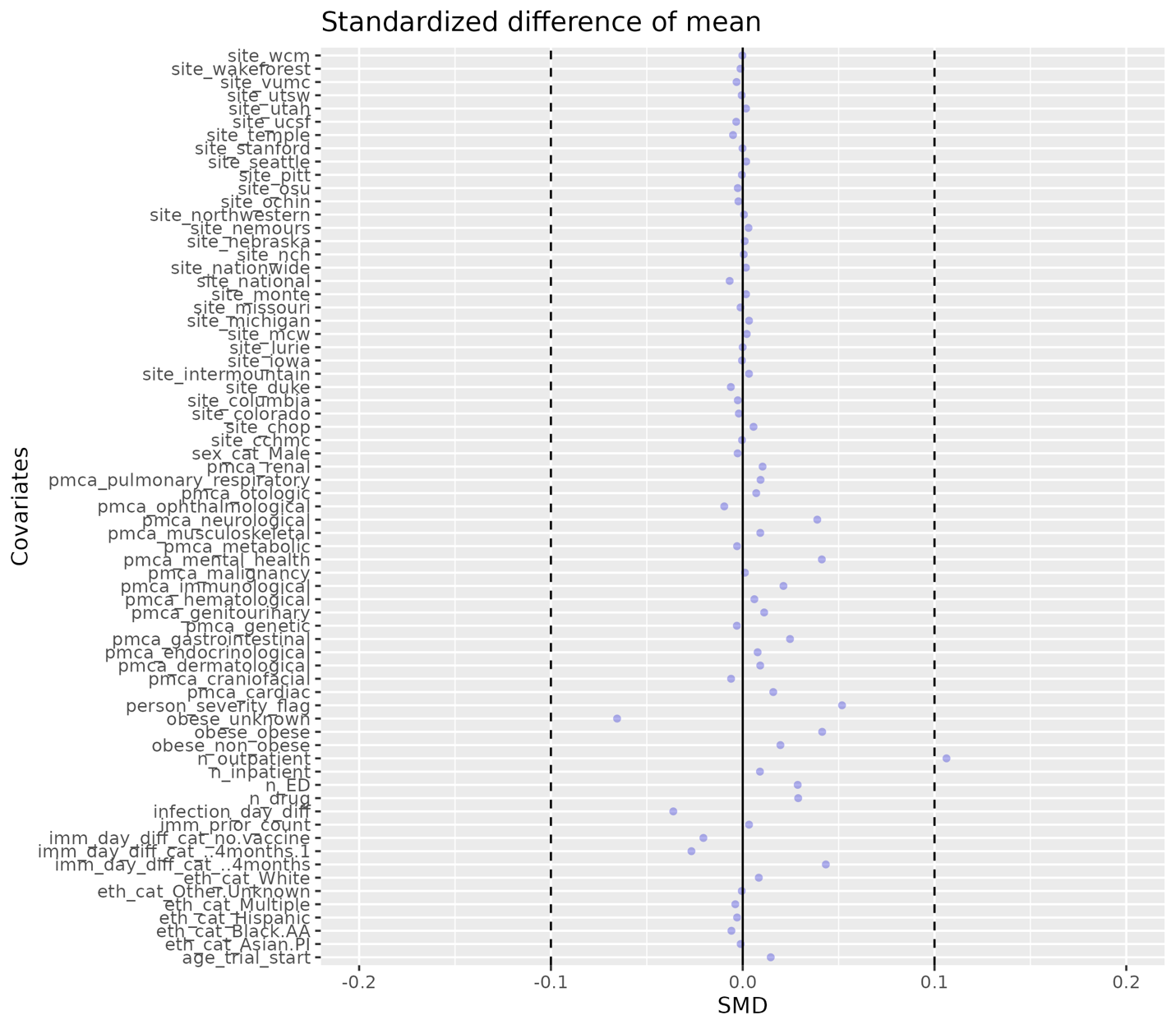

**Figure S21: Patient characteristic balance for Omicron B.A. 1/2 Adolescents Cohort after matching.**
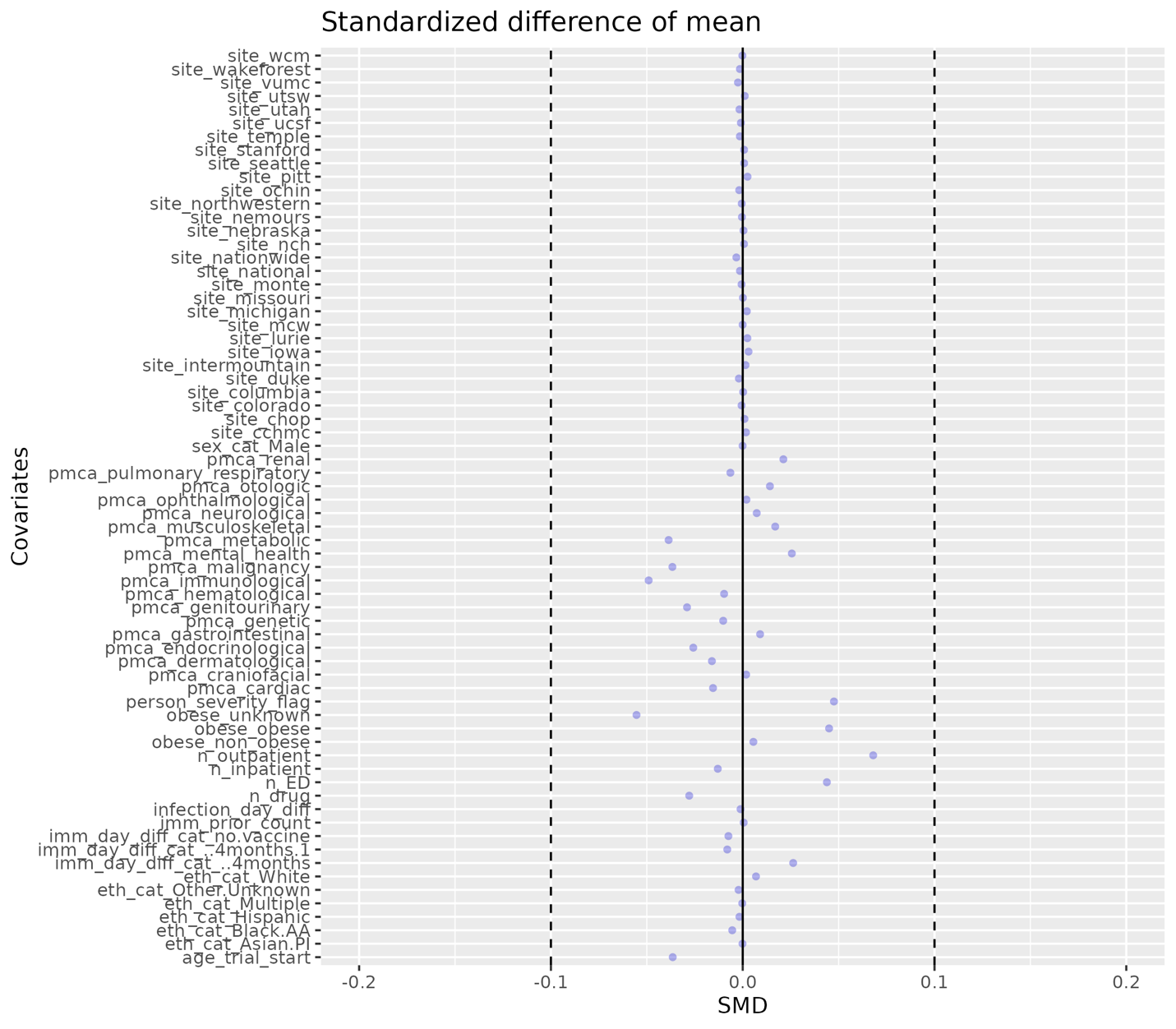

**Figure S22: Patient characteristic balance for Omicron B.A. 4/5 Children Cohort after matching.**
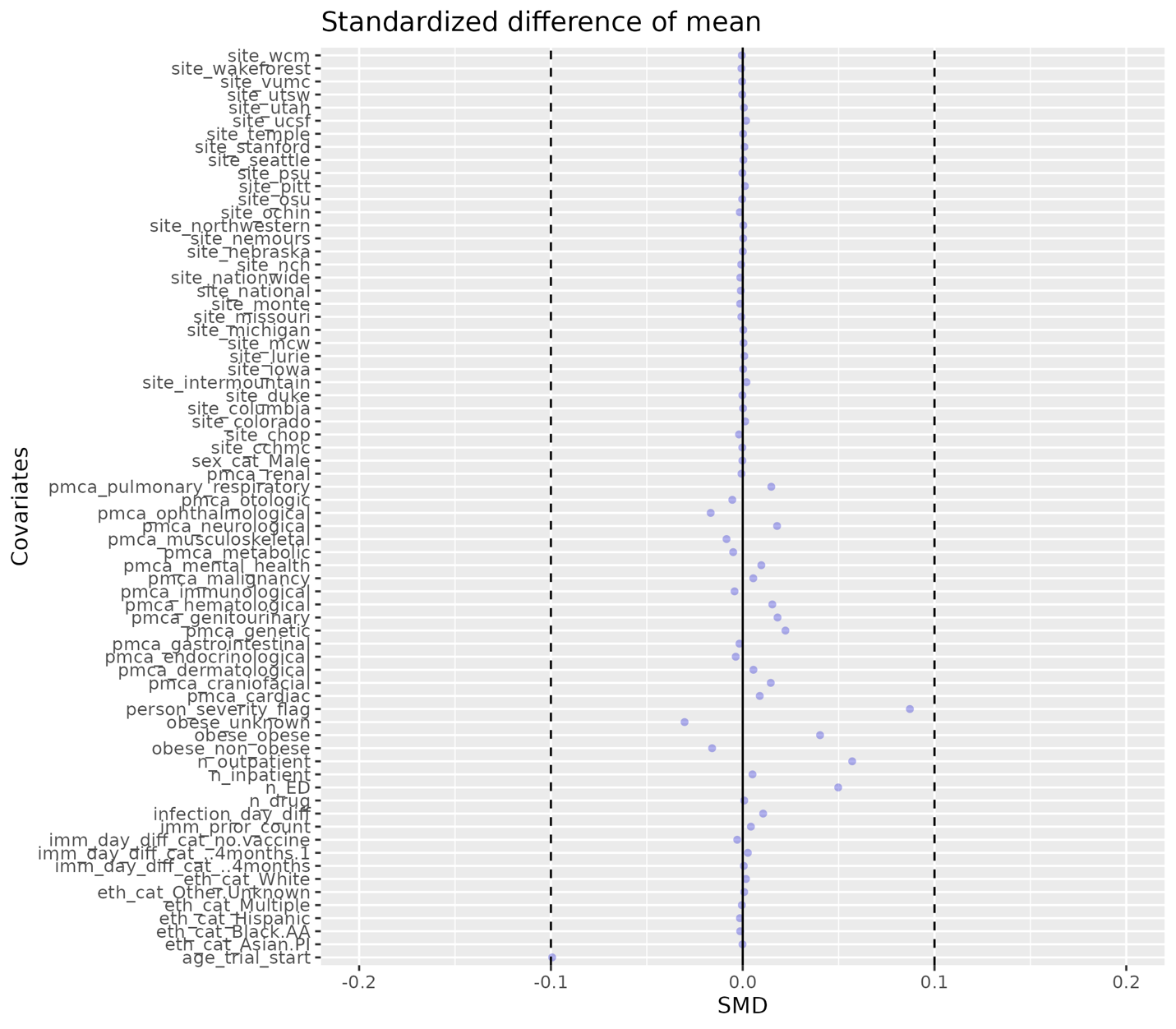

**Figure S23: Patient characteristic balance for Omicron B.A. 4/5 Adolescents Cohort after matching.**
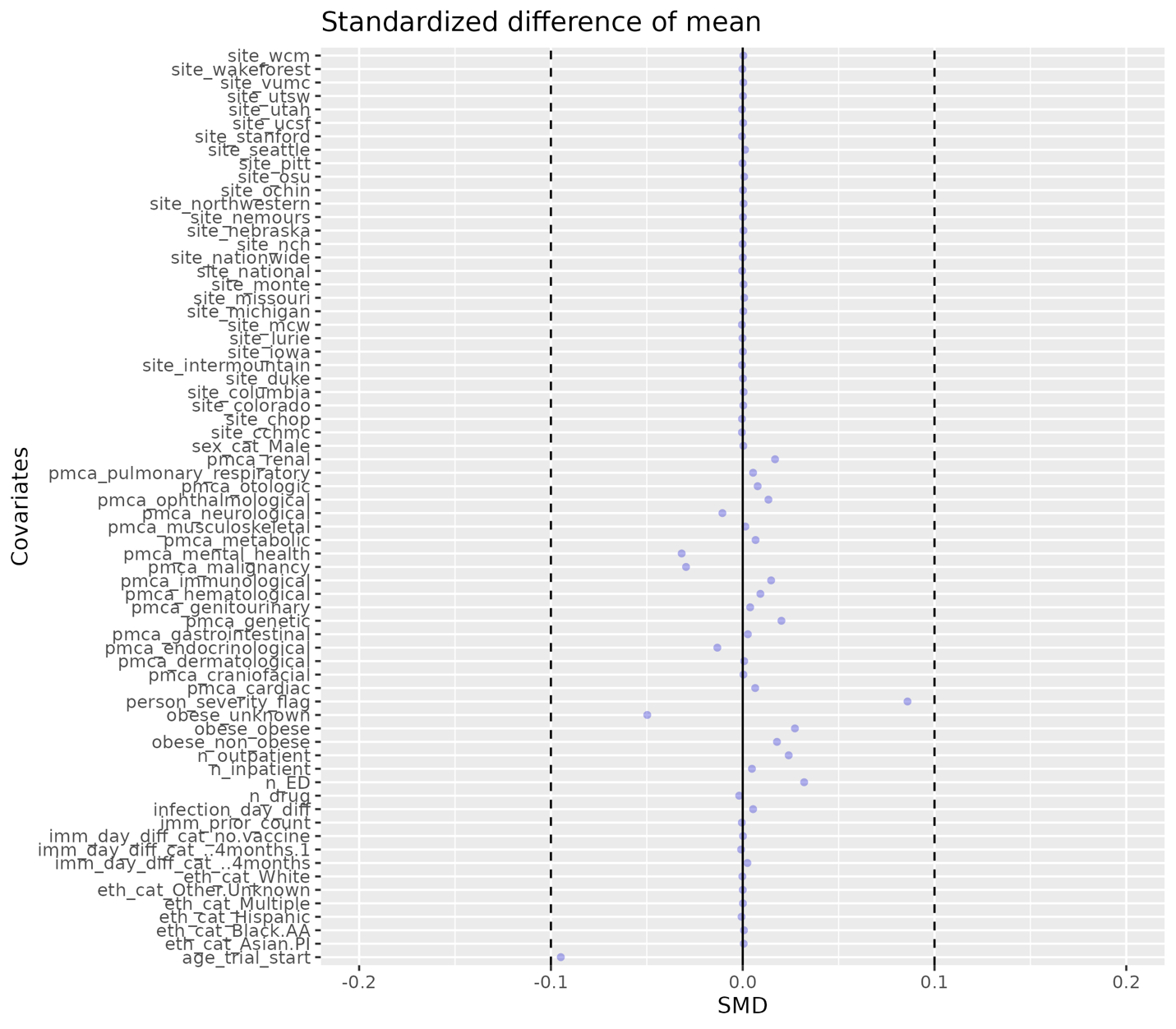

**Figure S24: Patient characteristic balance for Omicron XBB or later Children Cohort after matching.**
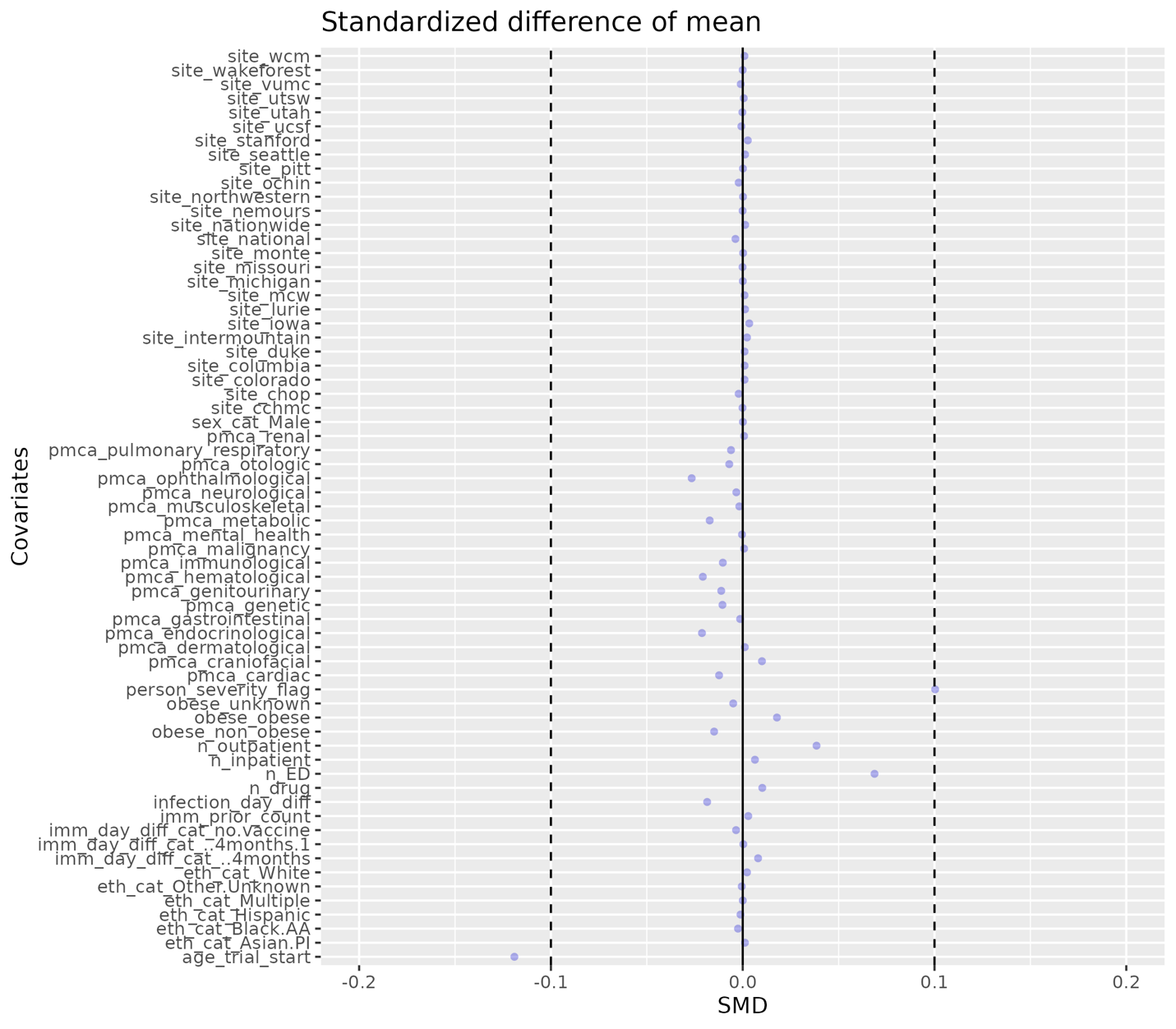

**Figure S25: Patient characteristic balance for Omicron XBB or later Adolescents Cohort after matching.**
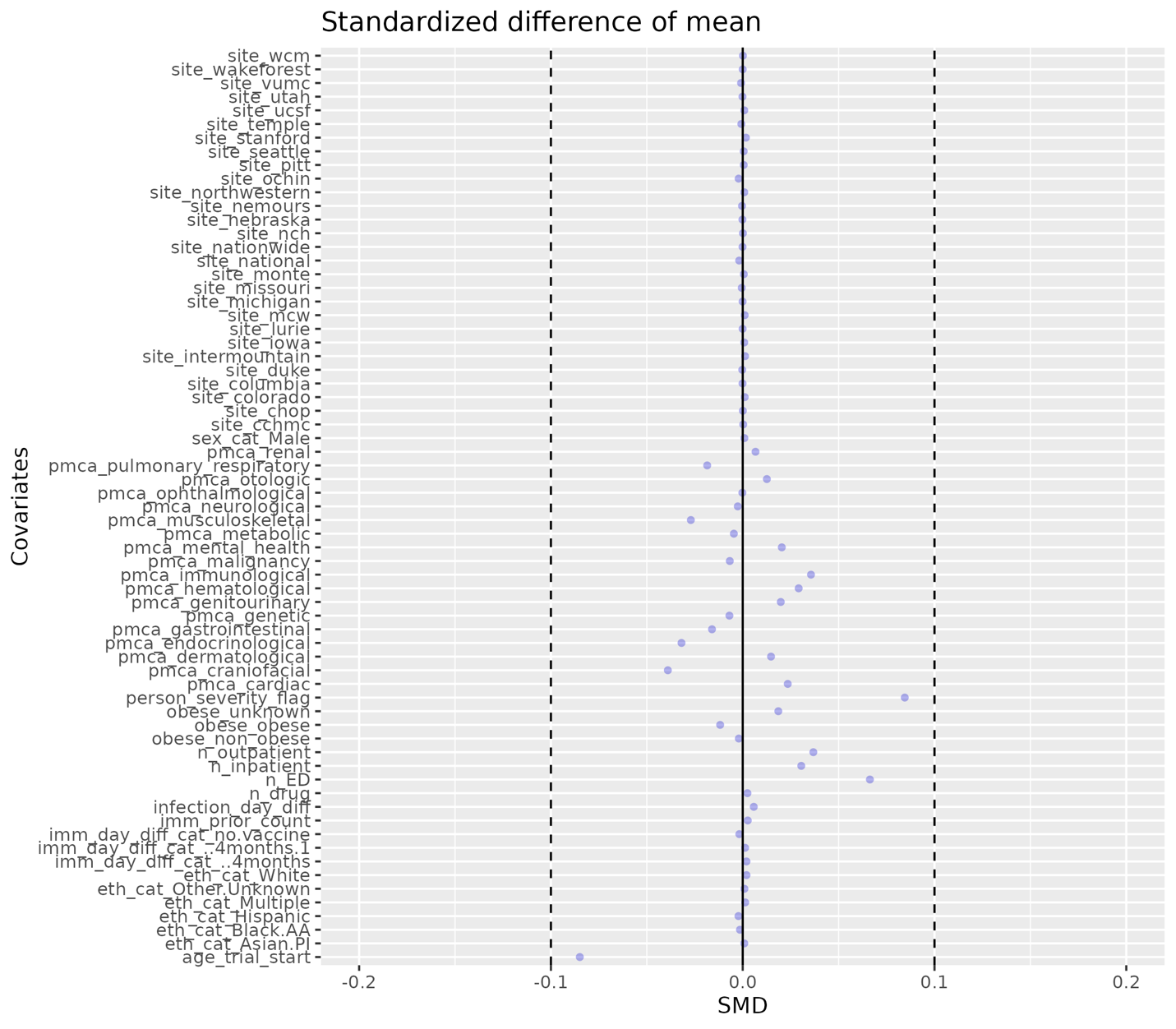

#### F. Distribution of the time since the last COVID-19 immunization prior to trial start date after matching

As an extension of **Figures S13–S18**, we present the distribution of time since the last vaccination prior to each corresponding trial start date for both vaccinated and unvaccinated individuals after matching (**Figures S26–S31**). The distributions exhibit similar patterns between the vaccinated and unvaccinated groups across all study cohorts, indicating successful balancing between the vaccinated and comparator groups. This balance helps mitigate potential bias in the primary trial emulation analysis results.

**Figure S26: Distribution of the time since the last COVID-19 immunization prior to Omicron B.A. 1/2 trial start date for vaccinated and unvaccinated individuals in Children Cohort.**
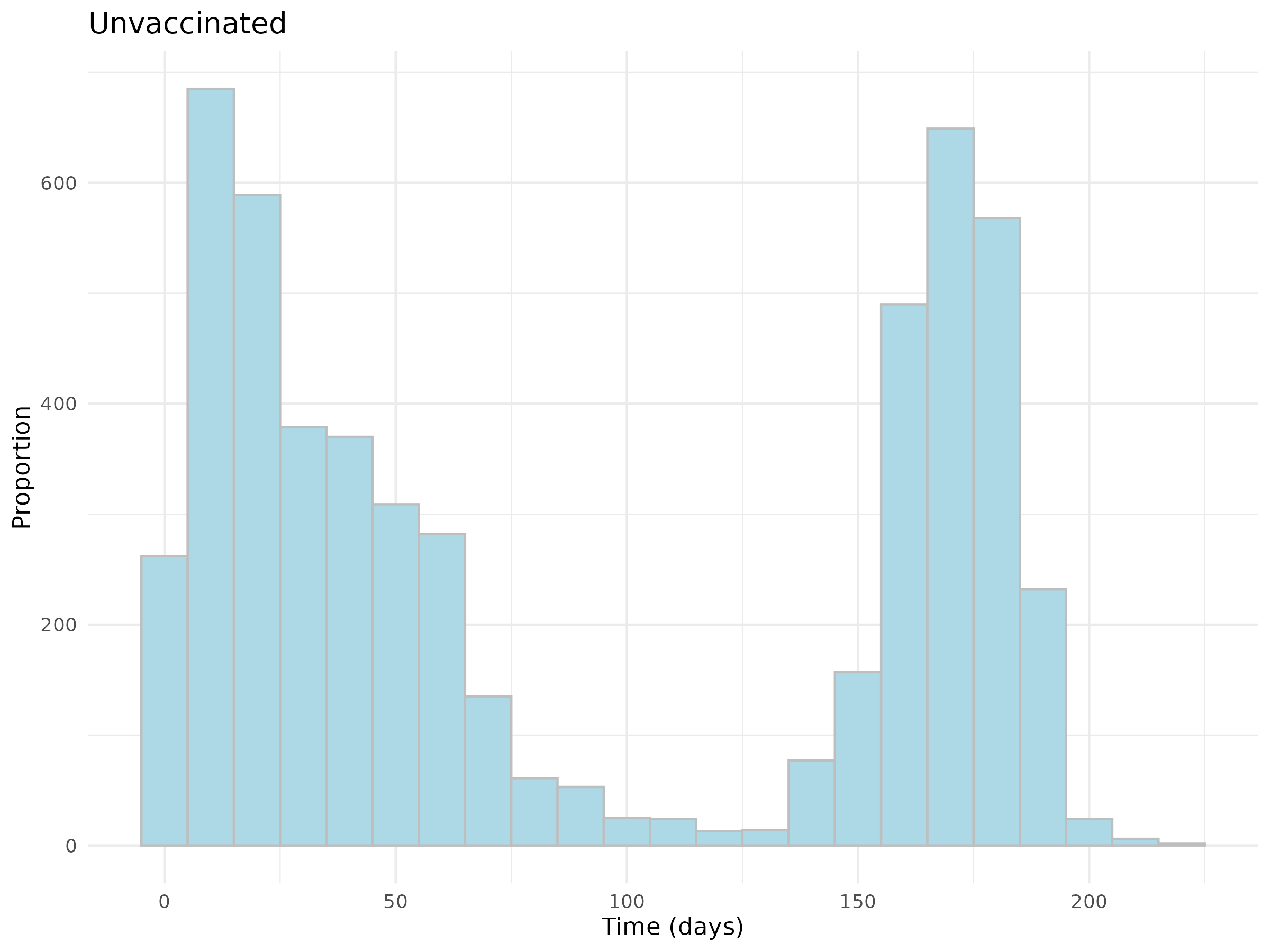

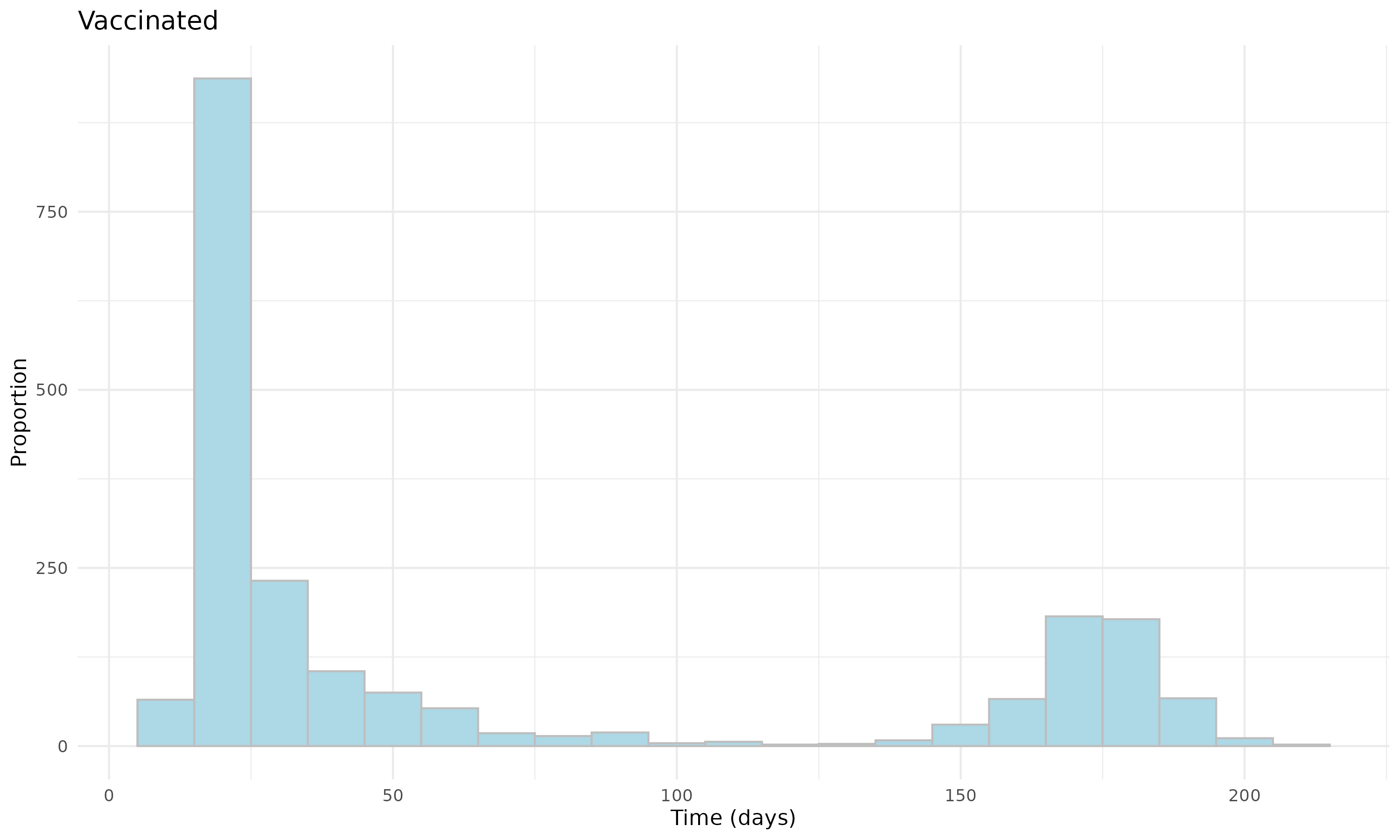

**Figure S27: Distribution of the time since the last COVID-19 immunization prior to Omicron B.A. 1/2 trial start date for vaccinated and unvaccinated individuals in Adolescent Cohort.**

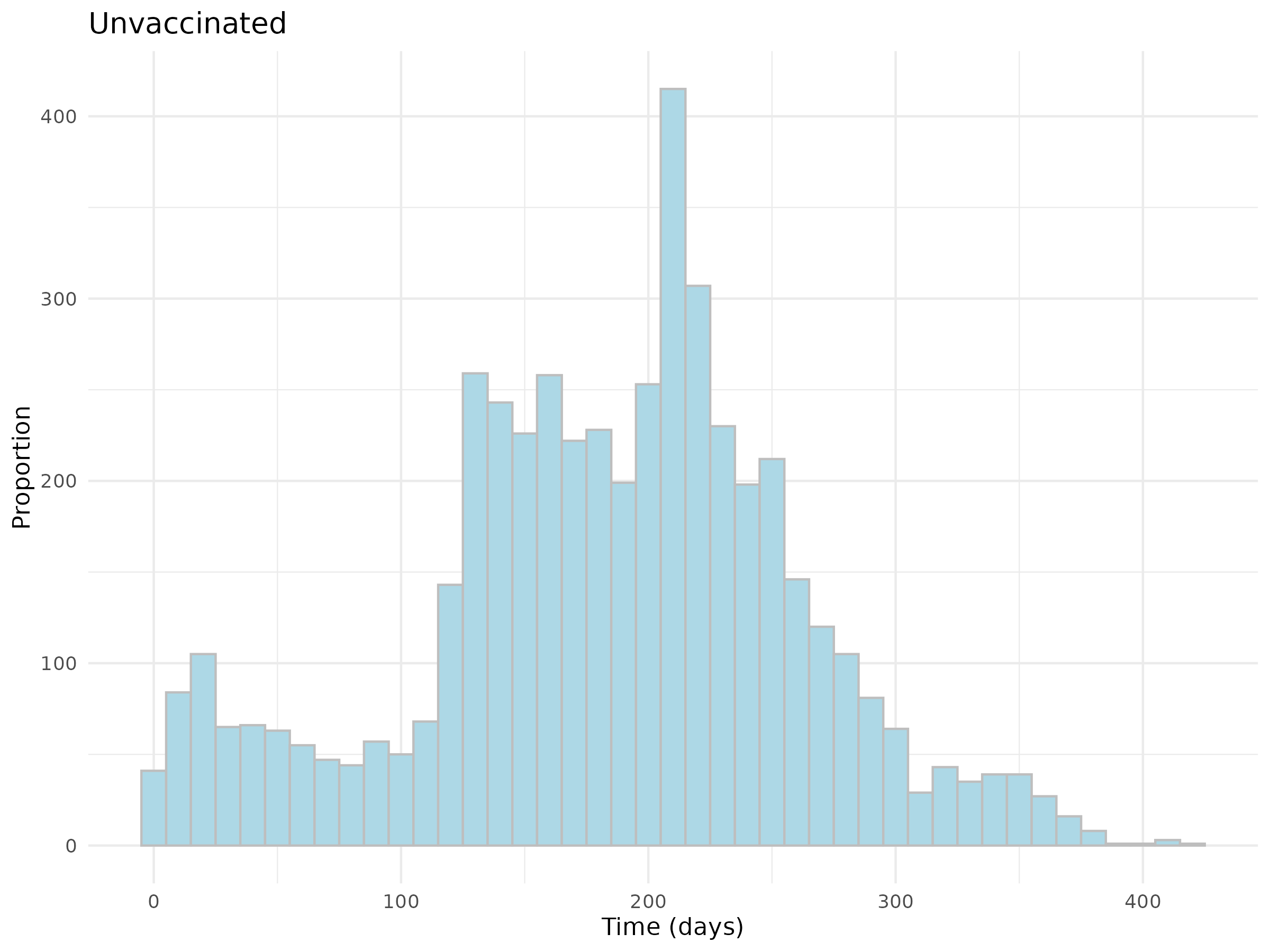

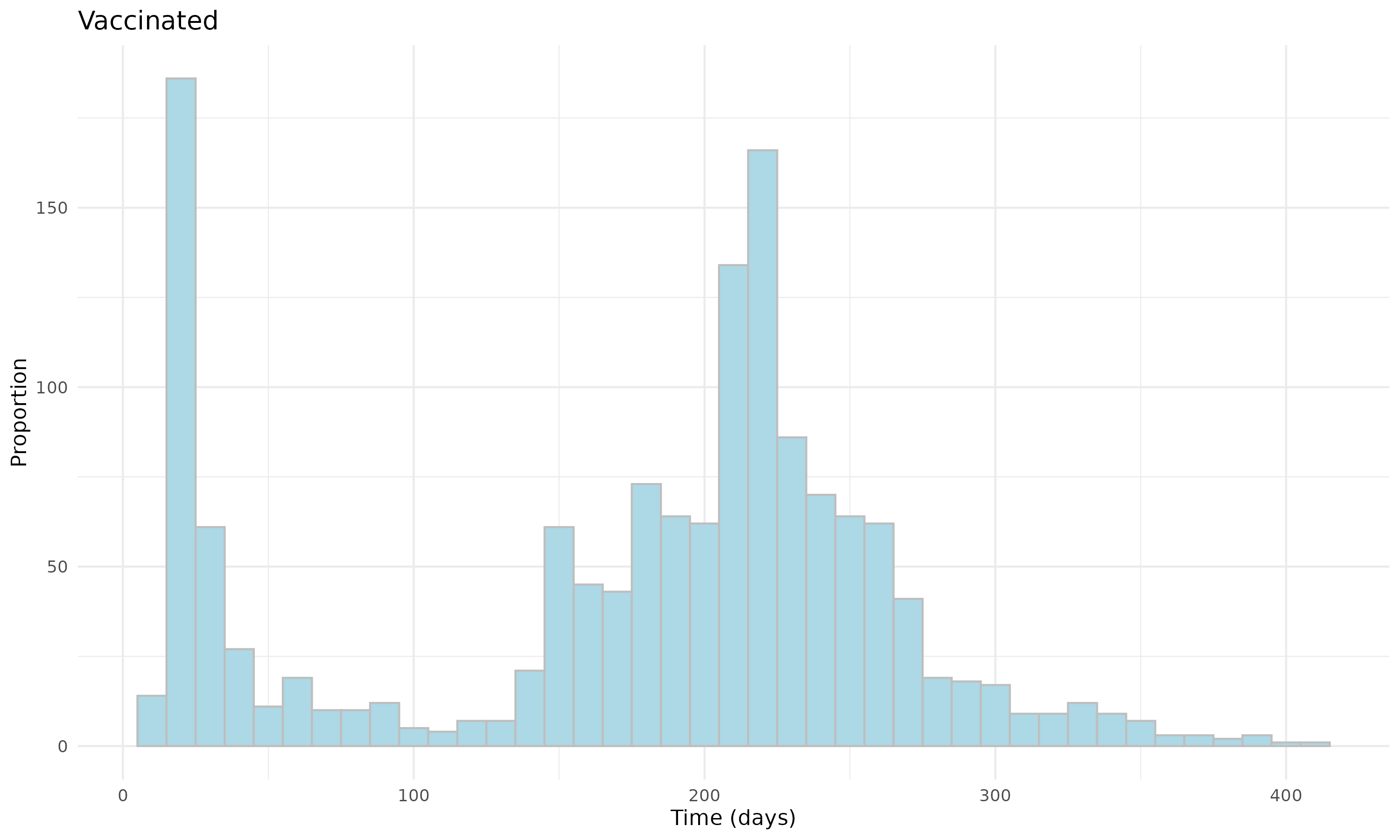

**Figure S28: Distribution of the time since the last COVID-19 immunization prior to Omicron B.A. 4/5 trial start date for vaccinated and unvaccinated individuals in Children Cohort.**

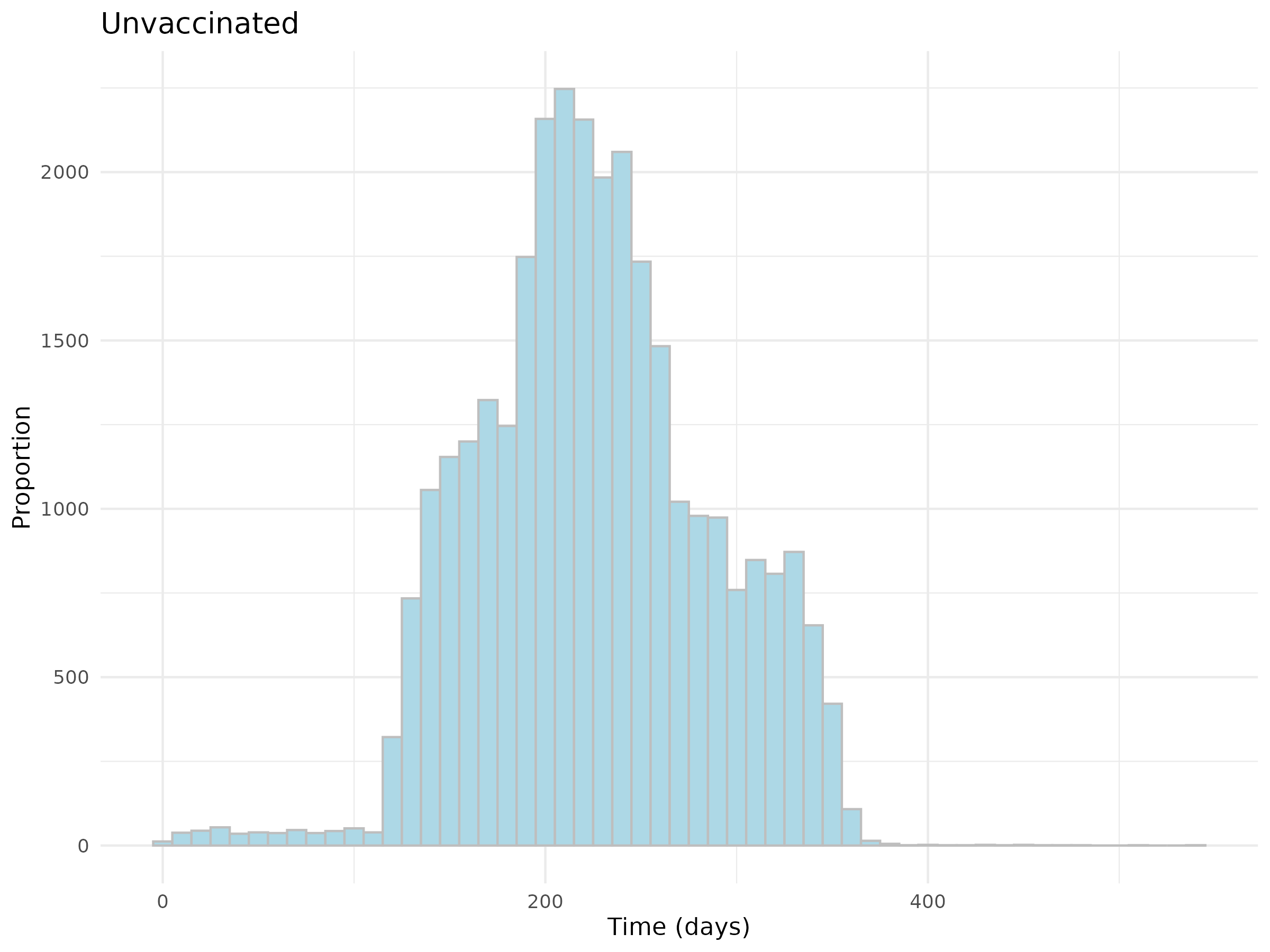

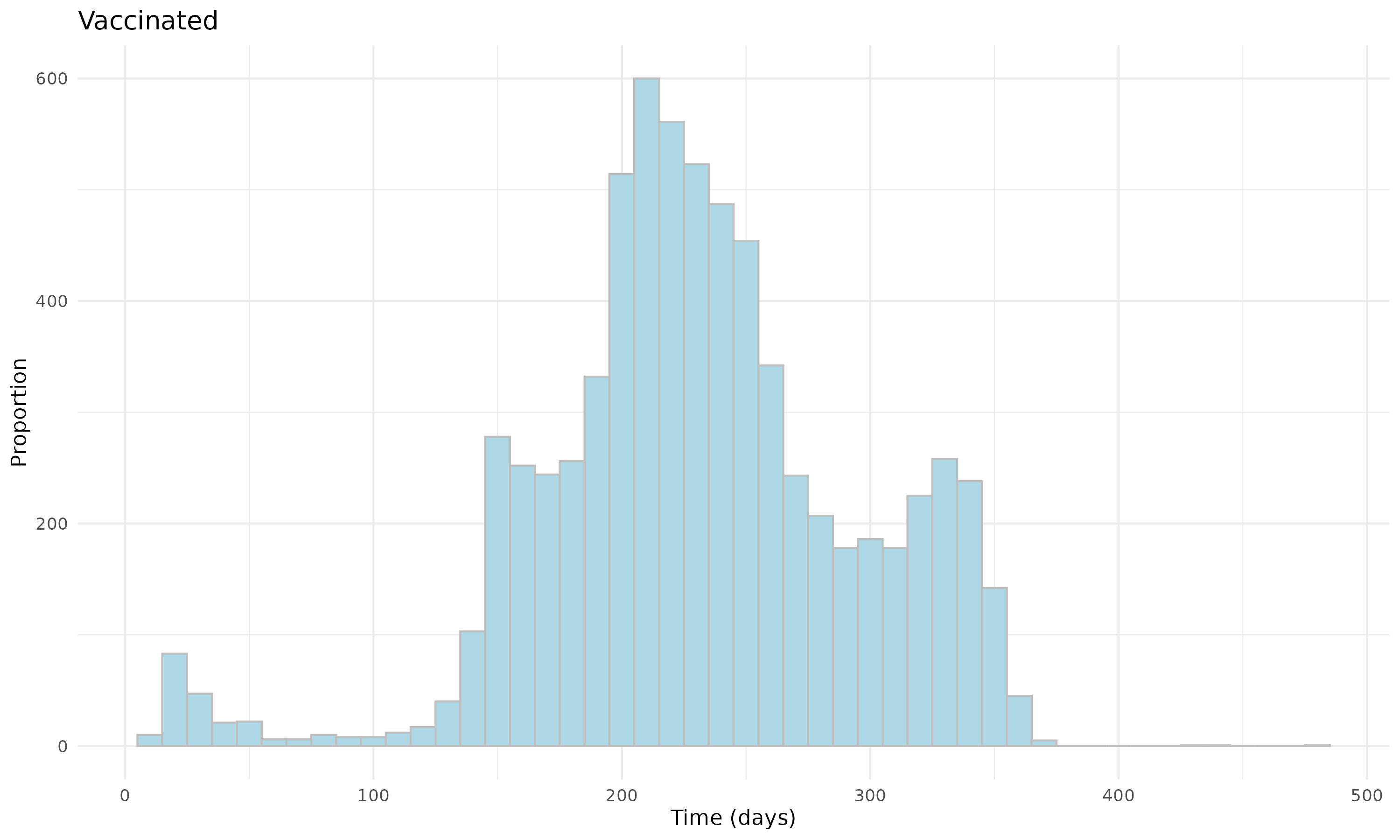

**Figure S29: Distribution of the time since the last COVID-19 immunization prior to Omicron B.A. 4/5 trial start date for vaccinated and unvaccinated individuals in Adolescent Cohort.**

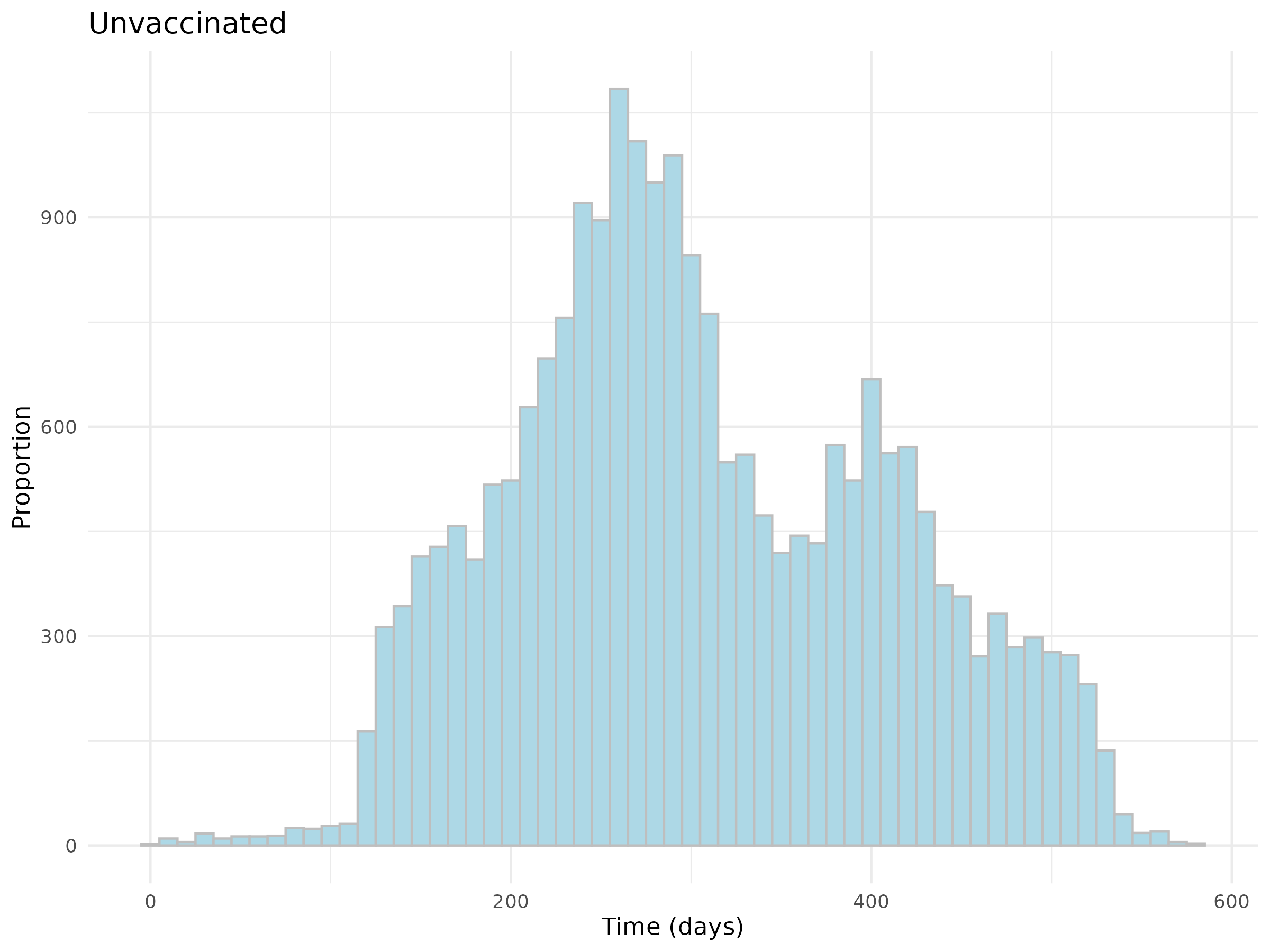

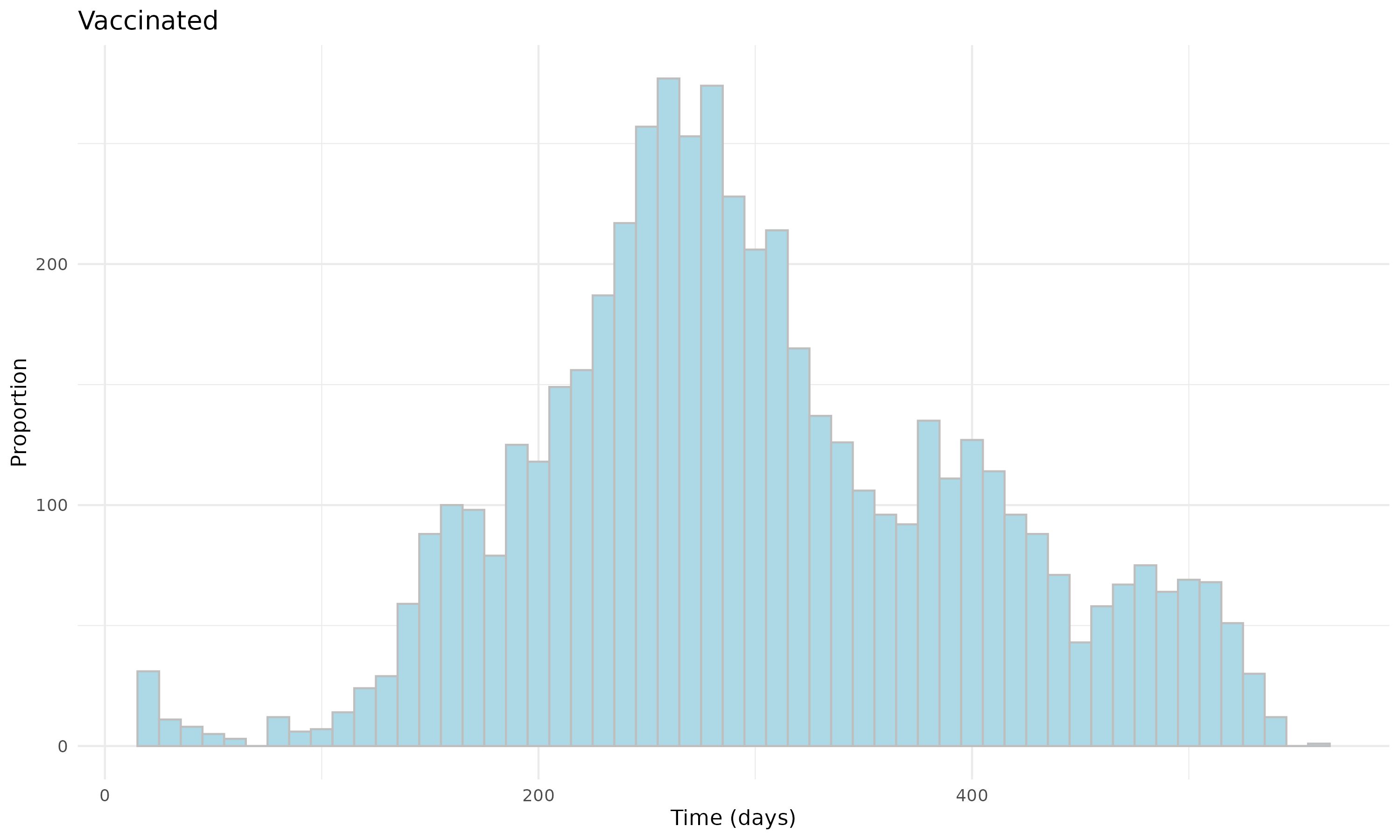

**Figure S30: Distribution of the time since the last COVID-19 immunization prior to Omicron XBB or later trial start date for vaccinated and unvaccinated individuals in Children Cohort.**

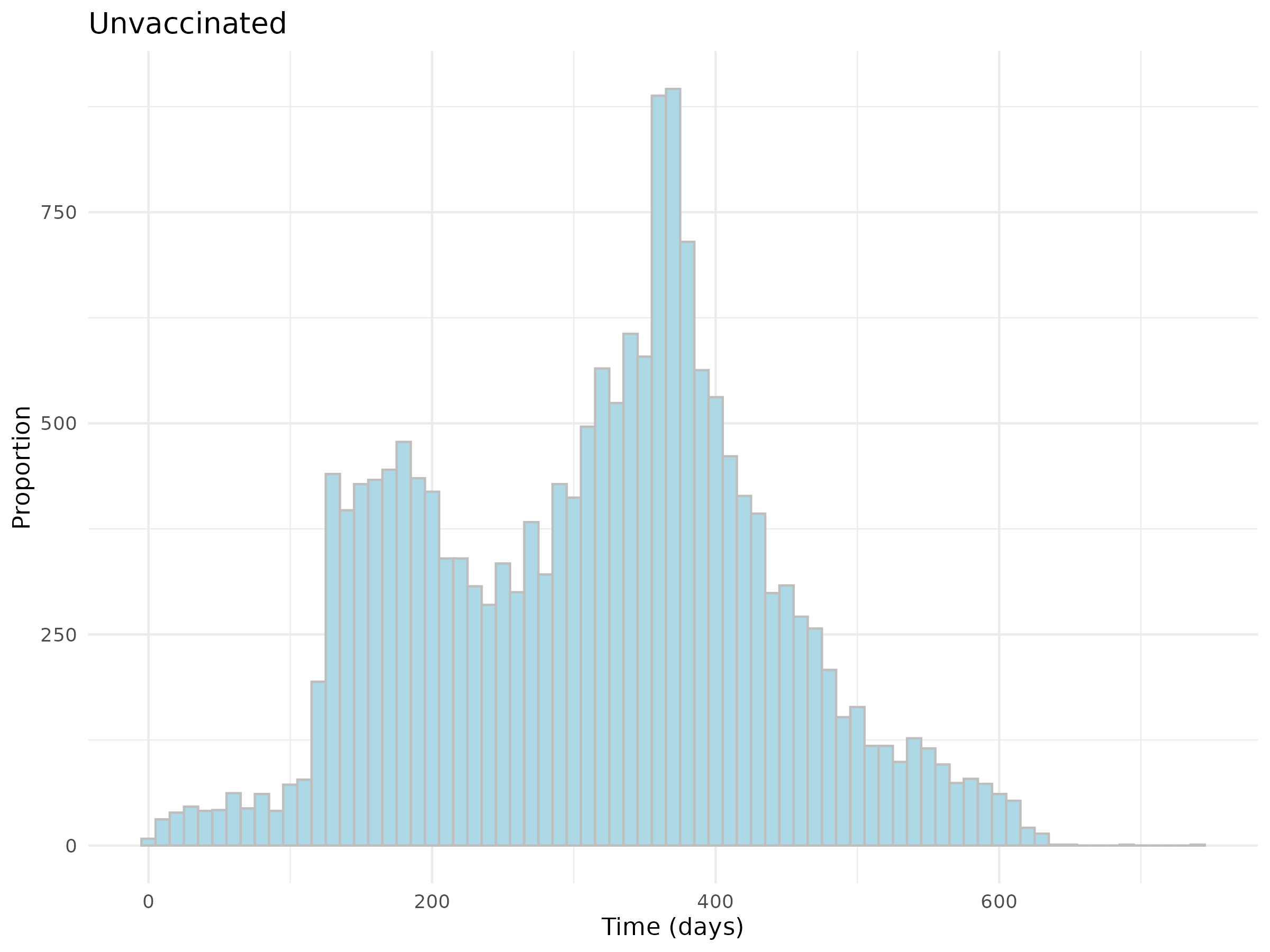

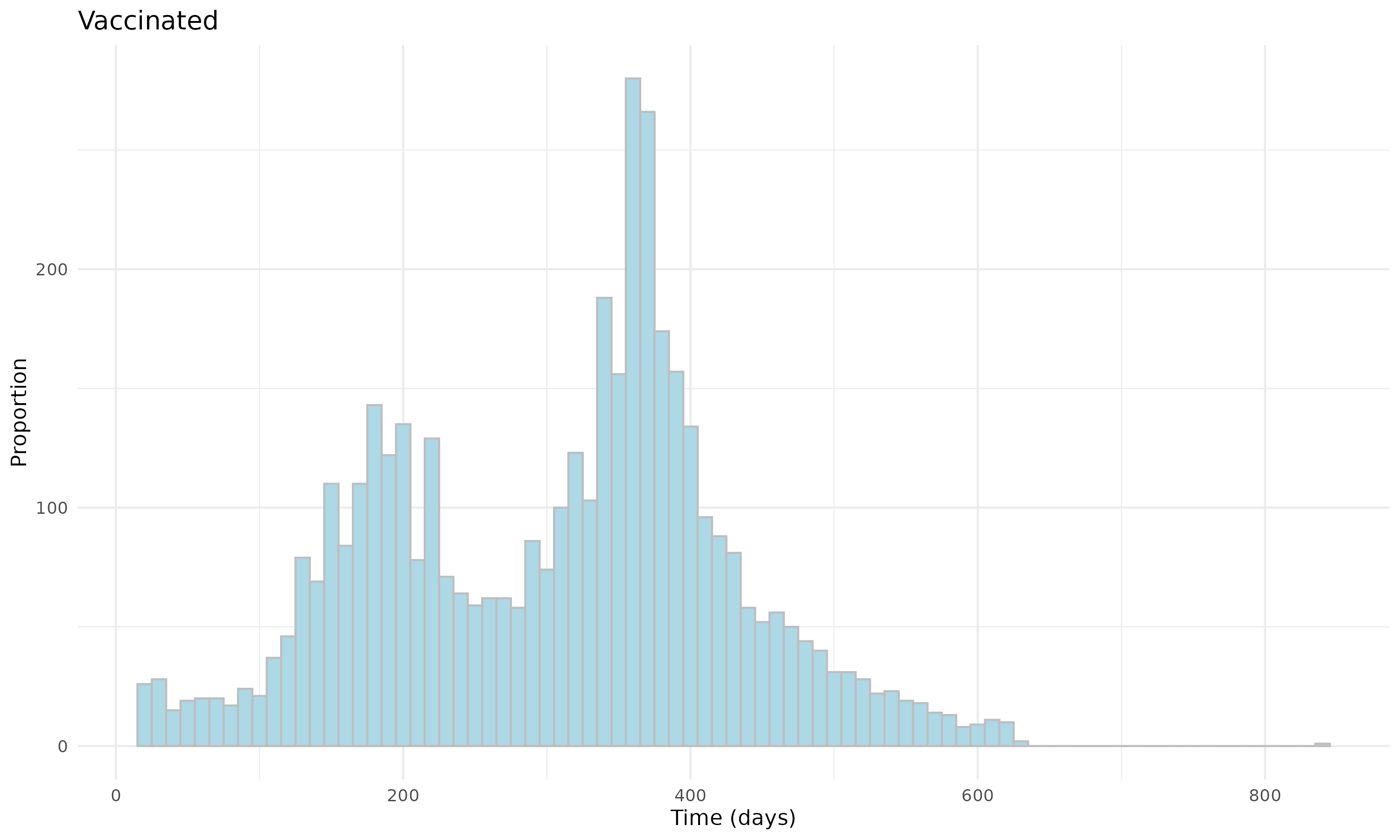

**Figure S31: Distribution of the time since the last COVID-19 immunization prior to Omicron XBB or later trial start date for vaccinated and unvaccinated individuals in Adolescent Cohort.**

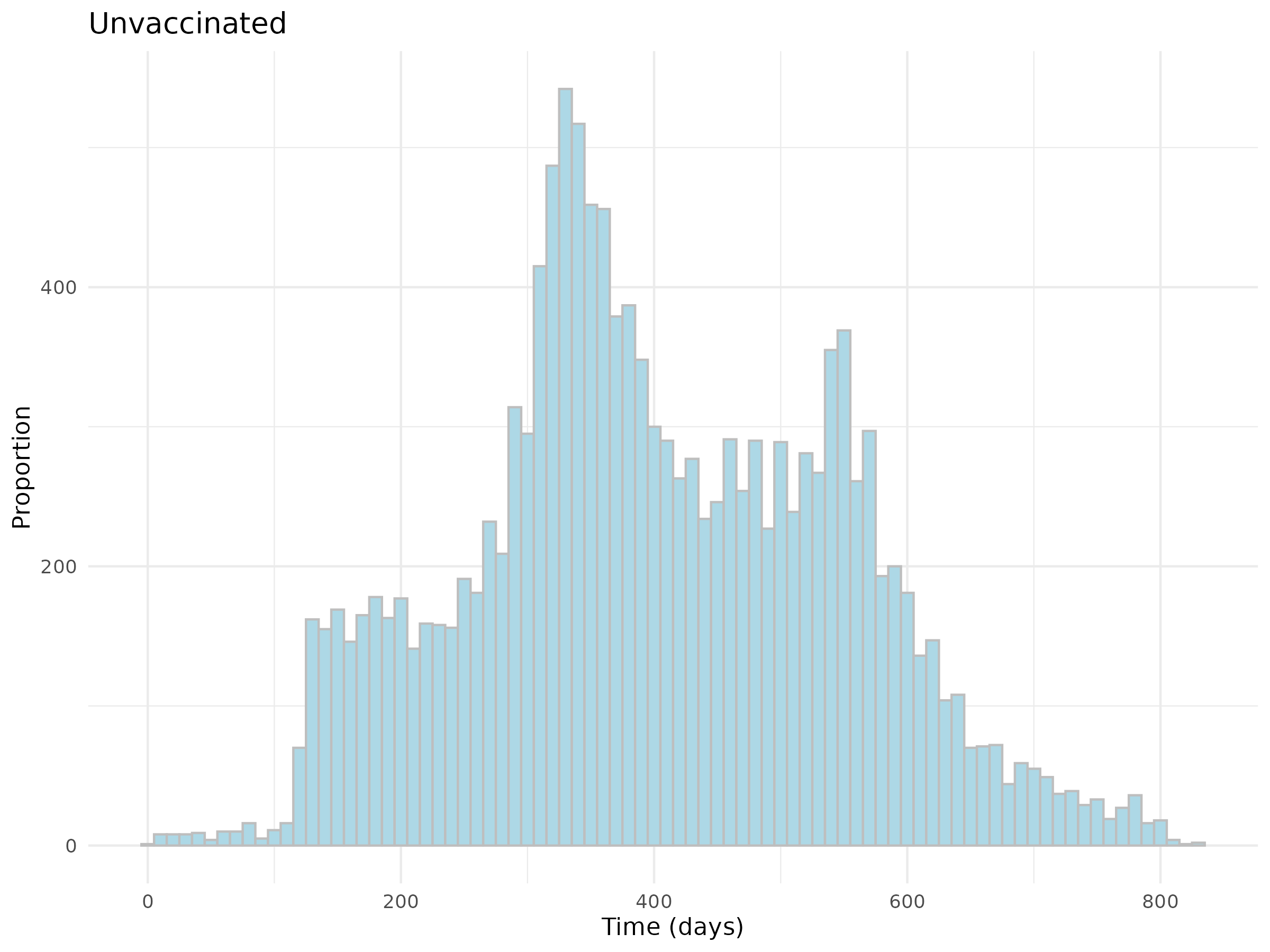

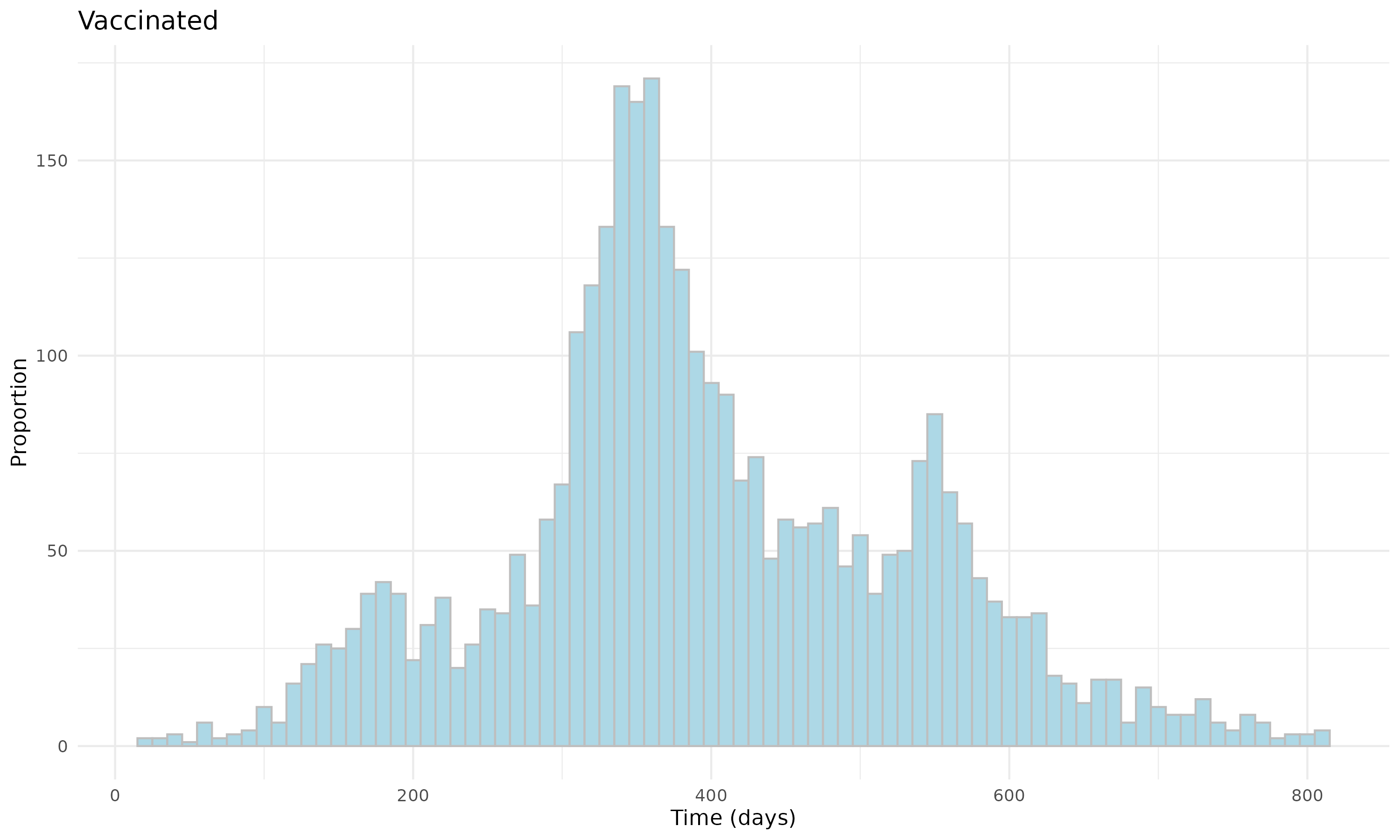

#### G. Survival Curves

Additionally, we presented the survival curves (i.e., Kaplan-Meier curves) of documented SARS-CoV-2 reinfection for the unvaccinated (red) and vaccinated (blue) individuals after matching, **Figure S32–S37**. From the figures, the curves present the cumulative survival probabilities, and a steeper slope indicates a higher event rate (i.e., documented SARS-CoV-2 reinfection rate).

**Figure S32: Survival curves of documented SARS-CoV-2 infection for vaccinated and unvaccinated individuals in Omicron B.A. 1/2 Children Cohort.**
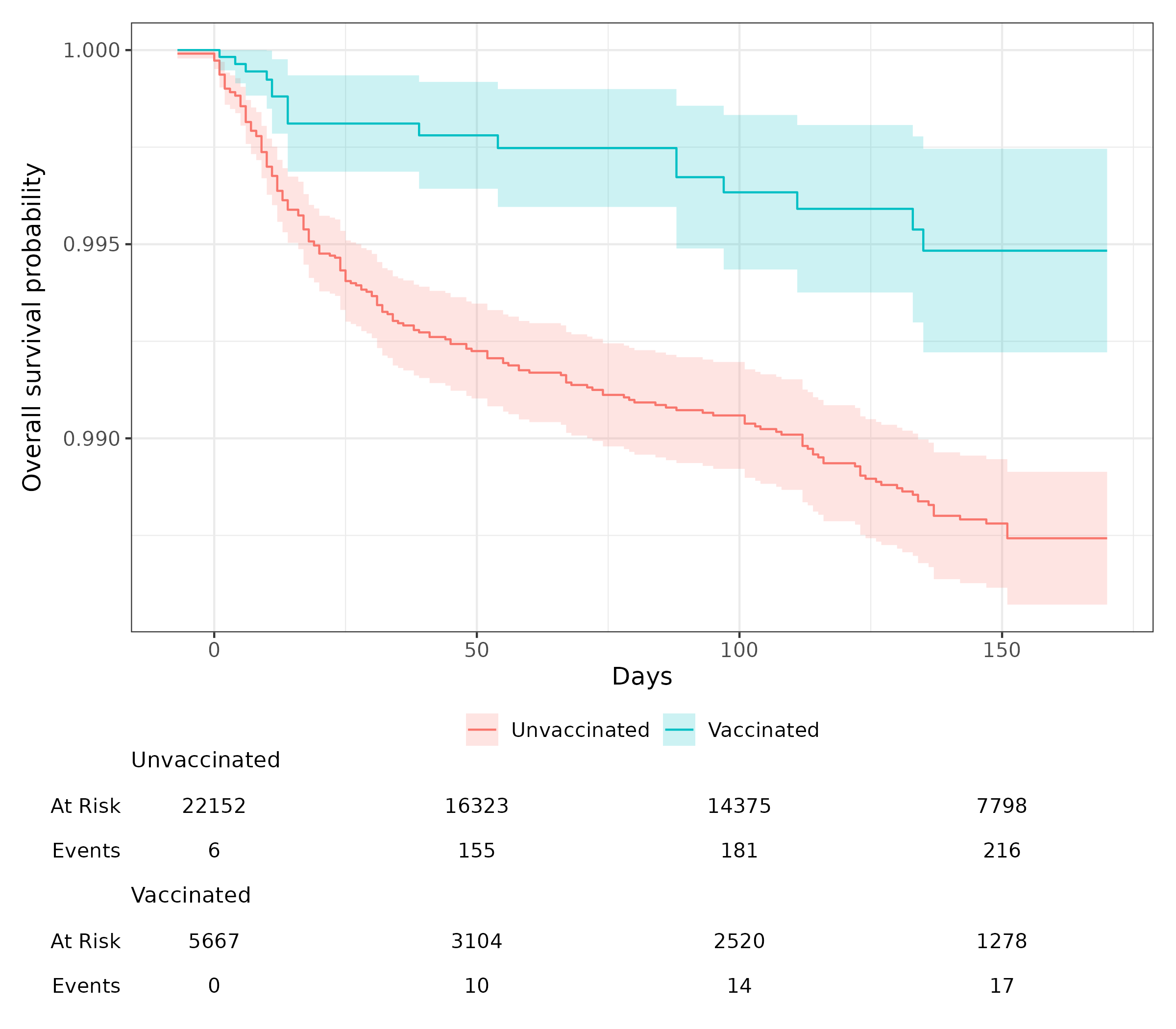

**Figure S33: Survival curves of documented SARS-CoV-2 infection for vaccinated and unvaccinated individuals in Omicron B.A. 1/2 Adolescent Cohort.**
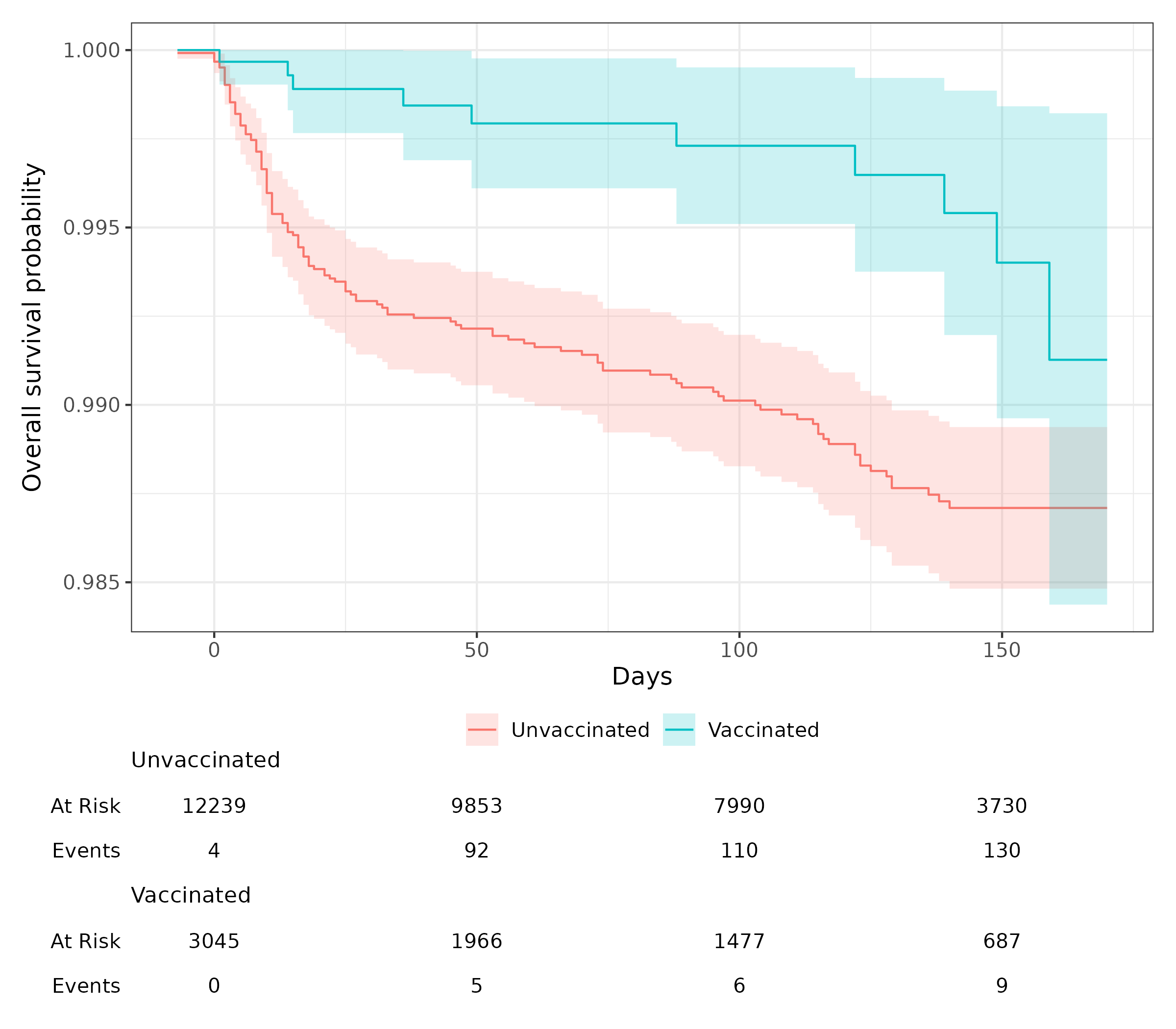

**Figure S34: Survival curves of documented SARS-CoV-2 infection for vaccinated and unvaccinated individuals in Omicron B.A. 4/5 Children Cohort.**
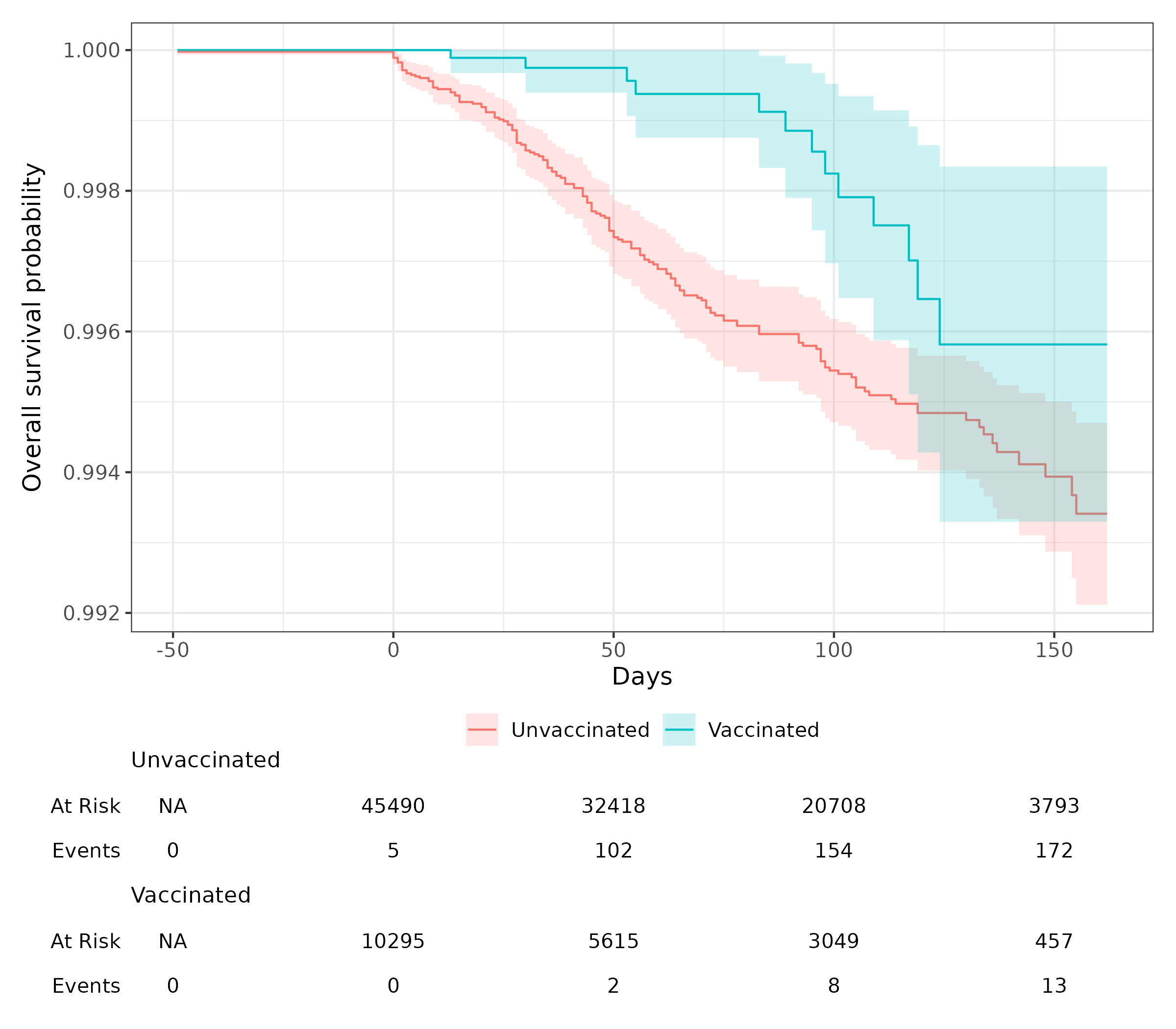

**Figure S35: Survival curves of documented SARS-CoV-2 infection for vaccinated and unvaccinated individuals in Omicron B.A. 4/5 Adolescent Cohort.**
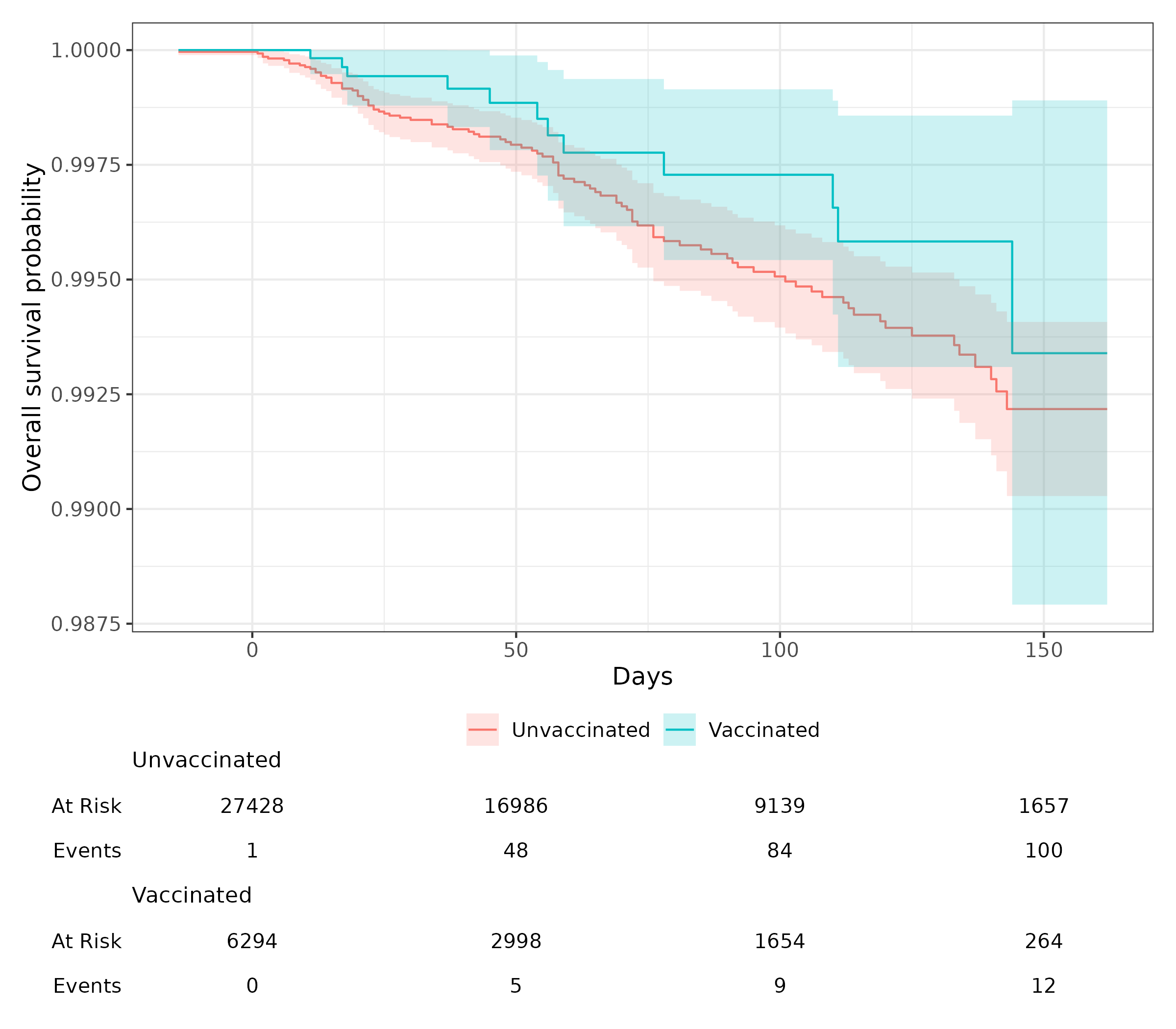

**Figure S36: Survival curves of documented SARS-CoV-2 infection for vaccinated and unvaccinated individuals in Omicron XBB or later Children Cohort.**
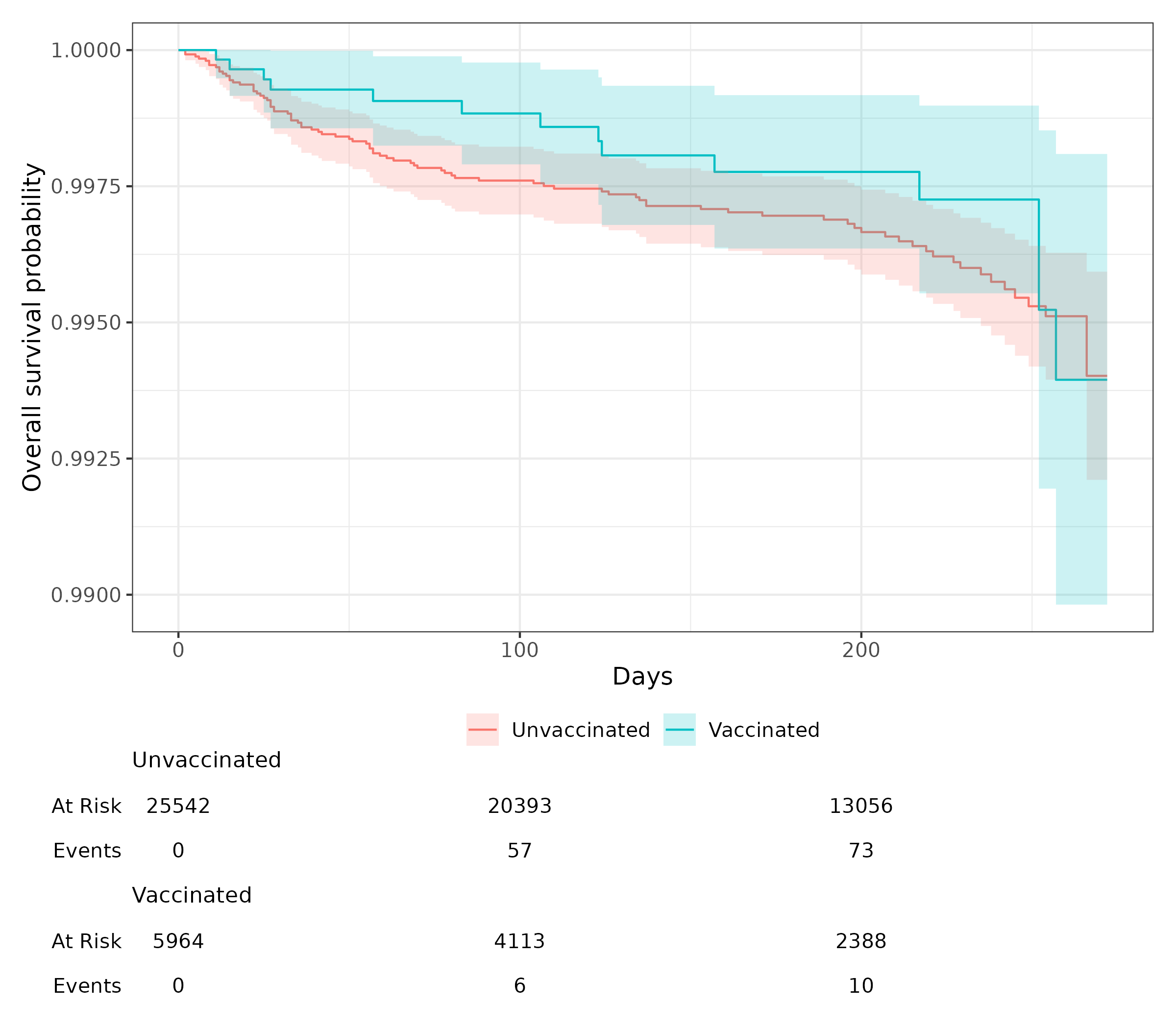

**Figure S37: Survival curves of documented SARS-CoV-2 infection for vaccinated and unvaccinated individuals in Omicron XBB or later Adolescents Cohort.**
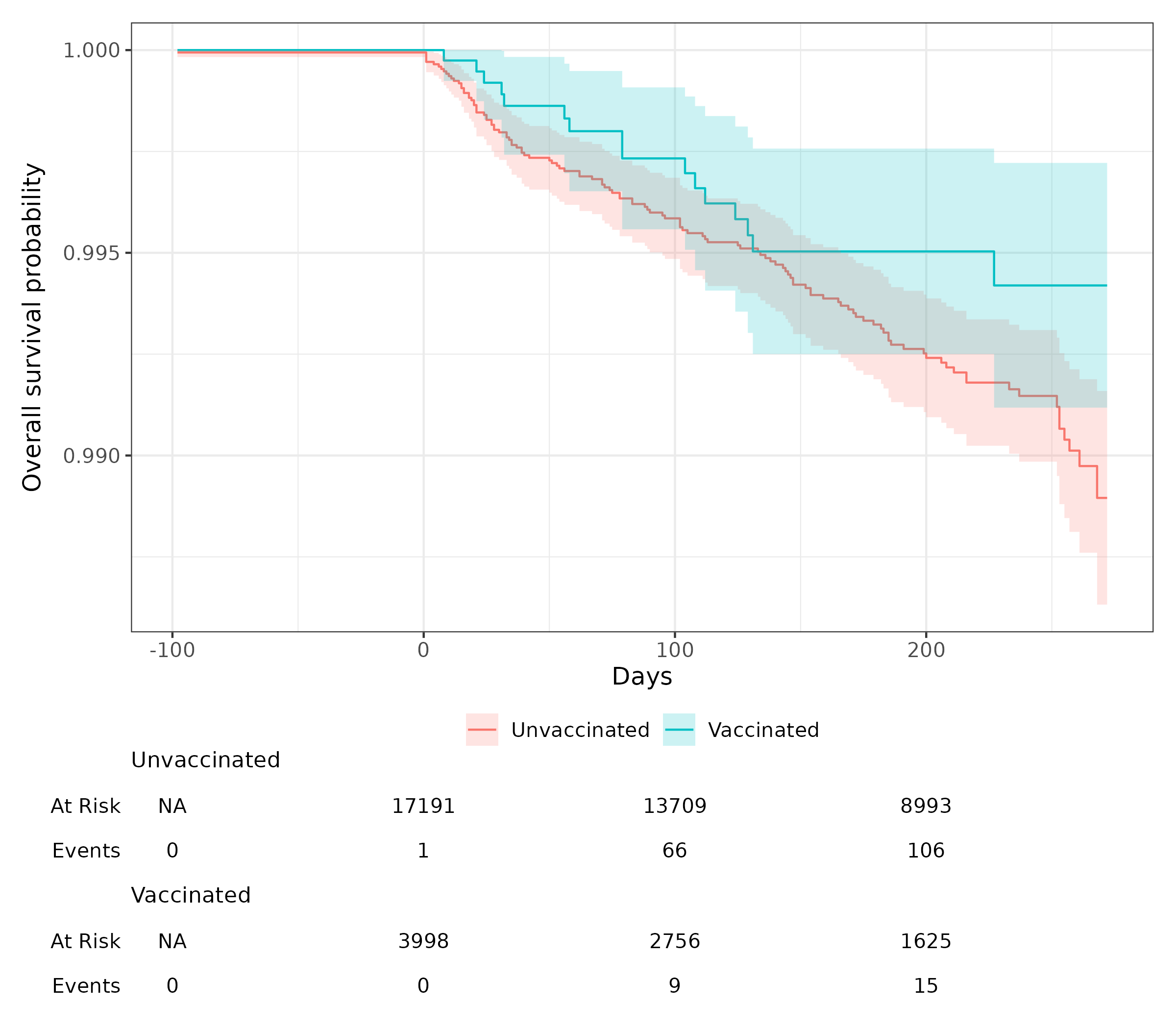

### Section S3 Supplemental Results

#### A. Stratification by last infected variants

To further explore and enrich the primary analysis and initial cohort design, we conducted a stratified analysis by previous SARS-CoV-2 variant infection to study the vaccine effectiveness in preventing reinfection with different variants. The results are summarized in **Table S2**. In addition, results with fewer than five cases were not included to protect patient privacy.

For adolescents who had previously been infected with the Delta variant, the vaccine exhibited an effectiveness of 64.5% (95% CI: 32.4% to 81.3%) against documented reinfection with the Omicron BA.1/2 variant. However, the effectiveness was not significant (-7.7%; 95% CI: -184.5% to 59.2%) against reinfection with the Omicron BA.4/5 variant. For those adolescents who had earlier been infected with the Omicron BA.1/2 variant, vaccine effectiveness was observed at 48.7% (95% CI: -1.5% to 74.1%) against reinfection with the Omicron BA.4/5 variant, and at 42.7% (95% CI: -14.2% to 71.2%) against reinfection with the Omicron XBB and subsequent variants. Among adolescents previously infected with the Omicron BA.4/5 variant, the vaccine did not show significant effectiveness (-40.2%; 95% CI: -225.6% to 39.7%) against reinfection with the XBB and later variants.

In the pediatric population previously infected with the Delta variant, the vaccine effectiveness was 62.3% (95% CI: 38.3% to 77.0%) against reinfection with the Omicron BA.1/2 variant. Among children previously infected with the Omicron BA.1/2 variant, the vaccine demonstrated an effectiveness of 59.1% (95% CI: 19.6% to 79.2%) against reinfection with the Omicron BA.4/5 variant, and 56.4% (95% CI: -8.9% to 82.5%) against reinfection with the Omicron XBB and subsequent variants

**Table S2**: Vaccine effectiveness for reinfection stratified by last infected variants

(a) For adolescents (age 12-18):

| Previous Infected Variant | Study Wave | Infected Count (Total)  % in Vaccinated Group | Infected Count (Total)  % in Unvaccinated Group | VE (95% CI) |
| --- | --- | --- | --- | --- |
| Delta | Omicron B.A. 1/2 | 15 (3,045) 0.49% | 130 (12,240) 1.06% | 0.645 (0.324 – 0.813) |
|  | Omicron B.A. 4/5 | 7 (1,410) 0.50% | 22 (5,809) 0.38% | -0.077 (-1.845 – 0.592) |
|  | XBB or later | - | 13 (2,747) 0.47% | 0.645 (-1.648 – 0.955) |
| Omicron B.A. 1/2 | Omicron B.A. 4/5 | 11 (4,804) 0.23% | 96 (21,151) 0.45% | 0.487 (-0.015 – 0.741) |
|  | XBB or later | 9 (2,228) 0.40% | 80 (9,983) 0.80% | 0.427 (-0.142 – 0.712) |
| Omicron B.A. 4/5 | XBB or later | 7 (933) 0.75% | 24 (3,772) 0.64% | -0.402 (-2.256 – 0.397) |

(b) For children (age 5-11):

| Previous Infected Variant | Study Wave | Infected Count (Total)  % in Vaccinated Group | Infected Count (Total)  % in Unvaccinated Group | VE (95% CI) |
| --- | --- | --- | --- | --- |
| Delta | Omicron B.A. 1/2 | 21 (5,667) 0.37% | 219 (22,154) 0.99% | 0.623 (0.383 – 0.770) |
|  | Omicron B.A. 4/5 | - | 36 (12,209) 0.29% | 0.856 (-0.054 – 0.980) |
|  | XBB or later | - | 13 (5,126) 0.25% | -0.151 (-3.049 – 0.673) |
| Omicron B.A. 1/2 | Omicron B.A. 4/5 | 13 (7,322) 0.18% | 123 (32,071) 0.38% | 0.591 (0.196 – 0.792) |
|  | XBB or later | 5 (3,342) 0.15% | 57 (14,534) 0.39% | 0.564 (-0.089 – 0.825) |
| Omicron B.A. 4/5 | XBB or later | - | 20 (4,623) 0.43% | 0.550 (-0.927 – 0.895) |

#### B. Vaccine effectiveness for symptomatic illness

In addition to the primary analysis assessing vaccine effectiveness in preventing reinfection with various Omicron subvariants in a pediatric cohort previously infected with different SARS-CoV-2 variants, we also studied vaccine effectiveness in preventing symptomatic illness. **Table S3** summarizes the number and incidence rates of COVID-19 severity outcome (asymptomatic, mild, moderate, severe) and the uninfection in each study cohorts. Given the rarity of severe outcomes, we consolidated mild, moderate, and severe cases into a single binary variable to indicate the presence or absence of symptomatic illness. Given that severity outcomes are rare, we combined mild, moderate, and severe outcomes into a single binary variable, indicating the presence or absence of any symptomatic illness. The results are presented in **Figure S38**.

The COVID-19 vaccine demonstrated positive effectiveness in preventing symptomatic COVID-19 illness for cohorts infected with Omicron BA.1/2 and BA.4/5. However, the vaccine showed marginal negative effectiveness for Omicron XBB or later Cohorts. The wider confidence intervals and negative results observed for the Omicron XBB, or later Cohorts may be attributable to the small number of detected events.

**Table S3. Symptomatic reinfection of SARS-CoV-2**

| Study Cohort | BA.1/2 | | | | BA.4/5 | | | | XBB or Later | | | |
| --- | --- | --- | --- | --- | --- | --- | --- | --- | --- | --- | --- | --- |
| Age Constraint | **Children** | | **Adolescents** | | **Children** | | **Adolescents** | | **Children** | | **Adolescents** | |
| Vaccination | Vacc. | Unvacc. | Vacc | Unvacc | Vacc | Unvacc | Vacc | Unvacc | Vacc | Vacc | Unvacc | Unvacc |
| Severity | | | | | | | | | | | | |
| Asymptomatic | 27 (0.40%) | 565  (1.36%) | 20  (0.54%) | 472  (1.33%) | 9  (0.08%) | 533  (0.48%) | 14  (0.20%) | 522  (0.53%) | 11  (0.16%) | 335  (0.23%) | 12  (0.26%) | 545  (0.43%) |
| Mild | 13  (0.19%) | 331  (0.80%) | 10  (0.27%) | 322  (0.90%) | 12  (0.11%) | 447 (0.40%) | 14 (0.20%) | 361 (0.36%) | 13 (0.19%) | 511 (0.35%) | 11 (0.24%) | 508  (0.40%) |
| Moderate | - | 33  (0.08%) | - | 30  (0.08%) | - | 57  (0.05%) | - | 48  (0.05%) | - | 78 (0.05%) | - | 62  (0.05%) |
| Severe | - | 12  (0.03%) | - | 13  (0.04%) | - | 29 (0.03%) | - | 29  (0.03%) | - | 25 (0.02%) | - | 26  (0.02%) |
| Uninfected | 6,738  (99.34%) | 40,525  (97.73%) | 3,686  (99.17%) | 34,770  (97.65%) | 11,384  (99.75%) | 110,939  (99.05%) | 6,952  (99.57%) | 97,967  (99.03%) | 6,702 (99.54%) | 144,279 (99.35%) | 4,616 (99.44%) | 125,237 (99.10%) |

**Figure S38. Vaccine effectiveness against symptomatic illness (Mild/Moderate/Sever) of COVID-19.**

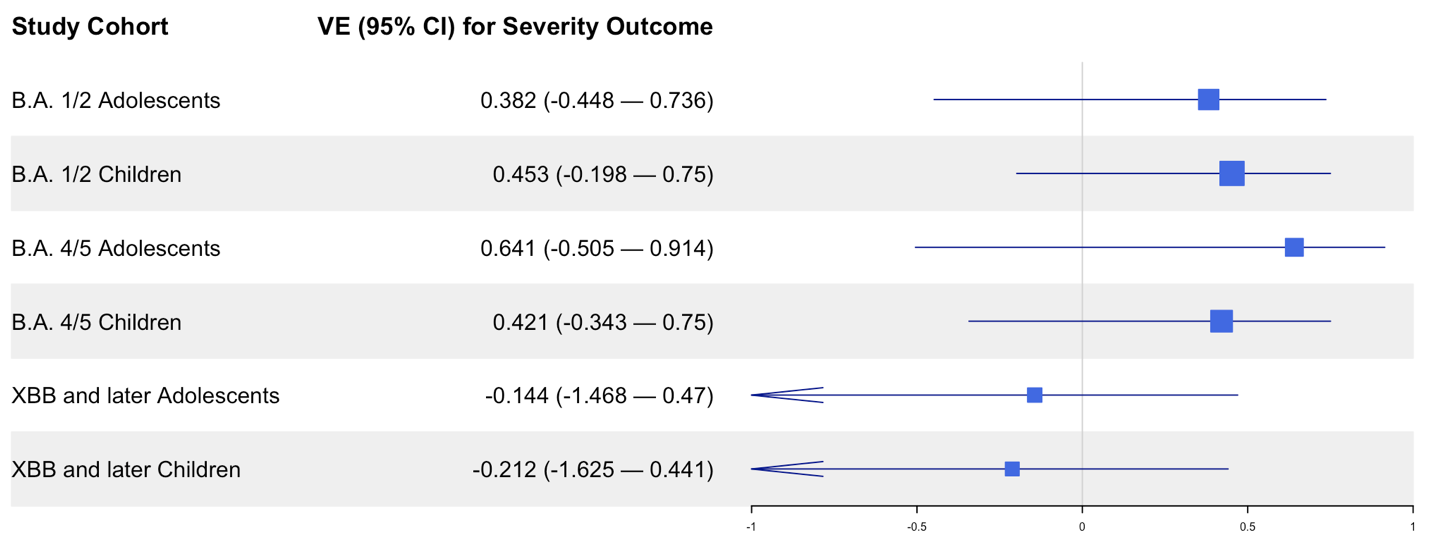

#### C. Vaccine effectiveness stratified by Pfizer

As shown in **Figure S19**, Pfizer is the predominant vaccine brand among vaccinated individuals in our study cohorts. To further investigate, we restricted the vaccination group to individuals who received the Pfizer vaccine exclusively and estimated vaccine effectiveness by comparing them with matched comparator groups within each study cohort. The results of this sensitivity analysis, presented in **Figure S39**, were generally consistent with the primary outcome analysis (**Table 3**), confirming similar patterns of vaccine effectiveness across study cohorts.

**Figure S39. Vaccine effectiveness for Pfizer brand only**
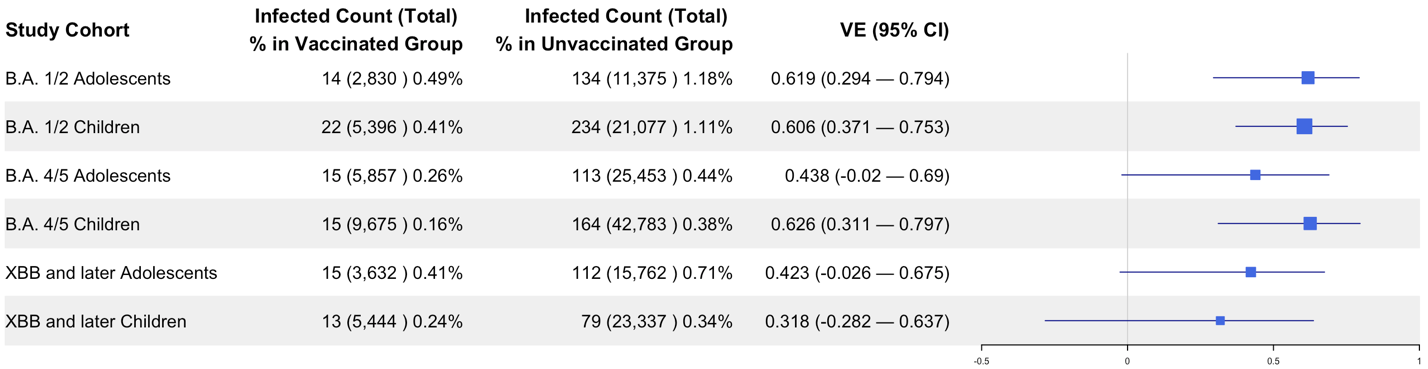

#### D. Negative Control Experiments

To evaluate the robustness of our study, we utilized a list of 40 negative control outcomes to conduct the negative control experiments. In this investigation, negative control outcomes are defined as clinical outcomes believed to have no causal relationship to the exposure. The full list of outcomes for our research was designated by pediatric physicians. A comprehensive list of these outcomes can be found in **Table S4**.

For these 40 negative control outcomes, the null hypothesis is that the exposure (i.e., COVID-19 vaccination) has no effect on these outcomes. Given the potential existence of systematic error, these negative control outcomes are used to assess the performance of our study design and contribute to calibrating our estimated VE^2,3^. Specifically, we initially estimated the empirical null distribution of these negative control outcomes, employing the identical CER method utilized for VE estimation. Subsequently, this empirical null distribution served to calibrate the effectiveness estimates of the primary analysis. Further details on the estimation of empirical null distribution and the calibration procedure are available in Schuemie et al. 2014 and Schuemie et al. 2018.

**Table S4. Negative control outcomes.**

Health conditions

Wax in ear/impacted cerumen

Snoring/Obstructive sleep apnea

Contact dermatitis

Injury of head

Diaper rash

Seizure

Speech delay

Autism/Autistic disorder

Displacements - bone

Closed fractue of distal end of radius

Acne

Falls

Visual testing abnormal

Sprain of ankle

Concussion

Impetigo

Scoliosis

Foot pain

Injury of free lower limb

Injury of. Upper extremity

Speech dysfunction

Umbilical hernia

Insect bite

Myopia

Injury of finger

Injury of right leg

Astigmatism

Injury of left leg

Tinea capitis

Obesity

Injury of right hand

Tinea corporis

Epilepsy

Tongue tie

Plagiocephaly

Inguinal hernia

Pain in wrist

Closed injury of head

Foreign body in ear

Injury of right foot

**Figure S40–S45** show the estimated hazard ratios and corresponding standard errors for these negative control outcomes for each study cohort. The vaccine effectiveness after NCO calibration as presented in **Figure S46**, reveals a small systematic error and residual bias of the study.

**Figure S40. Systematic error control when estimating vaccine effectiveness of B.A. 1/2 Children Cohort.** The plot displays HRs and their corresponding standard errors for each negative control outcome.

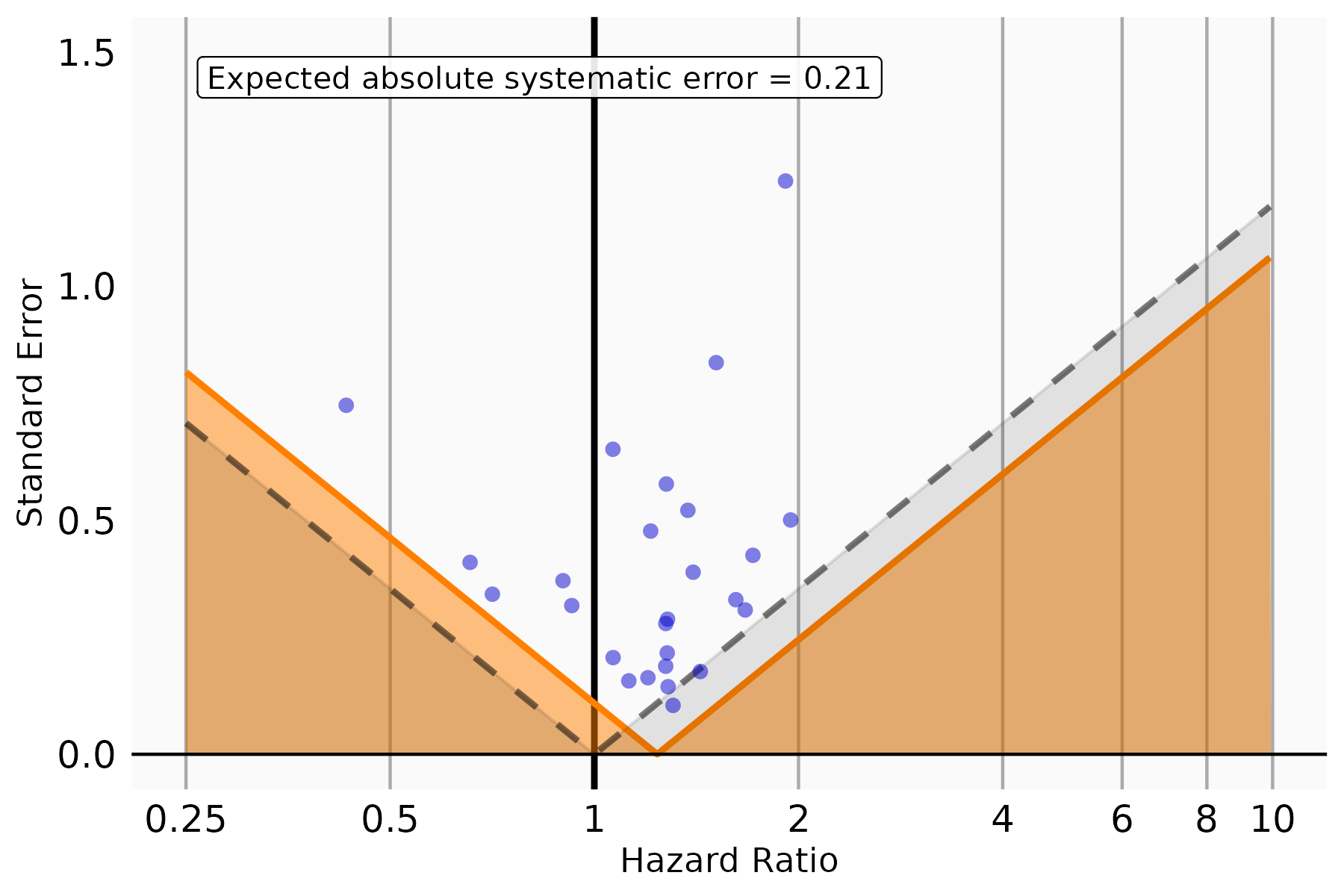

**Figure S41. Systematic error control when estimating vaccine effectiveness of B.A. 1/2 Adolescents Cohort.** The plot displays HRs and their corresponding standard errors for each negative control outcome.

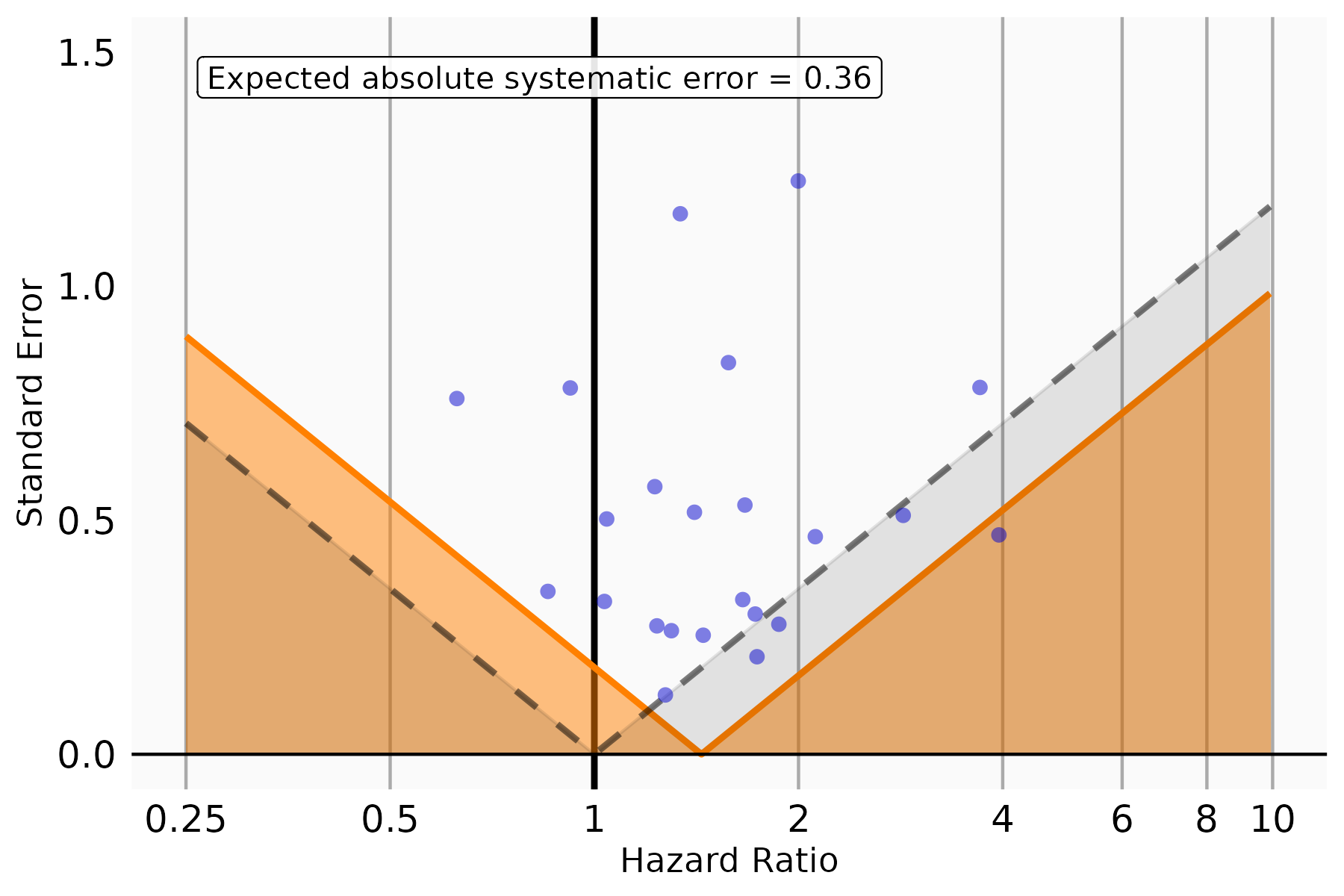

**Figure S42. Systematic error control when estimating vaccine effectiveness of B.A. 4/5 Children Cohort.** The plot displays HRs and their corresponding standard errors for each negative control outcome.

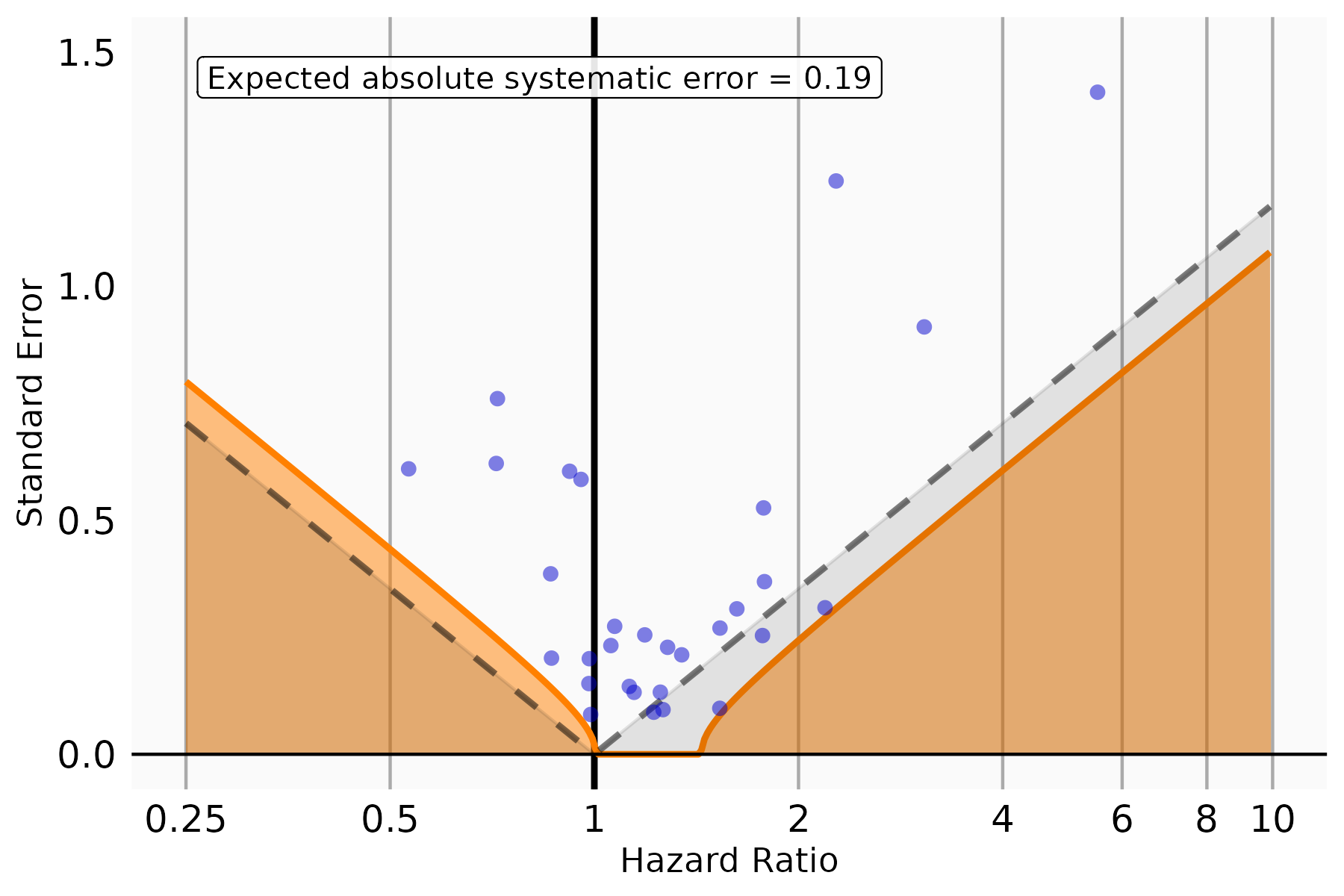

**Figure S43. Systematic error control when estimating vaccine effectiveness of B.A. 4/5 Adolescents Cohort.** The plot displays HRs and their corresponding standard errors for each negative control outcome.

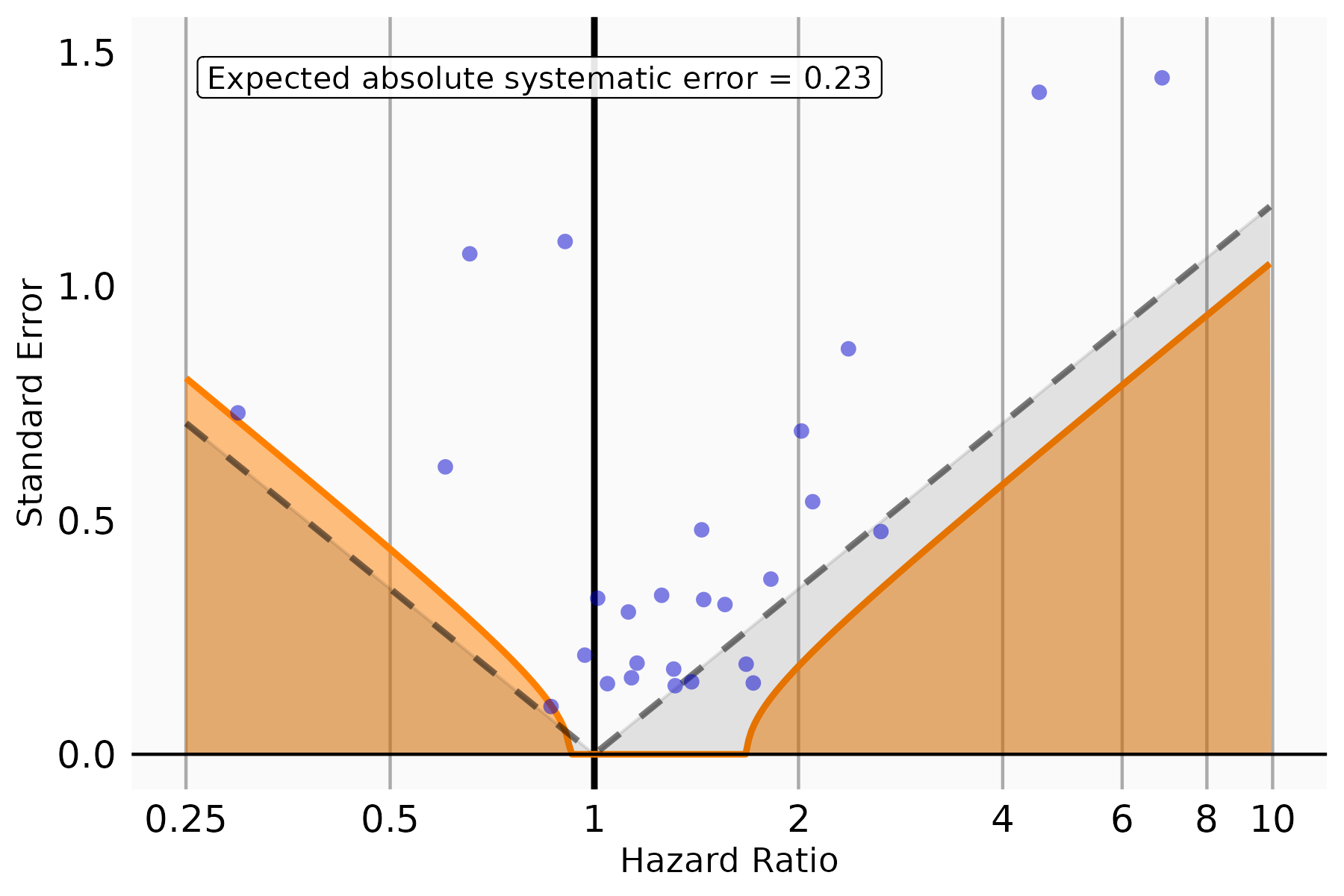

**Figure S44. Systematic error control when estimating vaccine effectiveness of XBB or later Children Cohort.** The plot displays HRs and their corresponding standard errors for each negative control outcome.

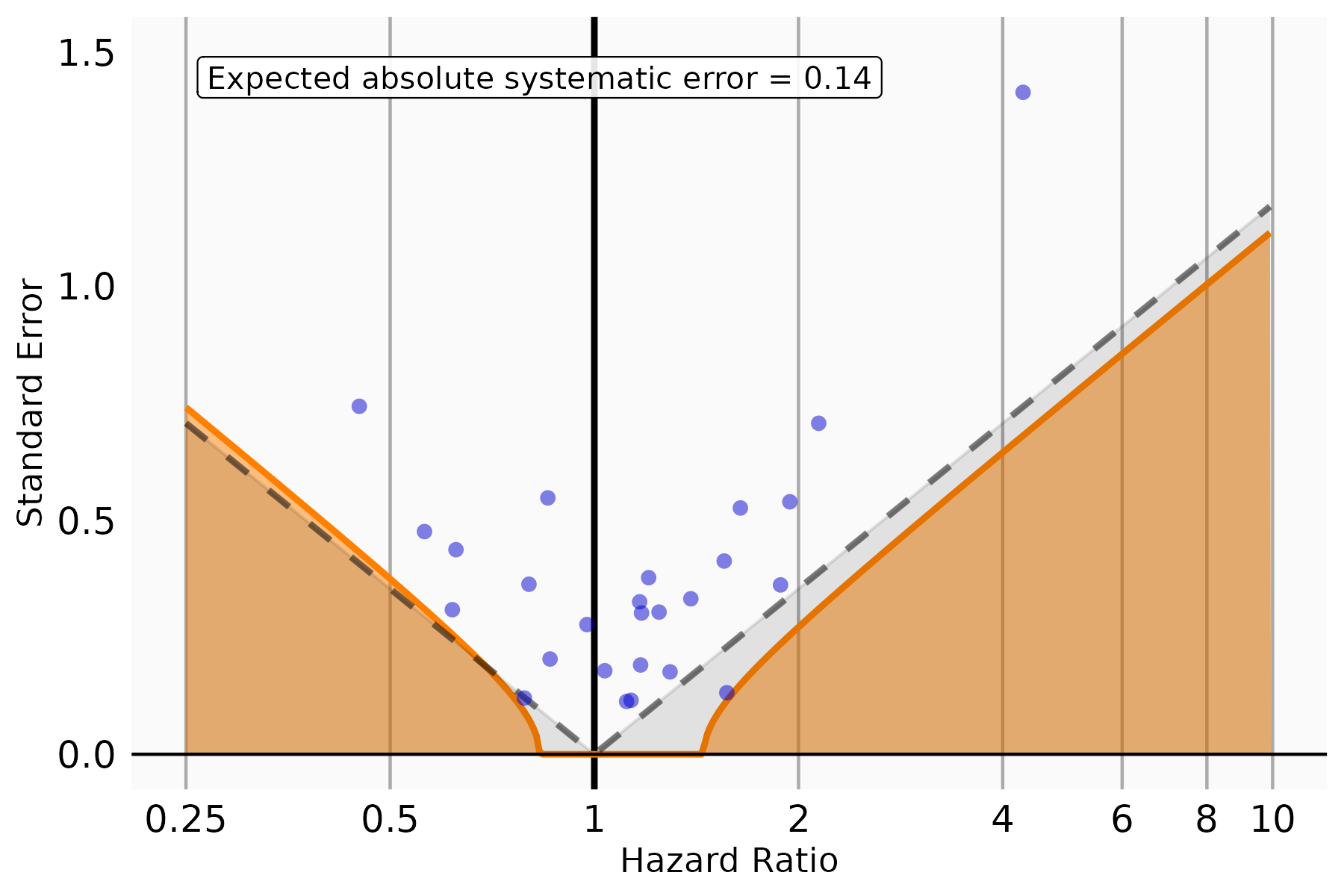

**Figure S45. Systematic error control when estimating vaccine effectiveness of XBB or later Adolescents Cohort.** The plot displays HRs and their corresponding standard errors for each negative control outcome.

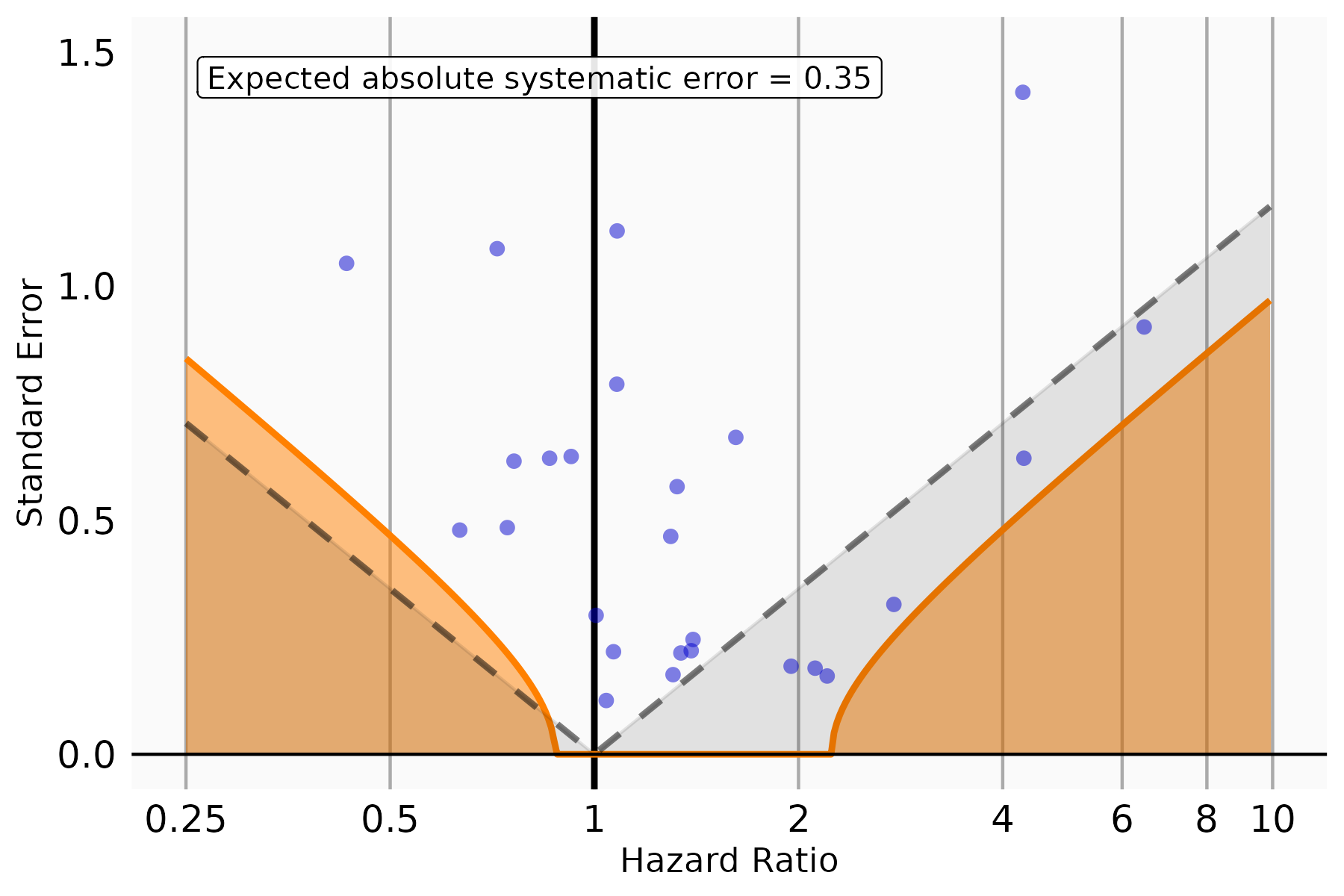

**Figure S46.** Vaccine effectiveness and 95% CI of BNT162b2 vaccine against SARS-CoV-2 Infection after negative control calibration for each study cohort

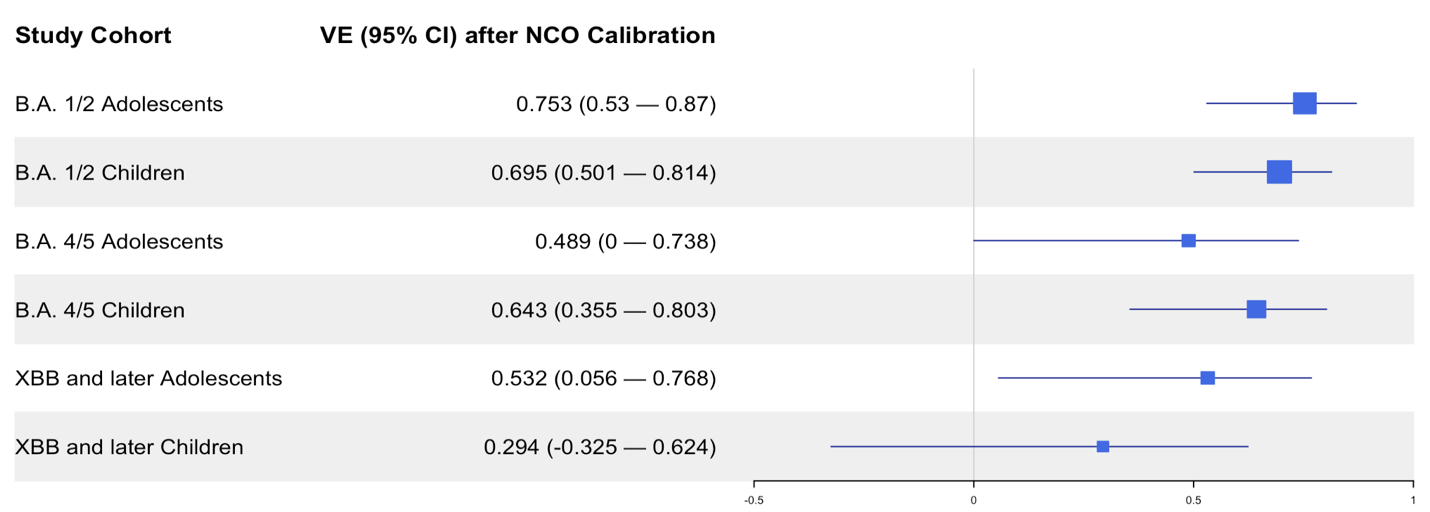
